# A causal link between autoantibodies and neurological symptoms in long COVID

**DOI:** 10.1101/2024.06.18.24309100

**Authors:** Keyla Santos Guedes de Sá, Julio Silva, Rafael Bayarri-Olmos, Christopher A. Baker, Zhenni Lu, Wilson Gipson, Daxiang Na, Bandy Chen, Li Wenxue, Delyar Khosroabadi, Ryan Brinda, Robert Alec Rath Constable, Britney Omene, Patricia A. Colom Díaz, Dong-il Kwon, Gisele Rodrigues, Harald Heidecke, Kai Schulze-Forster, Amanda Gross, Tom Shneer, Amanda Clarke, Thomas Linnekin, Ashley Brate, Lev Brown, Henry Buda, Shashi Jatiani, Lenny Moise, Kerrie Greene, Sachin Bhagchandani, Bornali Bhattacharjee, Jeffrey Gehlhausen, Jamie Wood, Laura Tabacof, Carmen Scheibenbogen, Yansheng Liu, Leying Guan, Marc Schneeberger Pane, David Putrino, Tamas L. Horvath, Akiko Iwasaki

**Affiliations:** Department of Immunobiology, Yale School of Medicine, Center for Infection and Immunity, New Haven, CT 06519, USA; Center for Infection and Immunity, Yale School of Medicine, New Haven, CT 06519, USA; Cohen Center for Recovery from Complex Chronic Illness, Icahn School of Medicine at Mount Sinai, New York, NY10029, USA; Department of Comparative Medicine, Yale University School of Medicine, New Haven, CT 06520, USA; Howard Hughes Medical Institute, Chevy Chase, MD 20815, USA; Department of Cellular & Molecular Physiology, Yale University School of Medicine, New Haven, CT 06520, USA; Department of Pathology, Yale University School of Medicine, New Haven, CT 06520, USA; Yale Cancer Biology Institute, Yale University, West Haven, CT 06516, USA; Department of Biostatistics, Yale School of Public Health, New Haven, CT 06510, USA; SeromYx Systems, Woburn, Massachusetts, 01801, USA; CellTrend GmbH, Luckenwalde, 14943 Germany; Institute of Medical Immunology, Charité - Universitätsmedizin Berlin, Corporate Member of Freie Universität Berlin and Humboldt Universität zu Berlin and Berlin Institute of Health (BIH), Berlin, Germany

## Abstract

Acute SARS-CoV-2 infection triggers the de novo production of diverse, functional autoantibodies (AABs) that remain elevated in Long COVID (LC), but their pathogenic role remains unclear. Using tissue-based immunofluorescence, ELISA, human protein array, and mass spectrometry assays, we identified a broad range of AAB targets among individuals with LC. Individuals with neurocognitive symptoms showed increased AABs against central and peripheral nervous system proteins. Purified IgG reacted with human locus coeruleus, thalamus, adrenal gland, thyroid, and cross-reacted with mouse sciatic nerve and meninges. CNS-reactive AABs correlated with several neurological symptoms. MED20-targeting IgG from patients with LC showed enhanced antibody-dependent phagocytosis. Passive transfer of IgG from individuals with LC into mice induced fatigue-like behavior, loss of balance/coordination, thermal hyperalgesia, small fiber nerve damage, and increased pain-related neuronal activity, recapitulating patients’ symptoms. These findings suggest that targeting AABs might offer therapeutic benefits for this LC subgroup.

## Introduction

Long COVID (LC) develops in over 10% of individuals after a SARS-CoV-2 infection^1–5^. A wide range of neurological symptoms has debilitating effects on people with LC, impacting multiple regions of the brain^6,7^. Women are more likely than men to develop LC and other post-acute infection syndromes (PAIS), including myalgic encephalomyelitis/chronic fatigue syndrome (ME/CFS)^8,9^ , affecting >1% of Americans^10,11^, suggesting sex differences in the mechanisms that cause these disorders^12–15^. Since many autoimmune diseases (e.g., multiple sclerosis, rheumatoid arthritis, systemic lupus, Sjögren’s syndrome) are more prevalent in women, we hypothesize that autoimmunity may play a central role in the development of LC and other PAIS. Although the root causes of PAIS remain unclear, a wealth of data supports an autoimmune etiology of PAIS^3,16–27^. For example, AABs have been found in ME/CFS^28–30^ and LC^3,16–25^, some of which target GPCRs and GABA receptors involved in pathways relevant to neurological symptoms.

Infections naturally trigger the production of antibodies, which are essential in clearing pathogens. In addition, AABs often arise during infection due to bystander activation or molecular mimicry ^31–36^. Diverse and functional AABs are found during the acute phase of SARS-CoV-2 infection^16,37^. Typically, after infection is cleared, AAB levels subside as the immune system returns to homeostasis ^24^. However, four large-scale independent retrospective studies of medical records from millions of patients with COVID-19 reported a 20-40% increased risk of multiple new-onset autoimmune diseases after SARS-CoV-2 infection^38–41^. Therefore, we postulate that persisting AABs may cause symptoms for some patients with LC even though these patients may not meet classical criteria to be diagnosed with known autoimmune diseases.

The heterogeneity and current lack of understanding of the underlying mechanisms of LC pose significant challenges for the development of diagnostics and treatments. Identifying biomarkers that predict AAB-driven LC would have an immediate impact by enabling the matching of these patients to existing immunotherapies. Here, we examined the contributions of autoantibodies (AAB) to the development of neurological symptoms in patients with LC. Leveraging our Mount Sinai Yale Long COVID (MY-LC) cohort^42–45^, and an additional validation cohort (MY-LC II), we examined the autoreactivity of purified IgG against various human and mouse tissues. We identified autoantigens using a >21,000 human protein array and an antibody pull-down of autoantigens followed by mass spectrometry. Importantly, we demonstrated that passive transfer of these purified IgG samples from LC patients to healthy C57BL/6 mice recapitulated phenotypes of neurological symptoms of the donors.

## Results

### IgG targeting the CNS and PNS correlates with neurological symptoms in LC

From our Mount Sinai-Yale LC (MY-LC) cohort^42^, we analyzed 82 Long COVID patients, 35 healthy controls, and 30 convalescent controls (total 147 individuals). This group is predominantly comprised of women (73.3%) with an average age of 44.8 years, with no statistical differences between genders in cases and controls (Chi-squared test = 1.2117; p-values = 0.5456). Self-reported racial composition of the cohort was as follows. By race: 71.4% white, 6.1% Asian, 5.4% Black or African American, and 10.8% other or multiracial. By ethnicity, 17.6% identified as Hispanic or Latino. Most acute infections (80%) in the LC group occurred between the epidemiological weeks 7-18 of 2020, when the SARS-CoV-2 strain WA-1 drove most of the new cases. The sample collection took place on average 416 days after the acute symptoms. Only 13.8% of the patients had severe acute disease. These patients exhibited neurological symptoms, with 55.2% of them experiencing five or more symptoms. For this study, we considered the following neurological symptoms: brain fog, confusion, dizziness, loss of memory, sleep disturbance, alterations in mood, pins and needles, disorientation, hearing, fainting, and numbness in the face. The most common symptoms were fatigue (87.4%), brain fog (79.3%), headache (64.4%), loss of memory (62.1%), dizziness (65.5%), sleeping disturbance (58.6%), and confusion (56.3%). At the time of enrollment, these individuals reported not having an autoimmune diagnosis.

To evaluate if study participants had functional AAB profiles that could provoke their LC-associated neurological symptoms, we purified total IgG from plasma and performed immunofluorescence analyses using healthy human tissues (**Fig. 1A**). Due to sample limitations, we performed the immunofluorescence analysis on available samples consisting of 33 healthy controls, 28 convalescent controls, and 46 Long COVID individuals, with the indicated sample coverage (**Supp. Fig. 1A, Supp. Fig. 1A and Supp. Fig. 3A**). We observed that purified total IgG from a subset of LC participants showed increased reactivity against human locus coeruleus (**Fig. 1B**), human thalamus (**Fig. 1C**), mouse sciatic nerve (**Fig. 1D**), and mouse meninges (**Fig. 1E**). Few participants also displayed positivity against human dorsolateral prefrontal cortex (**Supp. Fig. 1B**). In general, the staining presented a diffuse pattern, with few cells demonstrating distinct labeling (**Supp. Fig. 2 and Supp. Fig. 1C**). On average 93.94% of healthy and 85.71% of convalescent controls tested positive for at least one neurological tissue. At the same time, long COVID patients exhibited reactivity to an average of 3.6 human neurologic tissues per individual, compared to healthy control (2.4 tissues) and convalescent control (2.5 tissues) (**Supp. Fig. 1A**). We observed that on average 52.17% of the Long COVID individuals were positive to at least one human peripheral tissue, with a mean of 3.41 tissues per individual (**Supp. Fig. 3A)**. Individuals with Long COVID showed diverse and increased positivity to several human peripheral tissues, including the adrenal gland, heart muscle, parathyroid, and thyroid tissues (**Supp. Fig. 3B**). A representative staining pattern for specific tissues are depicted in **Supp. Fig. 2 and Supp. Fig. 4**. The number of positive tissues per individual with Long COVID is described in **Supp. Fig. 3A**. Thus, Long COVID patients have a diverse and increased number of autoantibodies against human peripheral and CNS tissues. Receiver operating characteristic (ROC) analysis was performed to evaluate the ability of immunofluorescence (IF) signal intensity to discriminate between clinical groups. The area under the curve (AUC) was used as a summary measure of classification performance. ROC analysis demonstrated good discriminatory capacity between LC and controls for locus coeruleus and mouse meninges, supporting the robustness of the IF-based findings (**Fig. 1B and Fig. 1E**). ROC analysis for all tissues is shown in **Supp. Fig. 1 and Supp. Fig. 3**. To account for age and sex as a confounder in our analysis, we performed a linear regression analysis adjusting for age and sex (**Supp. Fig. 5I**). Notably, only mouse meninges remained significant despite several tissues showing an overall increase in autoreactivity in the LC group. We stained meninges from both female and male mice to compare sex-related differences in staining patterns in a sample classified as positive but observed no differences (**Supp. Fig. 1D**).

**Figure 1.**
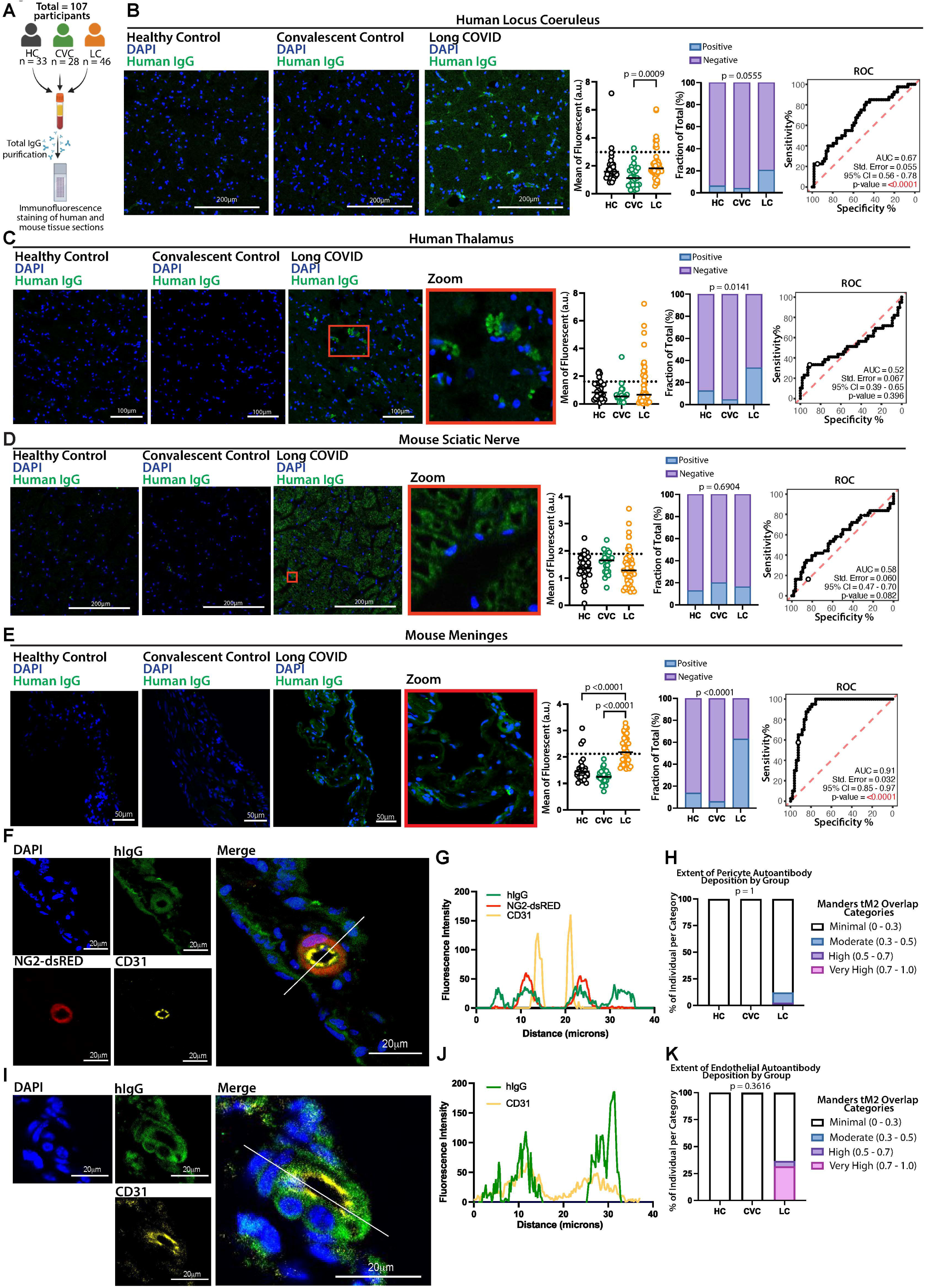
Purified IgG from participants with Long COVID reacts with CNS and PNS tissues. Confocal microscopy showing human and mouse tissues immunostained with human total IgG (green) purified from Long COVID, healthy or convalescent controls, as indicated, and nuclear DNA stain (DAPI, blue). **A.** Experiment schematic. **B.** Representative images of human locus coeruleus immunostaining, mean of fluorescence intensity, percent of positivity and Receiver operating characteristic (ROC) analysis. **C.** Representative images of human thalamus immunostaining, mean of fluorescence, percent of positivity, and ROC analysis. **D.** Representative images of mouse sciatic nerve immunostaining, mean of fluorescence, percent of positivity, and ROC analysis. **E.** Representative images of mouse meninges immunostaining, mean of fluorescence, percent of positivity, and ROC analysis. Scale bar described in the image. Insets indicate a higher magnification of the region indicated (red rectangle). Each dot in the figure represents the value obtained from an individual participant. Data are presented as the mean. Significant p-values are described in the image, as determined by the Kruskal-Wallis test followed by Dunn’s post-hoc multiple comparisons test. **F.** Confocal microscopy showing meninges from NG2-dsRED mouse stained with total IgG from Long COVID participant (green) and CD31 (yellow), NG2-dsRED stains in red, nuclear DNA stain in blue (Dapi). **G.** Line scan analysis showing fluorescent intensity for hIgG, CD31, and NG2-dsRED. The white line indicates the scan analysis in the graph. **H.** Manders tM2 co-localization analysis for pericytes. Scale bar described in the image. **I.** Confocal microscopy showing meninges from NG2-dsRED mouse stained with total IgG from Long COVID participant (green) and CD31 (yellow), nuclear DNA stain in blue (Dapi). **J.** Line scan analysis showing fluorescent intensity for hIgG, and CD31. The white line indicates the scan analysis in the graph. **K.** Manders tM2 co-localization analysis for endothelial cells. Scale bar described in the image.

**Figure 2.**
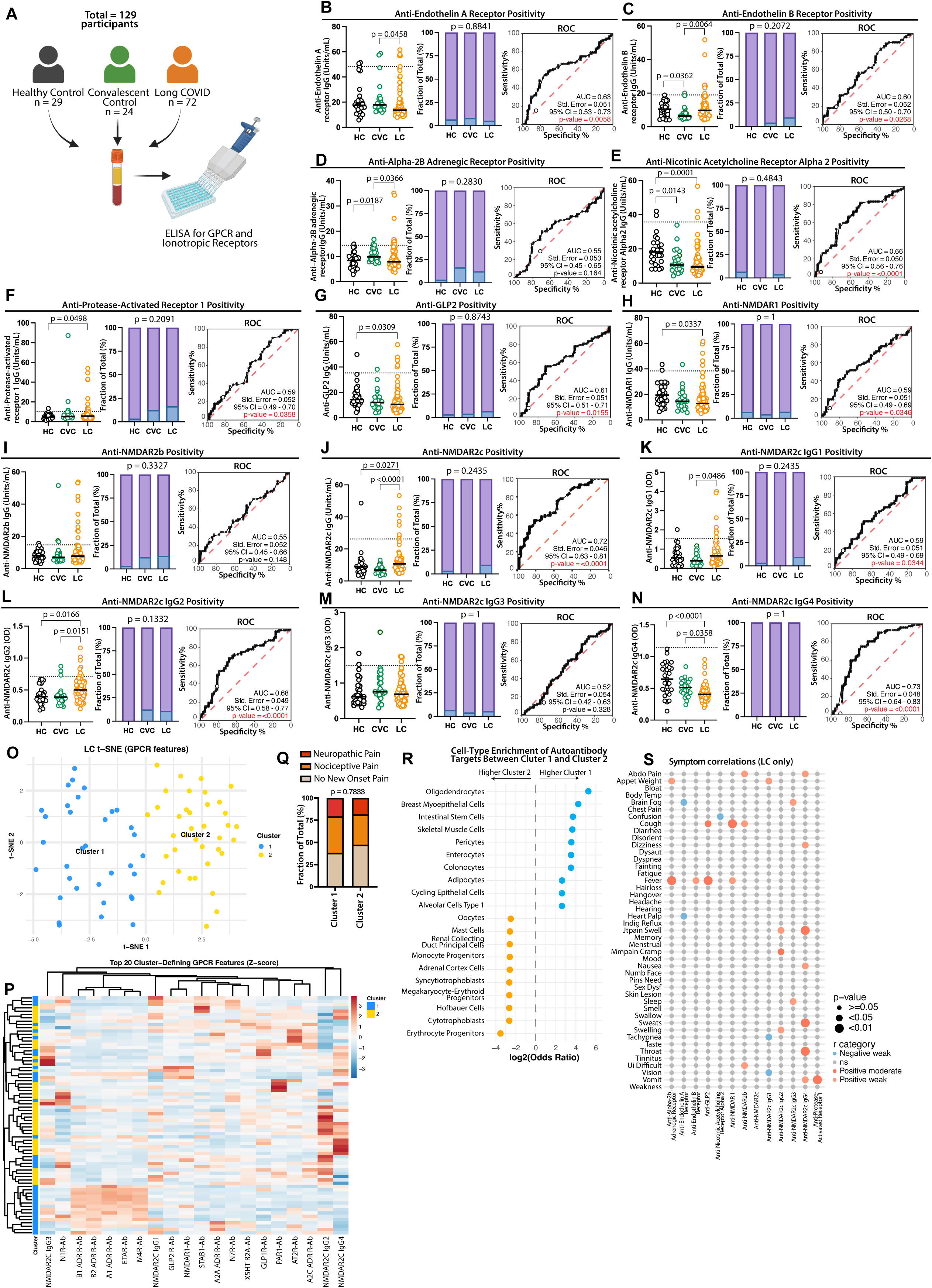
Elevated autoantibodies against GPCR and ionotropic receptors in LC. **A.** Number of samples per group that were used to perform ELISA for anti-GPCRs. **B-N.** ELISA for anti-GPCRs showing increased antibody levels in LC compared with controls. Plots shows mean of fluorescence intensity, percent of positivity and Receiver operating characteristic (ROC) analysis. Each dot represents one individual. P-values are described in the figure and were determined by the Kruskal-Wallis test followed by post hoc Dunn’s multiple comparisons test. p-values were adjusted for multiple comparisons using the Benjamini-Hochberg method, FDR method. **K-N.** IgG subclass distribution of anti-NMDAR2C antibodies, revealing increased IgG1 and IgG2 and decreased IgG4 in LC individuals. Each dot represents one individual. Dashed lines indicate positivity thresholds defined as the mean of HC ± 2 SD. P-values are described in the figure and were determined by the Kruskal-Wallis test followed by post hoc Dunn’s multiple comparisons test. **O.** T-distributed stochastic neighbor embedding (t-SNE) using ELISA IgG reactivity values showing two clusters between the long COVID participants. **P.** Heatmap showing the reactivity against the top cluster-defining features. **Q.** Frequency distribution of new onset pain between LC individuals divided by cluster. **R.** Cell-type enrichment of autoantibody target between Clusters. **S.** Point-Biserial symptom correlation for anti-GPCRs. p-values were adjusted for multiple comparisons using the Benjamini-Hochberg method, FDR method. Significant associations were visualized in a dot plot and color-coded by r category.

**Figure 3.**
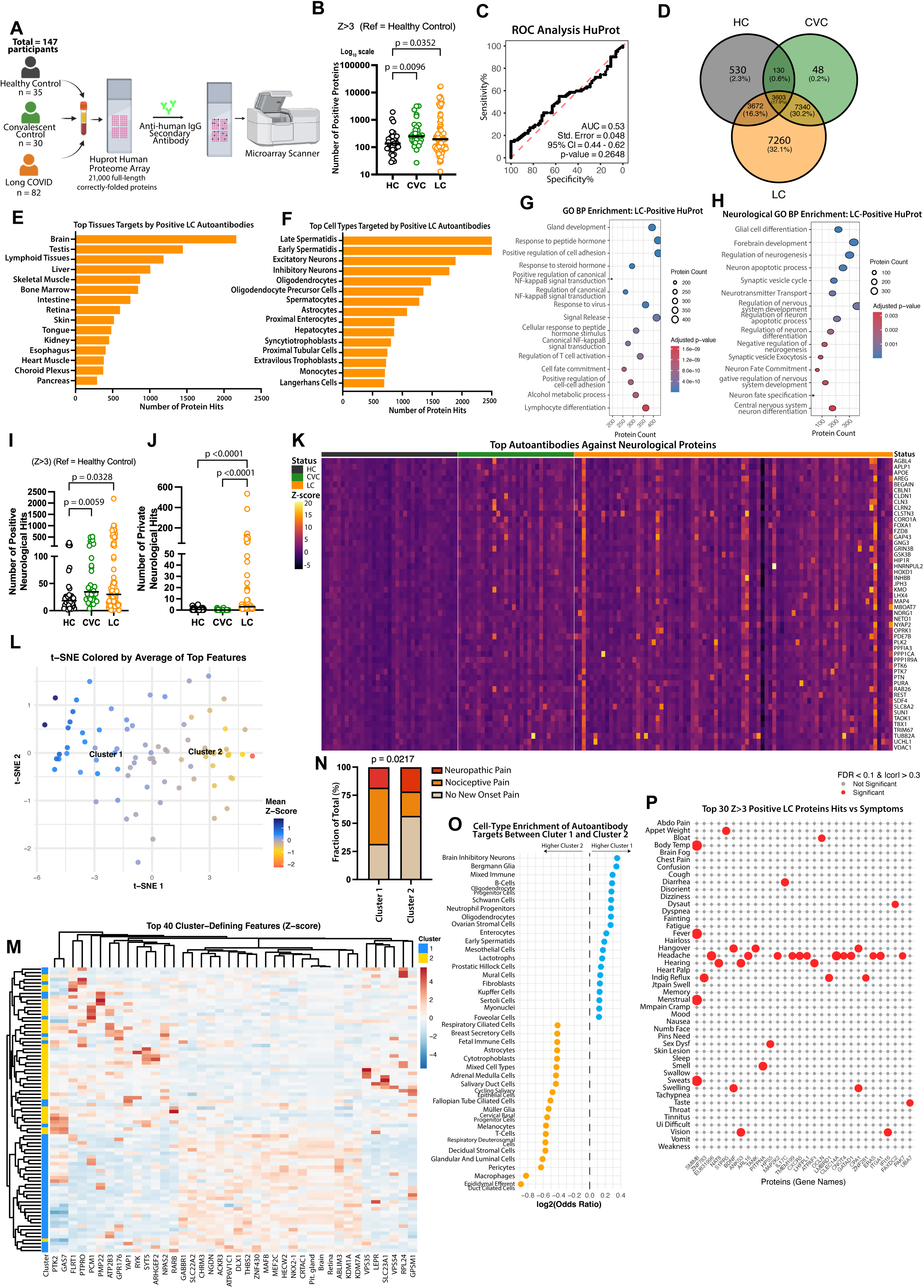
Elevated autoantibody reactivity in individuals with Long COVID, including autoreactivity of CNS tissues. **A.** Plasma from participants was incubated with HuProt microarrays and probed with a secondary antibody against human IgG; positive targets were identified using a microarray scanner. **B.** Number of auto-reactivities for each person within each group starting at a HuProt intensity threshold of Z-score > 3. P-values are described in the figure and were determined by the Kruskal-Wallis test followed by post hoc Dunn’s multiple comparisons test. **C.** ROC analysis of the number of positive hits for all proteins in the HuProt dataset. **D.** Venn diagram showing the number of shared and private positive hits between groups. **E.** Bar plot showing the number of positive autoreactivities per tissue. **F.** Bar plot showing the number of positive reactivity per cell type. **G.** Pathway enrichment analysis on the positive hits identified individuals with Long COVID. **H.** Neurological pathway enrichment analysis on the positive hits identified in individuals with Long COVID. **I.** Number of positive neurological auto-reactivities for each person with a threshold of Z-score > 3, p-values are described in the figure and were determined by the Kruskal-Wallis test followed by post hoc Dunn’s test. **J.** The number of private neurological auto-reactivities for each person with a threshold of Z-score > 3, p-values are described in the figure and were determined by the Kruskal-Wallis test followed by post hoc Dunn’s test. **K**. Heatmap showing the reactivity against the top 50 neurological hits for each group. **L.** Dot plot showing pairwise comparisons of tissue-level reactivity between groups. Circle size represents the statistical significance (−log₁₀ adjusted *p*-value) and color indicates the direction and magnitude of difference in mean tissue reactivity determined by Wilcoxon rank-sum tests, followed by Benjamini-Hochberg correction for multiple testing (FDR). **L.** T-distributed stochastic neighbor embedding (t-SNE) using z-score normalized IgG reactivity values showing two clusters between the long COVID participants. **M.** Heatmap showing the reactivity against the top 40 cluster-defining features. **N**. Frequency distribution of new onset pain between LC individuals divided by cluster. **O.** Cell-type enrichment of autoantibody target between Clusters. **P.** Dot plot showing point biserial correlation of top positive hits for Huprot with symptoms, p-values were adjusted for multiple comparisons using the Benjamini-Hochberg method, FDR method. Significant associations were visualized in a dot plot, with effect sizes represented as – log₁₀(p-value) and color-coded by significance.

**Figure 4.**
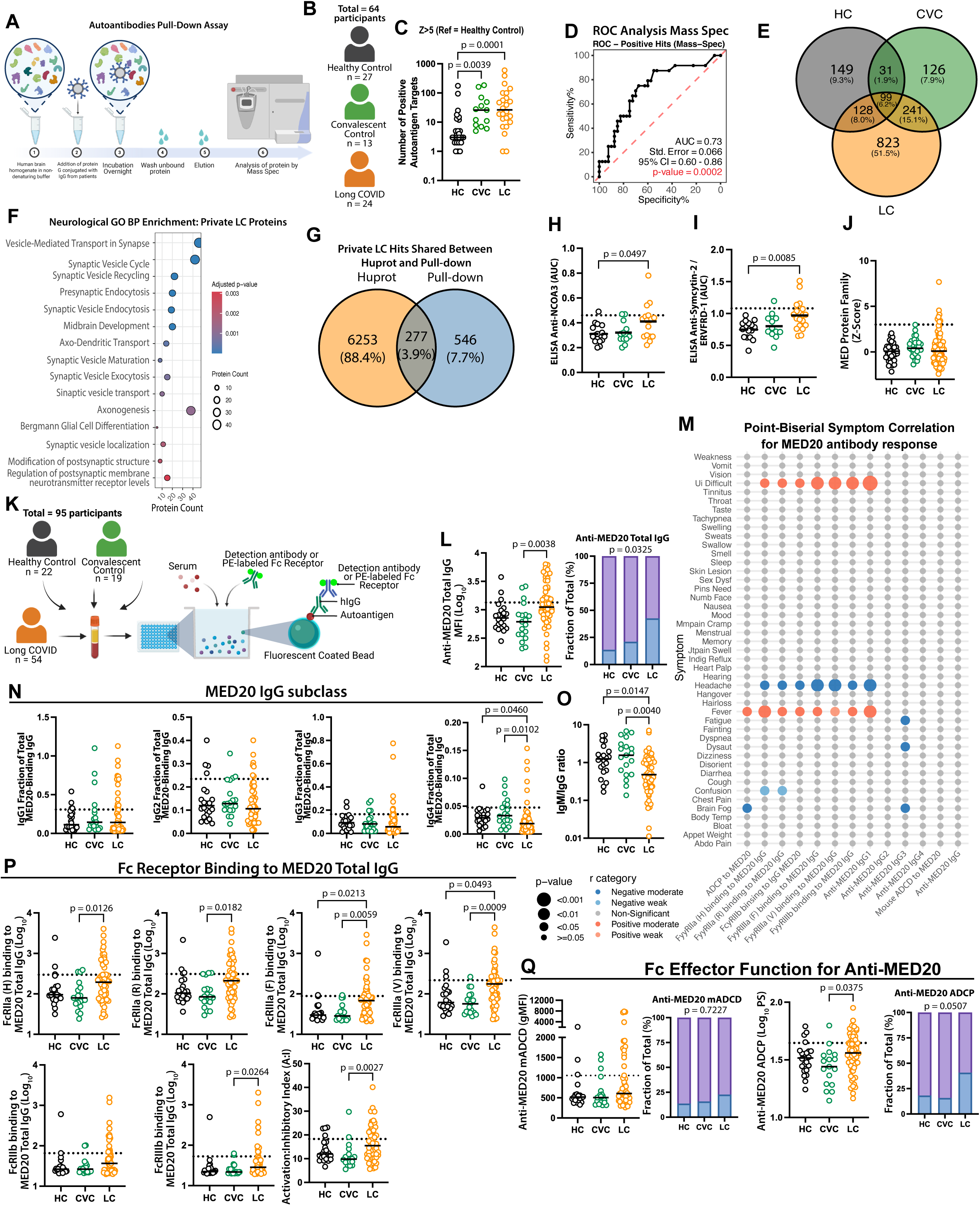
Autoantibodies validation and association with symptoms. A-B. Number of samples per group that were used to perform a pull-down of autoantigens using human brain homogenate, followed by mass spec. **C.** Number of auto-reactivities for each person within each group, starting at an intensity threshold of Z-score > 5. P-values are described in the figure and were determined by the Kruskal-Wallis test followed by post hoc Dunn’s test. **D.** ROC analysis of the number of positive hits identified by mass spectrometry (MS). **E.** Venn diagram showing the number of shared and private positive hits between groups. **F.** Pathway enrichment analysis on the private neurological hits identified in individuals with Long COVID. **G.** Venn diagram showing the number of shared and private positive hits between Huprot and pull-down, followed by mass spec. **H-I**. ELISA validation of autoantigens showing Area Under Curve (AUC) analysis for top hit targets identified by Huprot (NCOA3 and Syncyting-2). Each dot in the figure represents the value obtained from an individual person. **J.** HuProt Z-score for proteins that belong to the Mediator (MED) Family. **K.** Number of samples per group that were used to perform the effector function analysis for anti-MED20. **L.** Luminex assay measuring MED20-specific total IgG. **M.** Point-Biserial symptom correlation for anti-MED20. p-values were adjusted for multiple comparisons using the Benjamini-Hochberg method, FDR method. Significant associations were visualized in a dot plot and color-coded by r category**. N.** Luminex assay measuring MED20-specific total IgG subclasses. **O.** Anti-MED-20 IgM/IgG ratio. **P.** Binding of specific MED20 IgG to individual Fc Receptors (FcγRIIa (H); FcγRIIa (R) ; FcγRIIIa (F); FcγRIIIa (V); FcγRIIb; FcγRIIIb) and Activation:Inhibitory (A:I) index calculated as (FcγRIIA-H131 + FcγRIIA-R131 + FcγRIIIA-V158 + FcγRIIIA-F158 + FcγRIIIb) / FcγRIIb. **Q.** Fc Effector function analysis for anti-MED20 showing geometric Mean Fluorescence Intensity (gMFI) for mADCD and log_10_ phagocytic score (PS) for ADCP. Each dot represents one individual. Dashed lines indicate positivity thresholds defined as the mean of HC ± 2 SD. P-values are described in the figure and were determined by the Kruskal-Wallis test followed by post hoc Dunn’s multiple comparisons test.

**Figure 5.**
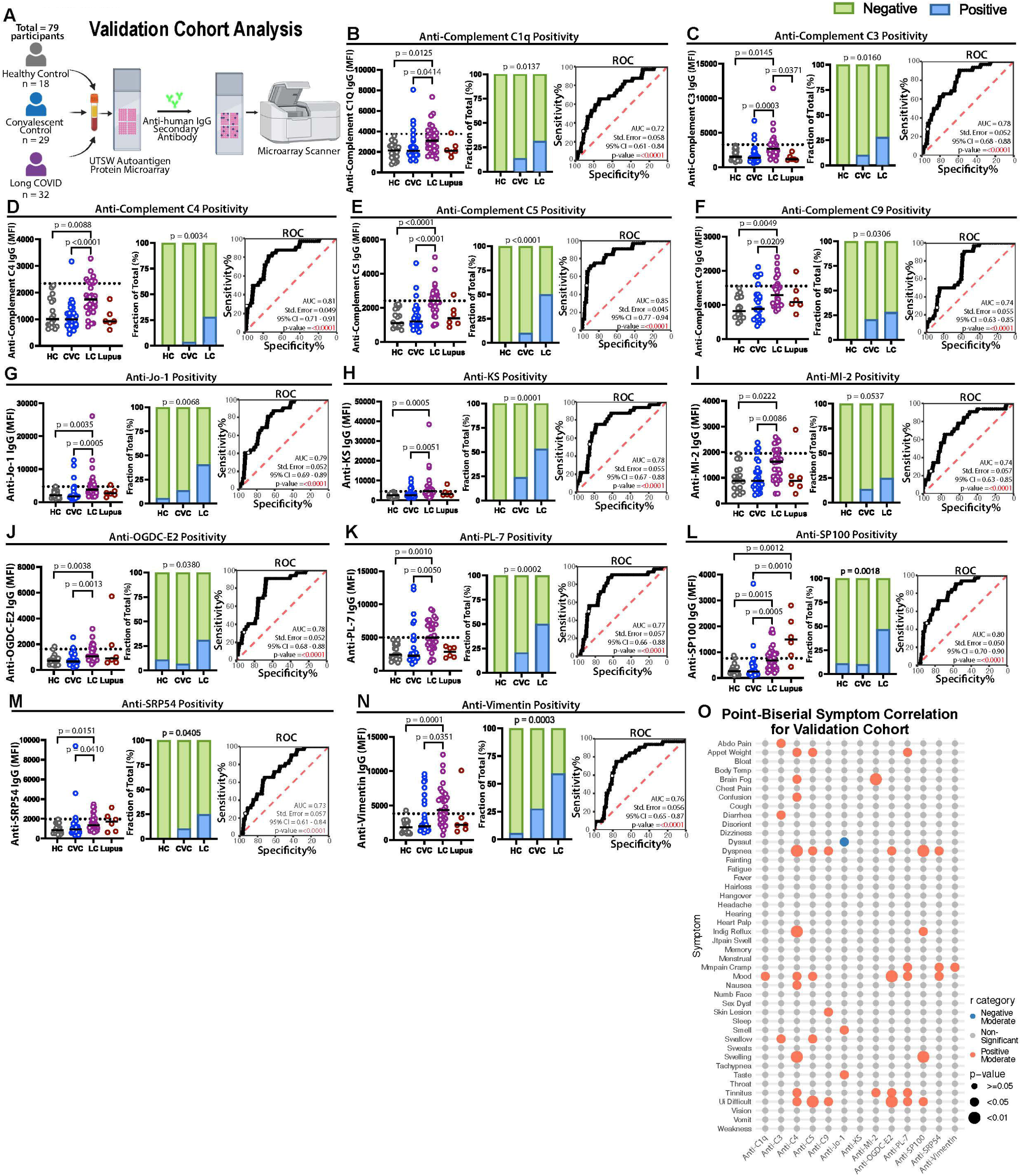
Increased autoantibodies in LC Individuals in an independent validation cohort. **A.** Protein microarray analysis performed in an independent validation cohort, showing increased antibody reactivities in LC compared with controls. **B-N.** Plots shows mean of fluorescence intensity, percent of positivity and Receiver operating characteristic (ROC) analysis. Each dot represents one individual. Dashed lines indicate positivity thresholds defined as the mean of HC ± 2SD. P-values are shown in the figure and were determined using the Kruskal–Wallis test followed by Dunn’s post hoc multiple comparisons test. p-values were adjusted for multiple comparisons using the Benjamini-Hochberg method, FDR method. **O.** Point-Biserial symptom correlation for significant autoantibodies identified in the validation cohort. p-values were adjusted for multiple comparisons using the Benjamini-Hochberg method, FDR method. Significant associations were visualized in a dot plot and color-coded by r category.

Next, we analyzed whether autoreactive antibodies targeting central and peripheral nervous system tissues correlate with LC symptoms. Specifically, we assessed whether patients reporting a given symptom were more likely to exhibit positive staining in a specific tissue, and whether the mean of fluorescence in a particular tissue was associated with the presence or absence of specific symptoms. We observed an essential heterogeneity of symptoms and autoantibody response in the LC group. Due to this diversity, only a few tissues and symptoms reached statistical significance after multiple comparison correction. Nevertheless, the overall increase in autoreactivity in the LC group is visible in the effect size of the mean of fluorescence or in the frequency of individuals with positive symptoms who had antibodies against specific tissues (**Supp. Fig. 5A-H**). Notably, antibodies against human locus coeruleus were significantly associated with loss of taste and smell (**Supp. Fig. 5A**), as well as nausea, skin lesions, joint pain and joint swelling (**Supp. Fig. 5B**). Despite not being significant, 75% of the patients with antibodies against mouse meninges reported headache (**Supp. Fig. 5C-D**).

To evaluate the specificity of the cross-reactive antibodies against the mouse meninges, tissue was co-stained using antibodies to NG2-dsRED (a marker for pericyte) and CD31 (endothelial cell marker). We observed that the staining with IgG from 22 LC participants co-localized with pericytes and endothelial cells (**Fig. 1F-K**).

### Identification of autoantibody targets

To identify autoantibody (AAB) targets against G protein-coupled receptors (GPCRs) and ionotropic receptors, we tested plasma samples from 29 healthy controls, 24 convalescent controls, and 72 individuals with LC using ELISA (**Fig. 2A-N and Supp. Fig. 6A-N**). We observed that patients with LC have elevated IgG levels against NMDAR2C (**Fig. 2J**). Anti-NMDAR2C reactivity showed discriminatory potential, with an AUC of 0.72 in ROC analysis, suggesting that elevated antibody levels moderately distinguish affected individuals from controls (**Fig. 2J**) and showed higher inflammatory subclasses IgG1 and IgG2 in LC (**Fig. 2K-N**).

**Figure 6.**
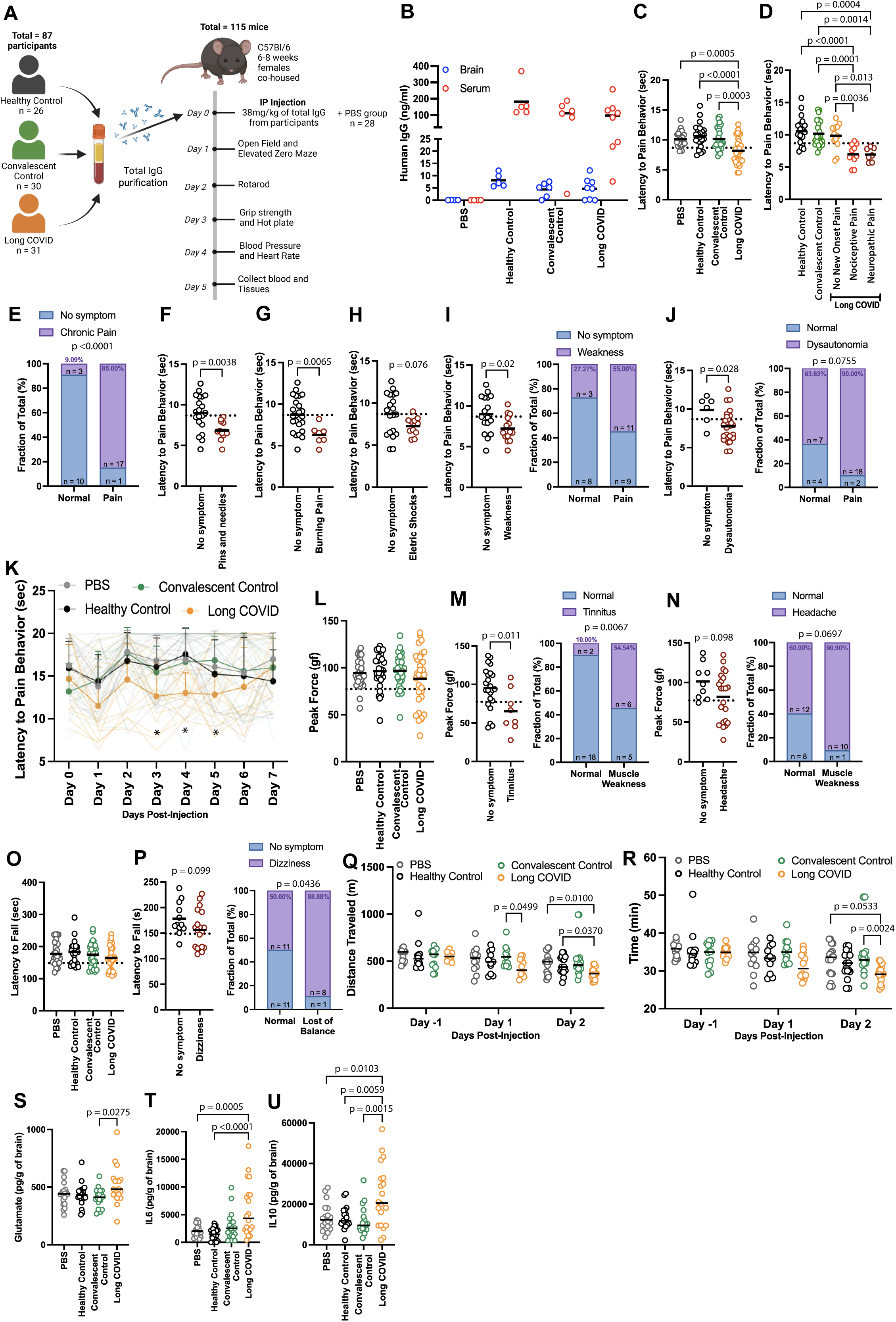
IgG from individuals with LC induces symptoms in mice. **A.** A dose of 38.4 mg/kg of total IgG purified from healthy, convalescent controls and Long COVID participants was administered to 6-8 weeks-old C57BL/6 female mice by intraperitoneal (IP) injection, and the mice were followed up for 5 days. **B.** Quantification of human IgG by ELISA in brain homogenate and serum at day 5 post-injection. **C.** Hot plate test performed on day 3 post-injection. **D.** Hot plate test divided by the status of chronic pain displayed by the participants. **E.** Frequency of mice with pain at the hot plate test and the patients with diagnosed chronic pain. **F-H.** The hot plate test was divided by how the participants described their pain sensation. **I**. The hot plate test was divided by participants who displayed weakness. **J.** Hot plate test divided by participants who displayed dysautonomia. **K.** Longitudinal hot plate test. **L.** Grip strength test performed on day 3 post-injection. **M.** Grip strength test was divided by participants who displayed tinnitus. **N.** Grip strength test divided by participants who displayed a headache. **O.** Rotarod test performed on day 2 post-injection. **P.** Rotarod test was divided by participants who displayed dizziness. **Q.** Distance traveled in the treadmill test. **R.** Total time to reach fatigue in the treadmill test. **S.** Glutamate levels in brain homogenate 3 days post-injection of IgG. **T.** IL6 levels in brain homogenate 3 days post-injection. **U.** IL10 levels in brain homogenate 3 days post-injection. Each dot in the figure represents the value obtained from an individual mouse. Each mouse received antibodies isolated from a single human participant. Data are presented as the mean. Significant p values are described in the image, as determined by a T-test for two-group comparisons or one-way ANOVA followed by Tukey’s post-hoc multiple comparisons test.

To evaluate whether LC individuals could be stratified based on their AAB profile, we performed t-distributed stochastic neighbor embedding (t-SNE) using ELISA measurements targeting GPCR and ionotropic receptors (**Fig. 2O**). This dimensionality reduction analysis identified two clusters of patients with distinct AAB signature. Cluster 1 displayed a relatively homogeneous pattern of reactivity, characterized by consistent positivity against Beta 1 and Beta 2 adrenergic receptors, endothelin A receptor and muscarinic acetylcholine receptor M4 (**Fig. 2P**). In contrast, Cluster 2 exhibited a more heterogeneous AAB profile with variable reactivity across other targets (**Fig. 2P**).

Despite these differences in AAB profile, the clusters were not significantly enriched for chronic pain (**Fig. 2Q**) or any other symptoms (data not shown). Notably, expression enrichment analysis of the targets revealed that targets preferentially recognized in Cluster 1 are enriched in oligodendrocytes (**Fig. 2R**), suggesting potential cell-type specificity of the AAB response. Importantly, although cluster-level symptom enrichment was not observed, analysis at the individual AAB level revealed significant positive correlations between specific receptor-directed antibodies and multiple clinical symptoms (**Fig. 2S)**.

We also screened plasma samples from 35 healthy controls, 30 convalescent controls, and 82 individuals with Long COVID using the HuProt™ microarray, which includes over 21,000 full length human proteins (**Fig. 3A**). Applying a Z-score threshold of 3 to define positivity, we quantified the number of positive hits per sample and found that individuals with Long COVID and convalescent controls exhibited a higher frequency of autoreactive antibodies compared to the healthy control group (**Fig. 3B**). ROC analysis for the HuProt protein microarray yielded an AUC value of 0.53 for all positive hits (**Fig. 3C**), reflecting broad autoreactivity in the CVC group (**Fig. 3B**) and poor discrimination between the CVC and LC groups. Notably, we identified 7,260 private hits unique to the Long COVID group (**Fig. 3D**). Among the positive hits, brain tissue was the top target of these AAB, followed by testis and lymphoid tissues (**Fig. 3E**). Among the affected cell types, we observed reactivity against neurons, oligodendrocytes, and astrocytes, as well as spermatocytes (**Fig. 3F**). Given the predominance of females in our cohort, the relevance of the presence of AAB targeting male-specific tissues remain unclear.

To identify the biological pathways affected by AAB, we performed pathway enrichment analysis on the positive and private hits found in individuals with Long COVID (**Fig. 3G-H**). This analysis revealed significant enrichment in pathways related to hormone signaling, cell adhesion, synaptic function, neurogenesis and regulation of neuronal apoptosis. Given that brain tissue and cells of the central nervous system (CNS) were identified as the top targets of these AAB, we focused further on the neurological implications. We observed that individuals with Long COVID had an increased frequency of positive and private autoantibodies targeting proteins associated with neurological function (**Fig. 3I, J**). However, when examining the top 50 neurological targets, we found significant heterogeneity in autoantibody positivity among the Long COVID cohort (**Fig. 3K**). Although neural tissues were the primary targets of these AAB, individuals with Long COVID exhibit increased reactivity to a broad range of tissues (**Supp. Fig. 7A**).

**Figure 7.**
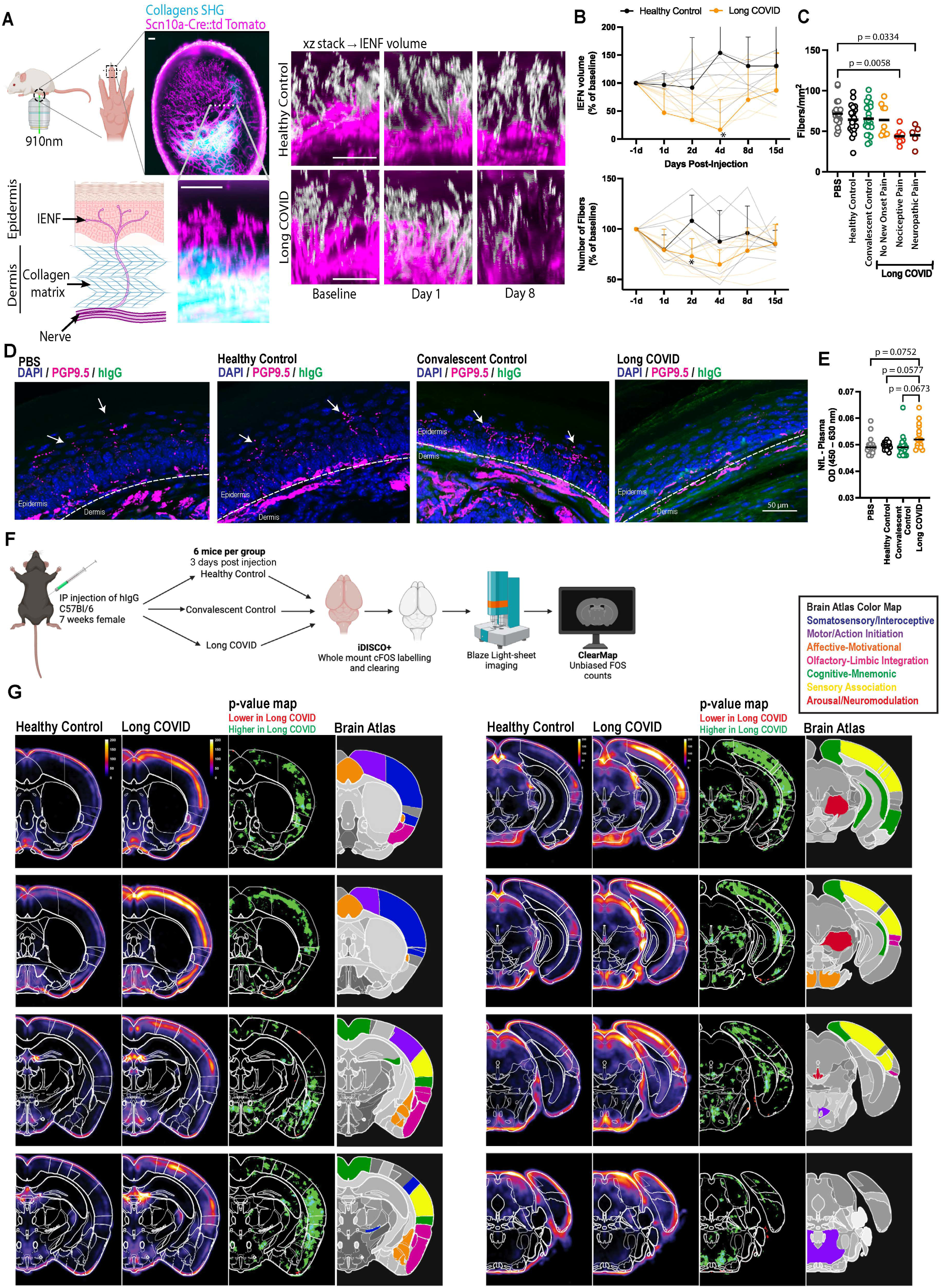
IgG from individuals with Long COVID induces injury of peripheral small nerves and brain-wide neuronal activity in mice. **A.** A dose of 38.4 mg/kg of total IgG purified from healthy controls and Long COVID participants was administered to 8-10 weeks-old C57BL/6 Scn10a-Cre:td Tomato female mice by intraperitoneal (IP) injection, which were followed up for 15 days. **B.** Longitudinal **t**wo-photon imaging of reporter mice reveals collagen fibers by second harmonic generation (SHG) as well as nociceptor axons and the perpendicular intraepidermal nerve fibers (IENF; white/magenta). Quantification of total IENF volume and number of fibers crossing the dermis boundary after passive transfer. Scale bars 50μm. Each dot in the figure represents the value obtained from an individual mouse. Each mouse received antibodies isolated from a single human participant. Data are presented as the mean. Significant p values are described in the image, determined by unpaired *t*-test for two-group comparisons or by two-way ANOVA with Šídák’s post-hoc correction for multiple-group comparisons. **C.** A dose of 38.4 mg/kg of total IgG purified from healthy, convalescent controls and Long COVID participants was administered to 6-8 weeks-old C57BL/6 female mice by intraperitoneal (IP) injection, and the mice were followed up for 5 days. Confocal microscopy showing IENF counts in the paws of those mice, divided by the status of chronic pain displayed by the participants. **D.** Representative images of IENF staining in the paws, PGP9.5 (magenta) hIgG (green), and nuclear DNA stain (DAPI, blue). Significant p values are described in the image, as determined by the T-test for two-group comparison or one-way ANOVA followed by Tukey’s post-hoc multiple comparisons test. **E.** ELISA for neurofilament light chain (NfL) in the plasma 5 days post-injection. **F.** Schematic for whole brain activity mapping for mice after passive transfer of hIgG. **G.** A dose of 38.4 mg/kg of total IgG purified from healthy, convalescent controls and Long COVID participants was administered to 7-week-old C57BL/6 female mice by intraperitoneal (IP) injection, which were followed up for 3 days. Fos mapping results for brain areas, first two columns showing heatmap with Fos intensity, third column represents p-value maps by Kruskal-Wallis test, followed by Dunn’s post-hoc multiple comparisons test, last column represents Allen Brain annotation. The heatmaps are the average of all mice in each group (n = 6 per group).

Considering the heterogeneity in autoantibody reactivity, we aimed to determine whether a subset of patients with a higher autoantibody burden drove this variability. To assess this, we analyzed the number of private hits per patient and identified a distinct subgroup of patients with significantly higher autoantibodies (**Supp. Fig. 7G**). To further investigate the clusters of neural autoantibody profiles in Long COVID patients, we performed t-SNE using z-score normalized IgG reactivity data from the HuProt protein array (mean silhouette score = 0.55; PERMANOVA R² = 0.69, p = 0.001; **Fig. 3L**). This analysis allowed us to identify clusters of patients with the strongest overall reactivity to neural proteins (**Fig. 3M**). Cluster 2 demonstrated greater overall positivity toward proteins enriched in glandular and epithelial cell types (**Fig. 3O**). In contrast, Cluster 1 preferentially targeted proteins enriched in neurological cell populations (**Fig. 3O**). Clinically, Cluster 1 exhibited a higher proportion of patients reporting new-onset pain (**Fig. 3N**). Although this cluster also showed enrichment for neurological symptoms, no other symptom comparisons reached statistical significance (data not shown). We observed 66.7% agreement in cluster assignments between ELISA against GPCRs and ionotropic receptors and HuProt (**Supp. Fig. 6O**).

We next analyzed the correlation between symptoms and the positive hits identified by HuProt. We observed significant correlations between private and positive hits, and several symptoms (**Fig. 3P**). Notably, headache was significantly correlated with multiple autoantibody targets (**Fig. 3P**).

We also assessed the presence of common autoantigens using HuProt in the Long COVID group, comparing our findings with those from patients with systemic lupus erythematosus (SLE). We observed that a subset of individuals with Long COVID had a similar range of positive hits as patients with SLE (**Supp. Fig. 7C**). However, while SLE patients exhibited higher z-scores for anti-TPO autoantibodies, very few samples were considered positive for anti-TPO (**Supp. Fig. 7D**). A similar trend was seen for SS-A 60kD (Ro60), Scl-70, and Lupus LA. SLE patients showed increased antibodies against the Sm/RNP complex and PTGS2, a finding absent in the Long COVID cohort. No positive hits were observed in lupus or the Long COVID groups against Sm, JO-1, or dsDNA (**Supp. Fig. 7B**). These findings suggest that a small subset of Long COVID patients exhibit increased autoantibody levels against common autoantigens comparable to SLE patients, but most fall below the threshold of positivity. We also performed antinuclear antibody (ANA) testing across all groups and observed a trend toward higher ANA index values in LC patients, suggesting increased autoreactivity to nuclear antigens (**Supp. Fig. 7C**). However, the proportion of ANA-positive individuals above the clinical threshold (ANA Index 1) did not differ significantly between groups.

Given the increased diversity of targets identified by HuProt, we aimed to narrow down the list of potential pathogenic targets by performing a second screening. For this, we conducted an antibody pull-down assay followed by mass spectrometry using human brain homogenate and antibodies from 27 healthy controls, 13 convalescent controls, and 24 Long COVID patients (**Fig. 4A-B**). We applied a similar methodology as for HuProt analysis, but with a higher threshold (Z-score > 5) to identify positive and private hits, given the greater variability of autoantibodies identified in our cohort. Consistent with our previous findings, the Long COVID group exhibited increased autoreactivity (**Fig. 4C**). ROC analysis for the pull-down assay followed by mass spectrometry yielded an AUC value of 0.73 for all positive hits (**Fig. 4D**), reflecting a good discrimination power between controls and LC groups. LC individuals have 823 private hits (**Fig. 4E**). Notably, a subset of these individuals showed a higher burden of autoreactivity against neural proteins (**Supp. Fig. 7H)**, with most of those hits targeting synapse function pathways (**Fig. 4F**).

We next evaluated cross-platform concordance in identifying patients with a high autoantibody burden using the Jaccard index and assay-specific thresholds (ELISA > 2 positive antibodies; HuProt > 100 positive proteins; immunofluorescence (IF) > 2 positive tissues; mass spectrometry > 20 positive antibodies). As expected for platforms that interrogate distinct immunological features, agreement across assays ranged from modest to moderate. Specifically, agreement ratios were 0.35 for ELISA–HuProt, 0.64 for HuProt–IF, 0.32 for ELISA–IF, 0.54 for IF–mass spectrometry, 0.52 for HuProt–mass spectrometry, and 0.46 for ELISA–mass spectrometry (**Supp. Fig. 7I**). In general, 39 patients were classified as high burden in at least two assays, 15 in three assays, and 6 in all four assays, demonstrating reproducibility across platforms and biological coherence of the antibody signal (**Supp. Fig. 7J**).

To assess whether these overlaps exceeded chance expectation given assay-specific positivity rates, we performed permutation-based significance testing for each assay pair. High-burden labels were randomized 1,000 times within the overlapping sample set while preserving the marginal positivity rates of each assay. Only the ELISA–HuProt comparison showed statistically significant overlap, likely reflecting its larger shared sample size (n = 72; **Supp. Fig. 7I**). No other assay pair exceeded chance expectations, consistent with the fact that IF, mass spectrometry, and GPCR-focused ELISA measure distinct aspects of autoantibody binding (e.g., tissue binding, brain-antigen pull-down targets, and GPCR-specific reactivity) and were available for fewer participants.

To complement this threshold-based analysis, we also compared assays using their continuous hit counts. For each assay pair, we restricted the analysis to participants tested on both platforms and computed Spearman rank correlations to determine whether higher burden in one assay was associated with higher burden in another. This threshold-independent analysis provides an orthogonal assessment of cross-assay consistency (**Supp. Fig. 7K**). ELISA–HuProt again showed the strongest association (ρ = 0.453, p < 0.001), supporting concordance between these two high-throughput profiling methods. Additional significant correlations were observed for HuProt–mass spectrometry (ρ = 0.428, p = 0.03) and ELISA–mass spectrometry (ρ = 0.528, p = 0.03). In contrast, assay pairs involving IF showed weak correlations (ρ = −0.131 to 0.119, all p > 0.4), reflecting the fundamentally different biological aspects measured by tissue-based immunoreactivity and the other assays. Collectively, these analyses indicate that although each platform captures orthogonal aspects of autoantibody biology, convergent signals are detectable across independent assays, particularly between the two broad-spectrum profiling approaches (ELISA and HuProt). Together, these results demonstrate that LC-associated autoantibody reactivities have strong discriminatory power and are reproducible across multiple assay modalities, supporting their diagnostic and mechanistic relevance.

When comparing antibody pull-down assay followed by mass spectrometry with HuProt, we observed 277 shared positive hits (**Fig. 4G, Supp. Table 1**). To further validate these targets, we curated a list of 17 top hits that were validated by ELISA (**Supp. Fig. 7E**). In addition, we also added autoantigens that have been described as associated with small fiber neuropathy: heparin (as a substitute for TS-HDS)^46,47^, FGFR3^48^ , and Plexin D^49^. The participants identified by HuProt as positive for NCOA3 and Syncytin-2 also exhibited higher antibodies to these antigens by ELISA (**Fig. 4H-I**). No positivity for heparin, FGFR3, or Plexin D1 was detected. For the other antigens that were tested, similarly to what we observed in the HuProt data, we had diverse positivity across patients (**Supp. Fig. 7E**).

To evaluate the concordance of autoantibody detection between the HuProt and ELISA validation of curated targets, we conducted a Jaccard similarity analysis to compare the positive and negative samples across both methods. The results showed that most samples exhibited a concordance of over 70% (**Supp. Fig. 7F**). We also found strong correlations between the ELISA for GPCRs and HuProt datasets for NMDAR2C (r = 0.4; **Supp. Fig. 7L**) and NMDAR1 (R = 0.37; **Supp. Fig. 7M**), demonstrating cross-platform reproducibility of these reactivities.

In summary, we did not observe a striking quantitative difference in the number of autoantibodies between LC and CVC samples across the screenings we performed. Moreover, the substantial heterogeneity in autoantibody reactivity observed within the LC group presents a major challenge for identifying a single shared target with robust diagnostic utility or consistent associations with clinical symptoms. However, our data suggest that although autoreactivity may manifest following infection, the composition of these antibodies is notably different in LC (**Fig. 3D** and **Fig. 4E**). In order to explore the distinct nature of autoantibodies in LC patients compared to CVC individuals, we also explored the mechanistic basis of how specific autoantibody reactivity might lead to symptoms.

### Fc Effector Function Analysis of anti-MED20

Among our initial top candidates, we identified antibodies recognizing proteins of the Mediator (MED) family, which form a multiprotein complex essential for transcriptional regulation. To investigate the mechanistic basis linking autoantibody reactivity to long COVID symptoms, we characterized the Fc biophysical and functional properties of autoantigen-specific antibodies. We observed that a subset of LC patients had autoantibodies against proteins in the MED family, which form a multiprotein complex essential for transcriptional regulation (**Fig. 4J**). Within this family, we selected MED20 for further validation because of its consistent targeting by antibodies from LC patients. Using a Luminex assay, we analyzed 22 healthy controls, 19 convalescent controls, and 54 individuals with Long COVID (**Fig. 4K)**. We observed higher total IgG levels against MED20 in LC than in CVC (**Fig. 4L**). No significant differences were detected in MED20-specific IgG1, IgG2, or IgG3 levels between groups (**Fig. 4N),** however, IgG4 reactivity was decreased in LC samples (**Fig. 4N).** Because IgG4 exhibits relatively low Fc-mediated effector activity compared with IgG1 and IgG3, reduced IgG4 levels in LC individuals are consistent with enhanced overall Fc effector function^50^. Additionally, individuals with LC exhibited a reduced IgM/IgG ratio, consistent with enhanced class switching and/or altered humoral maturation dynamics in this cohort (**Fig. 4O**).

Antibodies targeting MED20 bound multiple Fcγ receptors (FcγRs) (**Fig. 4P**), suggesting potential for diverse effector functions. Prior work has established that the relative engagement of activating versus inhibitory FcγRs (e.g., FcγRI/IIa/IIIa vs. FcγRIIb) is a key determinant of antibody effector activity^51^. We quantified this ratio using an activation-to-inhibitory (A:I) index. We observed that the A:I index was significantly higher in LC than in CVC (**Fig. 4P)**, suggesting that autoantibodies, such as anti-MED20 IgG, may preferentially engage activating FcγRs, thereby enhancing inflammatory or cytotoxic functions.

To explore through which effector functions of IgG against MED20 could be pathogenic, we performed an antibody-dependent complement deposition (ADCD) assay and antibody-dependent cellular phagocytosis (ADCP) assay (**Fig. 4Q).** We observed increased ADCP with serum derived from LC individuals (**Fig. 4Q)**. Correlation analysis with clinical symptoms further revealed that MED20-specific IgG–mediated ADCP and Fc receptor binding positively associated with reported fever and urinary difficulty (**Fig. 4M**). A trend toward increased complement deposition was observed in LC, but statistical significance was not achieved, potentially due to limited sample size (**Fig. 4Q)**. Collectively, these results suggest that, while autoreactive IgG is present in CVC, the specificity and functionality of autoantibodies are uniquely pathological in patients with LC.

### Common Autoantigen Profiling in an Independent Validation Cohort

To ensure the robustness and generalizability of autoantibody associations identified in our cohort, we expanded our analysis to an independent validation cohort (18 HC, 29 CVC, and 32 LC) (**Fig. 5A** and **Supp. Table 2**). We assessed IgG levels using a human protein microarray containing 118 clinically relevant autoantigens^52^. This array included established neurological and systemic autoimmune targets. Several autoantibodies were significantly increased in LC patients, including antibodies against complement components C1q, C3, C4, C5 and C9, SP100, Jo-1, KS, MI 2, OGDC E2, PL7, SRP54, SP100, Vimentin (**Fig. 5B–N** and **Supp. Fig. 6P**). ROC analyses revealed good to excellent discrimination (AUC > 0.7) between the LC and control groups for these antigens (**Fig. 5B–N**). Point-Biserial symptom correlation revealed positive moderate correlation with several symptoms (**Fig. 5O**).

Together, these results demonstrate that the autoantibody targets identified by immunofluorescence staining of human and mouse tissues, protein microarray, pull-down followed by mass spectrometry, Fc effector function assays, and ELISA including clinically relevant neurological and systemic autoantigens are elevated in patients with LC across two independent cohorts.

### Transfer of IgG from patients with LC into mice recapitulates pain, loss of balance and coordination, and muscle weakness

To evaluate if AABs identified in the LC group are pathogenic in vivo, we developed a mouse passive transfer model. Mice received 38.4 mg/kg of total IgG intraperitoneally (IP), with each mouse receiving IgG from a different donor. The mice were co-housed, with each cage housing one mouse that received an injection of PBS, IgG from Healthy Control, Convalescent Control, or LC donors. All the behavioral experiments were performed between one to six pm (**Fig. 6A**). IgG purity and homogeneity were confirmed by size-exclusion chromatography and SDS-PAGE (**Supp. Fig. 8A-B**).

After the injection, we evaluated the presence of IgG in the brain and serum. At 24 h post-injection, we observed human IgG crossing the blood-brain barrier, and the concentration of antibodies remained relatively constant for five days (**Supp. Fig. 8C**). Five days after injection, we observed that 5% of the human IgG crossed the blood-brain barrier, as measured by ELISA of brain homogenates (**Fig. 6B**). After injection, the mice showed no significant weight or temperature variation across groups (**Supp. Fig. 8D**). Three days post-injection, these mice displayed increased thermal pain sensitivity based on the hot plate test (**Fig. 6C**). When we compared the hot plate test with the symptoms that the donors reported, we observed that mice receiving IgG from those that met criteria for new onset pain using the SLANSS questionary (nociceptive pain or neuropathic pain) displayed shorter latency to pain behavior in hot plate test (**Fig. 6D**), with 85% of the mice with the pain phenotype (i.e. defined by the threshold for pain positivity) having received IgG from participants with LC who self-reported new-onset pain since their LC diagnosis (**Fig. 6E**). The hot plate test was also associated with pain sensations described by the LC participants as pins and needles and burning pain, and a non-statistically significant trend with electric shock (**Fig. 6F-H**). The hot plate test phenotype was also associated with weakness and dysautonomia, with 55% of the mice scoring positive for pain receiving IgG from patients who reported weakness and 90% from patients with dysautonomia (**Fig. 6I-J**). All the other symptoms evaluated were not significantly associated with the hot plate test (**Supp. Fig. 9A**). Longitudinal analysis revealed that from the first day post injection, mice receiving antibodies from Long COVID patients had a drop in their latency to pain behavior, with the most significant differences observed on days 3, 4 and 5 post injection and returning to basal levels on day 6 post injection (**Fig. 6K**). To assess reproducibility of the pain behavior assay, a blinded observer repeated the hot plate test (**Supp. Fig. 8E**). We noted excellent concordance with previous data obtained by an unblinded observer.

The grip strength test (carried out on day three post-injection) showed a trend of muscle weakness in mice injected with IgG from those with LC (**Fig. 6L**). When we segregated by symptoms, we observed an association with tinnitus (54.5% of the mice that showed muscle weakness received IgG from participants who had tinnitus) (**Fig. 6M**). We also observed a trend when we compared the grip strength test with the participants who reported headaches (90.9% of the mice with muscle weakness were injected with IgG from participants reporting headaches) (**Fig. 6N**). All the other symptoms evaluated were not significant (**Supp. Fig. 9B**).

The rotarod test was evaluated two days post-injection. While we did not observe significant differences when comparing across the groups (**Fig. 6O**), when we evaluated symptoms reported by LC participants, we observed that 88.88% of the mice that showed loss of balance and coordination had received IgG from patients who reported dizziness (**Fig. 6P**). All the other symptoms evaluated were not significant (**Supp. Fig. 9C**).

Mice injected with antibodies from individuals with Long COVID showed increased fatigue-like behavior in the treadmill test, with reduced distance traveled and total trial time (**Fig. 6Q-R**), none of the comparisons with the symptoms reported by the LC participants were significant (data not shown). We also performed a blinded whole-body necropsy analysis that showed no gross lesions and no significant findings from the histological analysis by a pathologist (**Supp. Fig. 10A**). Whole blood analysis showed no significant differences on red blood cells analysis, platelets count, and white blood cells including neutrophils, lymphocytes, monocytes, eosinophils (**Supp. Fig. 10B**). Cytokine profile in the plasma showed no significant differences on IFN-γ, IL-6, IL-10, IL-12, MCP-1, TNF-α (**Supp. Fig. 10C**). However, we observed increased levels of glutamate, IL-6 and IL-10 in the brain homogenate of mice after passive transfer of IgG from patients with Long COVID (**Fig. 6S-U**), none of the other cytokines tested were significant (**Supp. Fig. 10D**).

We also evaluated anxiety-like behavior and locomotion by open field, elevated zero maze (**Supp. Fig. 11A-C**), and blood pressure and heart rate (**Supp. Fig. 11D**) to evaluate pathological changes to orthostatic responses. We did not observe significant differences in these tests between the groups receiving IgG from healthy controls, convalescent controls, or long COVID participants.

To evaluate how those antibodies may be inducing pain, we performed passive transfer into Scn10a-Cre::tdTomato mice, which exhibit fluorescent labeling of pain-sensing neurons of the dorsal root ganglion (DRG) and their peripheral nerve axons that express the Na_V_1.8 channel encoded by the *Scn10a* gene^53^ (**Fig. 7A**). All IENF analyses were carried out in a blinded fashion. We observed that mice injected with IgG from patients with LC had a reduction in the number and volume of Intraepidermal Nerve Fibers (IENF) when compared with mice that received IgG from healthy controls (**Fig. 7B**). The reduction in the IENF is a marker for small fiber neuropathy ^54^ that usually presents with chronic pain described as burning pain and/or pins and needles sensations (both donor symptoms were associated with the hot plate test results) (**Fig. 6F-G**). We also evaluated IENF counts in the glabrous hind-paw skin from the same mice we performed the hot plate test (**Fig. 6C-D**) and found that the reduction of IENF was observed in mice that had antibodies transferred from patients reporting chronic pain (**Fig. 7C-D**). Moreover, we found human IgG labeling around the remaining nerve fibers in the epidermis. The same mice showed increased neurofilament light chain (NfL) levels in their plasma (**Fig. 7E**), suggestive of axonal damage. We observed no gross loss of peripheral innervation in the paw after the passive transfer of IgG (**Supp. Fig. 11E-G**), suggesting that the IgG-induced pathology is confined to the local intraepidermal nerve fibers.

To visualize neuronal activation underlying pain and fatigue-like behavior following passive IgG transfer, we performed brain-wide neuronal activity mapping by evaluating c-Fos expression with iDISCO+ analysis three days after the injection (**Fig. 7F**). The mice were assessed without any behavioral or sensory stimulation, enabling us to evaluate the steady-state neuronal activation pattern driven by transferred antibodies alone. Unbiased quantitative mapping showed widespread neuron activation across areas associated with pain, fatigue, and cognitive-emotional dysregulation. Notably, increased c-Fos+ cells were observed in somatosensory/introceptive regions (blue), including primary and supplemental somatosensory areas, visceral area, medial lemniscus, and agranular insular area, consistent with increased nociceptive signaling. Concurrent activation of motor and action initiation areas (purple), including primary motor area, pontine reticular nucleus, tegmental reticular nucleus, and pontine gray, potentially reflecting motor slowing, decreased activity, or perceived fatigue (**Fig. 7G, Supp. Fig. 11H**).

Regions in the affective and emotional cluster (orange), including anterior cingulate, amygdala, and claustrum, can mediate the emotional distress associated with pain, contributing to affective discomfort and fatigue-like malaise. Limbic-olfactory integration cluster (pink), including piriform area, endopiriform nucleus, entorhinal area, perirhinal area, and ectorhinal area, linking internal state awareness with learned associations. Cognitive and mnemonic cluster (green), including retrosplenial, temporal association, and CA1 and CA3 (hippocampus), contributes to attention, spatial/contextual memory, and task persistence, often impaired in fatigue states. Sensory association cluster (yellow), including primary auditory and primary visual regions, is consistent with hypervigilance or sensory overload seen in chronic pain/fatigue. Finally, the arousal and neuromodulatory brainstem cluster (red), including the midbrain reticular nucleus, Edinger-Westphal nucleus, and rostral linear nucleus raphe, plays critical roles in sleep-wake regulation, descending pain control, and homeostatic tone. Dysregulation of these structures may contribute to fatigue and pain persistence. This data suggests that systemic autoantibodies engage the activation of brain regions associated with pain, fatigue, and cognitive dysfunction, via engagement of sensory, affective, and arousal pathways (**Fig. 6G, Supp. Fig. 11D**).

Together, these data show that passive transfer of total IgG from patients with LC induces heightened pain sensitivity, fatigue-like behavior, loss of balance and coordination, and muscle weakness in mice, and is associated with the symptoms reported by those participants, namely chronic pain, headache, tinnitus, dizziness, and dysautonomia. Moreover, widespread neuron activation consistent with chronic pain and fatigue was revealed through whole brain cFos mapping after passive transfer of IgG.

## Discussion

In this study, we evaluated the presence of AABs and their pathological potential in LC. Our data show that a subset of LC participants had AAB against several tissues, including the locus coeruleus and thalamus, and cross-reactive antibodies against mouse meninges and sciatic nerve. By employing a comprehensive human autoantigen microarray and ELISA for GPCR and ionotropic receptors, we demonstrated that LC patients’ IgG reacts against a wide array of antigens that are enriched in the nervous system tissues. We also validated the finding that Long COVID patients have increased autoantibodies against clinically relevant common autoantigens in an independent cohort. We validated some of the microarray hits using two other orthogonal methods, mass spectrometry and ELISA. To evaluate their pathogenic potential, we used two independent approaches: first, we performed an in vitro analysis of isotype and subtype analysis, Fc receptor binding, antibody-dependent complement deposition, and antibody-dependent phagocytosis using a candidate autoantigen, MED20; second, we passively transferred purified IgG from patients into mice. We showed that these antibodies are sufficient to recapitulate key neurological symptoms observed in patients with LC, as well as to cause loss of intraepidermal nerve fibers and activation of neurons in pain-sensing regions of the CNS. Collectively, our data illustrate the pivotal role of autoantibodies as a key driver of neurological disorders in a subset of LC patients.

Using a broad range of tissues and performing immunofluorescence using total IgG purified from LC participants, we were able to identify a subset of participants with AAB against neurological tissues and symptoms that are more closely associated with those AAB. In particular, autoreactivity involving the locus coeruleus raises the possibility that disruption of central noradrenergic circuits may contribute to persistent autonomic dysregulation, impaired stress adaptation, and fatigue in LC. Similarly, autoantibodies recognizing the thalamus may disturb central networks, consistent with functional MRI observations demonstrating altered thalamic connectivity in ME/CFS^55^. Supporting the presence of neuro-reactive antibodies in LC, a recent case report^20^ showed cross-reactive antibodies in the cerebrospinal fluid against several mouse neurological tissues, including the meninges and sciatic nerve, in a patient with severe neurological symptoms following SARS-CoV-2 infection. Among participants with cross-reactive antibodies against mouse meninges, we observed co-localization of those antibodies with pericytes and endothelial cells. These vascular-reactive AAB may explain their association with reported headache among LC participants, since changes in pericyte functions have been described to play a role in the vascular mechanisms associated with migraine with aura^56^.

Our findings showed high diversity of AAB found in individuals with LC, illustrated by immunofluorescence, GPCR/ionotropic receptor ELISA, HuProt™, and mass spectrometry data. This diversity is expected since LC likely consists of multiple endotypes, with a broad range of symptoms that vary from person to person, with distinct disease drivers including persistent virus, herpesvirus reactivation, tissue damage, and autoimmunity ^57^.

When comparing the HuProt and MS data, we applied different approaches. In the HuProt assay, we screened against the entire human proteome, whereas in the MS experiments, we used human brain homogenate, resulting in an enrichment of neurological targets. The pull-down using brain homogenate under non-denaturing conditions also favored the detection of transmembrane proteins, in contrast with the HuProt platform, where purified proteins are immobilized on a slide. Together, these complementary methodologies strengthened our findings and informed the selection of candidates for targeted validation.

Using a human protein array, mass spectrometry, and ELISA, we validated seventeen potential AAB targets, with NMDAR2C, NCOA3 and Syncytin-2 being the most prominent AAB targets. Importantly, whether these autoantibodies are pathogenic or directly contribute to symptom development remains unknown. Nevertheless, autoantibodies directed against these proteins may contribute to hormonal dysregulation or neuroinflammation. NCOA3 plays a critical role in regulating the transcriptional activity of estrogen, progesterone, and androgen receptors^58^. Syncytin-2 is encoded by the ERVFRD-1 gene derived from an endogenous retrovirus and functions as a fusogenic protein critical for implantation of embryo^59^. Little is known regarding autoantibody targeting syncytin-2. Other targets were antibodies against MED20, which can play a role in the development of LC neurological symptoms, since Mediator Complex (MED) proteins, such as MED12 and MED23, have been associated with neurological diseases^60,61^. It should be noted that reactivity against MED20 was present in only a subset of LC patients, emphasizing the diversity of AABs and LC phenotypes. To further investigate potential mechanisms linking autoantibodies to clinical manifestations, we observed that LC patients exhibit a reduced IgM/IgG ratio against MED20, consistent with enhanced class-switch recombination in this cohort. This shift was accompanied by a bias toward activating Fc receptor engagement, indicating a more pro-inflammatory effector profile in LC. Concordantly, we detected increased antigen-specific antibody-dependent phagocytosis in LC patients.

Our identification of autoantibodies targeting the NMDA receptor subunit NR2C (GRIN2C) in a subset of LC patients is in concordance with previous papers showing autoantibodies against GPCRs in Long COVID^23,26,27,62^. NMDA receptors are central regulators of excitatory synaptic transmission, synaptic plasticity, and learning and memory^63^. Autoantibody-mediated disruption of NMDA receptor signaling is well established in anti-NMDAR encephalitis, where antibodies primarily targeting the NR1 subunit cause receptor internalization and hypofunction, leading to neuropsychiatric manifestations^64–66^. Whether or not the anti-NMDAR IgG found in our LC patients is pathogenic remains to be determined.

Future studies are required to distinguish pathological autoantibodies from disease-irrelevant autoantibodies, which remains a key challenge in understanding Long COVID. Our study found that HuProt was poor at distinguishing LC vs. CVC, while more targeted antibody analysis using clinically relevant autoantigens, GPCR, ionotropic receptors, mass spec analysis of antigens after IgG-pull down, and systems serology had a better utility in distinguishing LC vs. CVC. Two clusters based on AAB reactivity, based on ELISA and HuProt, appear to target neural tissue cell types (cluster 1) vs. reproductive/vasculature/endocrine tissue cell types (cluster 2). While these clusters are not associated with a particular symptom, testing the consequences of AAB targeting identified targets are needed to determine the precise impact on host physiology. Using our passive transfer model, we observed that chronic pain is the symptom most strongly associated with AABs, providing direct functional evidence that antibodies from a subset can exert pathogenic effects.

Our human cohort included male and female participants, normalized by sex and age for the MyLC II validation cohort and only by sex for the MyLC cohort. Linear regression models adjusted by age and sex was performed for the autoantibody analysis. Biological sex was recorded based on self-report and included as a covariate in relevant analyses. Consistent with prior epidemiological reports, females were overrepresented among Long COVID participants, reflecting the known sex bias in Long COVID prevalence. For the passive transfer mouse model, female mice were used to reflect the female predominance observed in Long COVID patients. Tissue immunofluorescence analyses were performed using samples from both male and female mice; no sex-based differences in tissue immunofluorescence were observed.

Our results provide compelling evidence that passive transfer of total IgG from individuals with Long COVID induces pain and fatigue-like behavior in mice, resembling key symptoms reported by the patients^45^. Independent studies have reported similar findings^67^. Increased sensitivity to pain and IEFN damage had also been described after a passive transfer of autoantibodies from fibromyalgia patients^68^. Through iDISCO+ analysis, we observed widespread neuronal activation across brain regions associated with pain, fatigue, and emotional dysregulation. In addition to the central nervous system findings, we also explored the peripheral nervous system by analyzing the intraepidermal nerve fibers (IENF) in mice, revealing a reduction in both the number and volume of IENF, a diagnostic criterion for small fiber neuropathy. This phenotype in mice was associated with chronic pain and symptoms such as burning pain and pins and needles reported by the donor patients. Small fiber neuropathy is a common finding in Long COVID patients^69,70^. The activation of brain regions involved in pain processing, combined with peripheral nerve damage as evidenced by reduced IENFs, supports the idea that systemic autoantibodies from Long COVID patients engage both central and peripheral mechanisms to induce chronic pain. Furthermore, the association of HuProt-identified autoantibodies targets with headache, along with immunofluorescence findings showing increased autoreactivity to meninges, supports the hypothesis that AABs mediate persistent pain symptoms observed in Long COVID.

For the LC endotype driven by AABs, there may be existing therapies that would counter the pathologic impacts of the AABs. For example, intravenous immunoglobulin (IVIg) treatment might work by neutralization and clearance of AABs^71^. An analysis of 130 cases including IVIg treatment for neuro-COVID reveals that this technique is beneficial for patients with LC and ME/CFS^72^. A review of IVIg usage in post-COVID neurological disorders revealed that while virtually all studies showed benefit from IVIg treatment in patients with post-COVID neurological disorders, only 12 of 76 case reports (16%) demonstrated unequivocal diagnoses of autoimmune encephalitis. One in every four of these cases (26%) lacked evidence to support such a diagnosis^72^. Benefits from IVIg treatment were also reported in LC patients with small fiber neuropathy^73^. However, IVIg does not help all patients. Currently, there are no guidelines or clinical tests to distinguish between those who would benefit and those who do not, underscoring the urgent need to develop new biomarkers. Demonstration of functional AAB may be used as a diagnostic tool to inform IVIg therapy. In addition to IVIg, AAB-driven disease may be treatable with mechanisms to degrade circulating antibodies (e.g., FcRn inhibitors), neutralize their action (e.g., BC 007^74^), remove them temporarily (e.g., immunoadsorption^75^), or targeting of antibody-secreting cells (e.g., anti-CD20 or anti-CD38 monoclonal antibodies^76^).

Currently, AABs are not widely recognized as a driver of post-COVID neurological symptoms. Based on our data, an array of common autoantigens or a comprehensive human proteome would not be suitable as a tool to diagnose AAB-LC endotype. Effective diagnosis of this LC endotype will likely require an optimized panel of autoantigens that can sensitively and accurately detect functional AABs. In the absence of such tools, passive transfer of IgG from patients into mice followed by measurement of key validated behavioral phenotypes could provide an interim method, although not scalable. Finally, the use of blood products, including immunoglobulins from people with LC, in treatment settings requires careful consideration, as they may pose pathologic risks to the recipients. Our data provide key pathological roles of AAB in driving neurological phenotypes in vivo and warrant future studies to inform diagnosis and therapy for this subset of LC.

### Limitations of the Study

While this study provides a mechanistic link between autoantibodies and the pathology of Long COVID, several limitations exist. First, the broad screening performed using immunofluorescence, ELISA for GPCR, HuProt, and mass spectrometry could result in false-positive targets, particularly in the detection of low-abundance autoantibodies. It is essential to consider a potential selection bias in the seventeen hits validated by ELISA, which may not be the best targets to identify Long COVID patients. For autoantibody discovery platforms (HuProt HC mean + 3 SD; mass spectrometry HC mean + 5 SD). whereas validation and exploratory analyses used more permissive cutoffs (HC mean + 1–2 SD, depending on assay), which may capture low-level background reactivity. ROC analyses were performed across platforms to evaluate discriminatory capacity and support threshold selection. For ROC analysis HC and CVC group was pooled together to reflect the ability of these assays to distinguish Long COVID patients from individuals who recovered from COVID-19 without persistent symptoms. Given the substantial heterogeneity in autoantibody reactivity within the discovery cohort, the external validation cohort was used to confirm that autoantibody dysregulation is a reproducible feature of Long COVID in an independent sample. However, this cohort assessed a predefined panel of 118 established clinical autoantigens rather than the top-performing discovery candidates directly, limiting target-level replication. Direct validation of top discovery targets in independent cohorts remains a priority for future work. This represents a limitation of the current study design, and future prospective studies should prioritize direct validation of top-performing targets. For our cluster analysis, we didn’t observe perfect cluster separation, and partially overlapping clusters might be consistent with biologically continuous phenotypes in complex immune-mediated diseases.

Additionally, while passive transfer of human IgG to mice offers functional validation, it fails to fully capture all the symptoms described by the patients. This study also relied on a single time point analysis, which limits the ability to assess the longitudinal changes in symptom progression. Extended longitudinal analysis with repeated injections was not feasible due to the limited availability of patient-derived IgG. In addition, the clinical heterogeneity of Long COVID patients and the small cohort size that we evaluated may introduce confounding variables, affecting the generalizability of the findings. Accordingly, our in vivo experiments should be interpreted as proof-of-concept for the functional activity of circulating autoantibodies, and future studies using larger sample volumes will be required to determine durability and dose-dependence of these effects.

We did not directly assess blood–brain barrier integrity or permeability following IgG transfer. Thus, while CNS effects were observed, the mechanisms of antibody entry into the brain and its impact on local cytokine responses could not be determined in this study, which was designed to evaluate the functional consequences of circulating autoantibodies. Finally, future research is necessary to elucidate the mechanisms and specificities of autoantibodies responsible for *in vivo* pathology.

## Resource Availability

### Lead Contact

Further information and requests for resources and reagents should be directed to and will be fulfilled by the lead contact, Akiko Iwasaki

## Materials Availability

This study did not generate new unique reagents.

## Data and Code Availability

Analysis code is available on GitHub (https://github.com/keyla-sa/A-causal-link-between-autoantibodies-and-neurological-symptoms-in-long-COVID).

Processed GPCR/ionotropic receptor ELISA, mass spectrometry, Immunofluorescence, antibody effector function analysis, and autoantibody screening in the validation cohort have been deposited to ImmPort (Study Accession SDY3242) and are publicly available as of the date of the publication.

The HuProt autoantibody array data generated in this study have been deposited in Yale Dataverse under DOI 10.60600/YU/DRGWSZ and are publicly available as of the date of the publication.

## Data Availability

Analysis code is available on GitHub (https://github.com/keyla-sa/A-causal-link-between-autoantibodies-and-neurological-symptoms-in-long-COVID).
Processed GPCR/ionotropic receptor ELISA, mass spectrometry, Immunofluorescence, antibody effector function analysis, and autoantibody screening in the validation cohort have been deposited to ImmPort (Study Accession SDY3242) and are publicly available as of the date of the publication.
The HuProt autoantibody array data generated in this study have been deposited in Yale Dataverse under DOI 10.60600/YU/DRGWSZ and are publicly available as of the date of the publication.

https://doi.org/10.60600/YU/DRGWSZ

## Acknowledgements

We thank the members of the MY-LC study community who volunteered their time and effort to aid in the completion of this study and who also helped inform and educate on the practical and equitable communication of the results.

We are also grateful to Huiping Dong, Melissa Linehan, Suzanne Fischer, Victoria Fisher, and Pavlina Baenova for technical support.

We are grateful to the Yale Rodent Behavior Analysis facility for providing dedicated space and equipment for the behavioral analyses reported in this paper.

We acknowledge the Yale Cancer Center/Pathology Tissue Microarray Facility and the Department of Pathology for providing all the peripheral human tissues used in this study.

We acknowledge the Yale Pathology Dept., the Autopsy Service, and the Center for Human Brain Discovery, which is supported by philanthropic funds, in particular Dr. Gopal Pallavi and Amanda Masters, for providing human brain and non-CNS tissue.

We thank the Yale West Campus Imaging Core for the support and assistance in this work.

We thank the Yale Comparative Pathology Research Core for the necropsy and Carmen J. Booth, DVM, PhD, for the pathologic analysis performed in the mice after passive transfer.

Various graphical schematics were created using BioRender.

We thank Nils Kort for providing the meninges from NG2-dsRed mice.

This work was supported by the Else Kröner Fresenius Prize for Medical Research 2023, grants from the National Institute of Allergy and Infectious Diseases (R01AI157488 to A.I.), RTW Foundation (D.P.), the Howard Hughes Medical Institute Collaborative COVID-19 Initiative (A.I.), Howard Hughes Medical Institute Emerging Pathogens Initiative (A.I.), and the Howard Hughes Medical Institute (A.I.). Steve and Alexandra Cohen Foundation (D.P. and J.W.), Nash Family Foundation (D.P. and J.W.), and Polybio Research Foundation (D.P. and J.W.). D.K. receives research support from the Yale Predoctoral Pharmacology Training Program (PPTP, grant number T32 GM156537). K.S.G.S. receives research support from the Pew Latin American Fellowship from The Pew Charitable Trusts.

## Author Contributions

Conceptualization: K.S.G.S; T.L.H.; A.I.

Data Curation: K.S.G.S;

Formal Analysis: K.S.G.S; J.S.; C.A.B.; D.N.; B.C.; L.W.; D.N.; B.C.; L.W.;

Funding Acquision: J.W.; D.P.; A.I.;

Investigation: K.S.G.S; R.B.O.; C.A.B.; Z.L.; W.G.; D.N.; B.C.; L.W.; D.K.; R.B.; R.A.R.C.; B.O.; P.A.C.D.; D.K.; G.R.; H.H.; K.S.F.; A.G.; T.S.; A.C.; T.L.; A.B.; L.B.; H.B.; K.G.; S.B.;

Methodology: K.S.G.S;

Resources: S.J.; L.M.; B.B.; J.G.; J.W.; L.T.; C.S.; Y.L.; L.G.; M.S.P.; D.P.; A.I.;

Supervision: T.L.H.; A.I.;

Validation: K.S.G.S; A.I.; Visualization: K.S.G.S;

Writing – Original Draft: K.S.G.S;

Writing – Review and Editing: J.S.; R.B.O.; C.A.B.; Z.L.; W.G.; D.N.; B.C.; L.W.; D.K.; R.B.; R.A.R.C.; B.O.; P.A.C.D.; D.K.; G.R.; H.H.; K.S.F.; A.G.; T.S.; A.C.; T.L.; A.B.; L.B.; H.B.; K.G.; S.B.; S.J.; L.M.; B.B.; J.G.; J.W.; L.T.; C.S.; Y.L.; L.G.; M.S.P.; D.P.; T.L.H.; A.I.;

## STAR METHODS

### EXPERIMENTAL MODEL AND STUDY PARTICIPANT DETAILS

#### Ethics statement involving human studies

This study was approved by the Mount Sinai Program for the Protection of Human Subjects (IRB 20-01758) and Yale Institutional Review Board (IRB 2000029451 for MY-LC; IRB 2000028924 for enrolment of pre-vaccinated Healthy Controls, IRB 2000036624 for lupus samples). Informed consent was obtained from all enrolled participants.

#### MY-LC enrollment

MY-LC study design, inclusion, and exclusion criteria for recruitment of individuals have been described in detail previously^42–45^. Briefly, individuals experiencing persistent symptoms for > 6 weeks after initial SARS-CoV-2 infection were invited to participate in the study from various LC clinics within the Mount Sinai Health System and the Cohen Center for Recovery from Complex Chronic Illness at The Mount Sinai Hospital. The participants who enrolled in the LC group underwent complete medical evaluations by physicians to rule out alternative medical etiologies for their persistent symptoms before study enrolment, following the NASEM criteria for Long COVID diagnosis^77^. Those joining either the healthy or convalescent study groups were recruited through IRB-approved advertisements disseminated via email lists, study flyers in hospital public areas, and various social media platforms. Before enrollment, all participants provided informed consent. On the day of sample collection, each participant supplied peripheral blood samples and completed symptom surveys. Additionally, self-reported medical histories of all participants in the MY-LC cohort were collected during study visits and cross-checked by collaborating clinicians through the examination of electronic medical records. Individuals with a history of autoimmune disease were excluded from the analysis.

To independently validate findings derived from the primary MY-LC cohort, an additional cohort (MY-LC II) was recruited using the same study design, inclusion and exclusion criteria, recruitment sources, clinical evaluation procedures, biospecimen collection protocols, and symptom assessment instruments as the original MY-LC cohort. MY-LC II participants were enrolled at a later time period (November 2022-March 2023) and were analyzed as a fully independent validation cohort. Clinical and demographic information of the MY-LC II participants is summarized in Supplementary Table 2.

Self-reported symptoms assessed across both cohorts included vomiting, skin lesions (rash or lumpy lesions), difficulty swallowing, loss of taste, swelling, fainting or blackouts, loss of smell, sore throat, abdominal pain, joint pain or swelling, diarrhea, nausea, muscle pain or cramps, chest pain or discomfort, unexplained hair loss, difficulty sleeping (too much, too little, early awakening), problems seeing (double or blurry vision), fever or chills, confusion/difficulty thinking, general/muscle weakness, breathing faster than normal (tachypnea), cough, dizziness or lightheadedness, headache, unexplained sweats or flushing, dysautonomia, chronic pain (SLANSS), abnormal changes in body temperature, loss of appetite or unexplained weight loss, mood swings/irritability/depression, memory problems or forgetfulness, disorientation (getting lost; going to wrong places), fatigue or tiredness, tinnitus (ringing in the ears), tingling/numbness/burning/stabbing or “pins and needles”, shortness of breath (dyspnea), unexplained menstrual irregularity, heart palpitations/pulse skips/heart block, difficulty hearing, indigestion or esophageal/“acid” reflux, difficulty with concentration or reading or “brain fog”, exaggerated symptoms or worse hangover from alcohol, urinary incontinence or difficulty urinating, tingling and numbness in the mouth and/or face, bloating, and sexual dysfunction or loss of libido.

#### Animals

Behavioral experiments were performed on C57BL/6J (Jackson Laboratory, stock number 000664) female and male mice aged between six and eight weeks. Mice were maintained under specific pathogen-free conditions at Yale University. Mice were maintained in ventilated cages at an ambient temperature and humidity-controlled room with a 12 h light/12 h dark cycle with continuous access to food and water. The experiments using animals were performed according to the federal guidelines and the institutional policies of the Yale School of Medicine Animal Care and Use Committee (Approved Protocol 2024-10365 and 2022-07942). Given that Long COVID predominantly affects females, only female mice were used in the in vivo experiments.

## METHOD DETAILS

### Human tissue dissection

Human autopsy was performed by the Yale Pathology Department Autopsy Service and Center of Human Brain Discovery on one male, in his 40’s. The underlying cause of death was hypertensive cardiovascular disease (mSOP44). Dissected tissues were putamen, occipital cortex, spinal cord, hippocampus, middle temporal gyrus, thalamus, cerebellum, caudate, locus coeruleus, pituitary, amygdala, olfactory bulb, hypothalamus, parietal cortex, periventricular white matter, and substantia nigra. For each tissue, two independent biopsies were collected. One was fixed overnight (ON) in 4% paraformaldehyde (PFA), embedded in optimal cutting temperature (OCT) compound (Sakura Finetek, 4583), and subsequently frozen using a dry ice/acetone cooling bath. The other biopsy was processed fresh for mass spectrometry analysis.

The peripheral tissue microarray was customized with healthy human peripheral tissues from the tissue bank of the Yale Cancer Center/Pathology Tissue Microarray Facility and the Department of Pathology. All tissue samples were embedded in paraffin and fixed in formalin (Formalin-Fixed Paraffin-Embedded, FFPE).

### Purification of immunoglobulins

Total IgG was purified from serum samples using Protein-G beads (Sigma-Aldrich, St. Louis, MO, US). One and a half mL of serum samples were diluted 1:2 with phosphate-buffered saline (PBS) and incubated with 2 mL of Protein-G Sepharose beads (Millipore Sigma, GE17-0618-05) ON at 4 °C. Samples were transferred to elution columns, and depleted sera (flow-through) were collected and stored at –80 °C for later use. Beads were washed 3 times with PBS, and eluted using 100 mM glycine, pH 2.3, into six fractions. The eluates were neutralized to pH 7.4 using 1 M Tris pH 8, centrifuged, and the protein concentration was determined using a NanoDrop 8000 (Thermo Fisher Scientific, Waltham, MA, US). Fractions containing antibodies were pooled and dialyzed ON in PBS at 4 °C. Dialyzed samples were quantified, aliquoted, and stored at 4 °C.

### Characterization of human IgG quality

Fast Protein Liquid Chromatography (FPLC) was performed using a Quest 10 Plus NGC Chromatography System (Bio-Rad Laboratories Inc., Hercules, CA, US; 7880003) fitted with an ENrich SEC 650 10x300 column (Bio-Rad, 780-1650) equilibrated with filtered and degassed PBS. Fifty µL of total human IgG at 1 mg/mL (total 0.05 mg) were injected into the column at a flow rate of 0.5 mL/min. Bovine thyroglobulin (670 kDa), bovine γ-globulin (158 kDa), and chicken ovalbumin (44 kDa) (all from the gel filtration standard, Bio-Rad, 1511901) were used to estimate the MW of the IgG elution peaks. The SEC traces were analyzed using ChromLab Software (Bio-Rad).

A total of 0.04 mg of IgG was used for the analysis. Under reducing conditions, the antibodies were heated at 100 °C for 5 min in Laemmli buffer, loaded onto an SDS-PAGE gel, and stained with Coomassie Blue. For non-reducing conditions, the antibodies were mixed with LDS sample loading buffer and loaded onto an polyacrilamide gel without prior heating, followed by Coomassie Blue staining.

Endotoxin test was performed on random samples per batch of extraction using endosafe® nexgen-PTS^TM^ (Charles River), on average, samples had values of 0.202-0.0112 EU/mg of IgG. For reference, the accepted limit for injection in mice is 5 EU/kg^78^, which corresponds to 0.1 EU for a 20 g mouse.

### Autoantigen pull-down

Fresh neurological human tissues were dissected as described above and placed into 1 mL of non-denaturing lysis buffer (20 mM Tris HCl, pH 8, 137 mM NaCl, 1% Nonidet P-40, 2 mM EDTA) with complete protease inhibitor cocktail (Roche, Sigma, 11697498001) on ice. Tissues were then homogenized with MP FastPrep -24 at speed 2 for 20 s, then centrifuged at 14,000 g for 10 min, at 4 °C. The supernatant was aliquoted and frozen at −20 °C until use. For pre-clearing of supernatants, Protein G Sepharose beads (Millipore Sigma, GE17-0618-05) were washed with non-denaturing lysis buffer with complete protease inhibitor cocktail, then equilibrated in PBS. Subsequently, 100 µL beads were incubated with 100 µL pooled brain supernatant, constantly mixing for 1 h, at 4 °C. Beads were pelleted at 100 g for 3 min at 4 °C, discarded, and the supernatant extracted.

Four µg of total IgG purified from the participants were mixed with Protein G Sepharose beads (Millipore Sigma, GE17-0618-05) and incubated ON at 4 °C on a rocking platform. IgG/protein G bead complexes were incubated with pre-cleared supernatants for 1 h at 4 °C in a rocking platform. Beads were recovered by centrifugation at 100 g for 3 min at 4 °C, then washed 4 times with non-denaturing lysis buffer with complete protease inhibitor cocktail. Beads were again recovered by centrifugation, then eluted using 100 μL of elution buffer (100 mM glycine, pH 2.3). The eluates were neutralized to pH 7.4 using 1 M Tris pH 8, centrifuged, and dialyzed ON in PBS at 4 °C. Dialyzed samples were quantified by NanoDrop 8000 (Thermo Fisher Scientific, Waltham, MA, US), aliquoted, and stored at 4 °C.

### Antibody pull-down followed by mass spectrometry - sample preparation and data processing

A total of 20 μL of the above immunoprecipitated samples were digested for the mass spectrometry-based proteomic analysis. For each sample, 30 μL of 10 M urea was added. Reduction and alkylation were carried out using 10 mM Dithiothreitol (DTT) for 1 h at 56 °C, followed by 20 mM iodoacetamide (IAA) in darkness for 45 min at room temperature (RT). The samples were then diluted with 100 mM NH4HCO3 and digested with trypsin (Promega) at a ratio of 1:20 (w/w) ON at 37 °C. The purification of the digested peptides was performed using a C18 column (MacroSpin Columns, NEST Group Inc.). About 1 μg of the peptide was injected for mass spectrometry measurement.

The samples were measured by a data-independent acquisition (DIA) MS method as described previously^79,80^, on an Orbitrap Fusion Tribrid mass spectrometer (Thermo Scientific) coupled to a nanoelectrospray ion source (NanoFlex, Thermo Scientific) and an EASY-nLC 1200 system (Thermo Scientific, San Jose, CA). A 120-min gradient was used for the data acquisition at a flow rate of 300 nL/min with the column temperature controlled at 60 °C using a column oven (PRSO-V1, Sonation GmbH, Biberach, Germany). The DIA-MS method consisted of one MS1 scan and 33 MS2 scans of variable isolated windows with 1 m/z overlapping between windows. The MS1 scan range was 350–1650 m/z, and the MS1 resolution was 120,000 at m/z 200. The MS1 full scan AGC target value was set to be 2E6, and the maximum injection time was 100 ms. The MS2 resolution was set to 30,000 at m/z 200 with the MS2 scan range 200–1800 m/z, and the normalized HCD collision energy was 28%. The MS2 AGC was set to be 1.5E6, and the maximum injection time was 50 ms. The default peptide charge state was set to 2. Both MS1 and MS2 spectra were recorded in profile mode. DIA-MS data analysis was performed using Spectronaut v18^81^ with the directDIA algorithm by searching against the SwissProt human fasta sequences (September 2022). The oxidation at methionine was set as a variable modification, whereas carbamidomethylation at cysteine was set as a fixed modification. Both peptide and protein FDR cutoffs (q-value) were controlled below 1% (q ≤ 0.01) using Spectronaut’s target–decoy search strategy with q-value estimation (Storey–Tibshirani procedure). Peptide- and protein-level identifications that passed this threshold were retained, and the resulting quantitative data matrix was exported from Spectronaut. The sample normalization was off. All the other settings in Spectronaut were kept as Default.

### Immunofluorescence staining

Mice were euthanized and perfused with PBS and 4% paraformaldehyde (PFA). For meninges analyses, NG2DsRedBAC (Jackson, 008241) was used. Human tissues were dissected as described above. Target tissues were dissected and postfixed ON with 4% PFA, embedded in optimal cutting temperature (OCT) compound (Sakura Finetek, 4583), and then frozen on a cooling bath with acetone/dry ice. Tissues were stored at –80 °C. Tissues were sectioned with a cryostat at a thickness of 10 µm and thaw-mounted onto SuperFrost Plus Slides (Thermo Fisher Scientific). Slides were stored at – 80 °C until use.

For autoantibodies staining using purified total human IgG, the antibodies were conjugated with Alexa Fluor 488 using Alexa Fluor™ 488 Protein Labeling Kit (Thermo Scientific, A10235) following the manufacturer’s instructions.

For FFPE tissues, the slides were subjected to deparaffinization following three sequential xylene baths for 10 min each, and then rehydrated through a grade of ethanol 100% 2 times 10 min each, 95% for 5 min, 70% for 5 min, 50% for 5 min, and rinsed in distilled water for 5 min. The antigen retrieval was performed by incubating the tissue sections in 0.1% trypsin solution in PBS at 37 °C for 15 min. After enzymatic antigen retrieval, slides were washed in PBS for 5 min. For frozen tissues, slides were thawed at RT and washed in PBS to remove excess OCT. For both FFPE and frozen tissues, samples were outlined using a hydrophobic PAP pen.

Slides were blocked with a solution of 1% bovine serum albumin (BSA) and glycine (22 mg/mL) in a humidified chamber for 1 h. A 0.3% Sudan Black B/70% ethyl alcohol (EtOH) solution was added to each sample and incubated before being washed thoroughly with PBS. Samples were incubated with 0.039 mg/ml of purified total IgG. For staining endothelial cells in the meninges, the samples were also incubated with Alexa Fluor® 647 anti-mouse CD31 Antibody (Biolegend, 102516). Each sample was assigned a patient antibody ID number and incubated with said patient IgG ON in a humidified chamber. For mouse meninges and mouse sciatic nerve, slides were washed in PBS and incubated with Alexa Fluor® 488 AffiniPure™ Donkey Anti-Human IgG (Jackson ImmunoResearch, 709-545-149) in a humidified chamber for 60 min, for Human-on-Human immunofluorescence (human tissue microarray and human brain tissues) the total IgG purified from the patients were labeled with Alexa Fluor 488 using Alexa Fluor 488 Protein Labeling Kit (Thermo Fisher Scientific, A10235), without the use of secondary antibody. Slides were once again washed in PBS and immersed in distilled water. Washed slides were mounted with a solution of DAPI Prolong® Gold antifade reagent (Invitrogen, Thermo Fisher Scientific, P36930) with 17.5 µg/ml of DAPI and covered with a coverslip. Slides were stored in the dark at 4 °C. Images were acquired by the Stellaris Confocal microscope at ×20 magnification and LAS X software (both from Leica Microsystems, Wetzlar, Germany). The percentage of stained area and mean of fluorescence were calculated using ImageJ software.

#### Autoantibody analysis by HuProt

Plasma samples were analyzed using the ImmuneProfiler on HuProt™ human proteome arrays (CDI Laboratories, Baltimore, MD, US). Arrays and plasma samples, diluted 1:1000 into CDISampleBuffer, were blocked for 1 h. Each blocked and diluted sample was then probed onto a HuProt™ microarray for 1 h. Following the probing step, the arrays underwent three 10-min washes with TBS-T (TBS, 0.1% Tween 20). Subsequently, they were probed with Alexa 647-anti-human IgG Fc specific for 1 h within a light-proof box. This was followed by three washes with TBS-T, each lasting 10 min, and three rinses with ddH_2_O. Blocking and all incubation steps were performed at RT with gentle shaking. The arrays were then dried with an air duster and scanned using a GenePix® 4000B scanner (Molecular Devices, San Jose, CA, US).

### Validation of autoantigens by ELISA

Top hits identified by HuProt™, MED20 (MyBiosource, San Diego, CA, US; MBS5308819) and USP5 (MyBiosource, MBS8121224), NCOA (MyBioSource, MBS1561562), ERVFRD-1 (MyBioSource, MBS1020059), BCL10 (MyBioSource, MBS1164977), FGD3 (MyBioSource, MBS1299828), ERP27 (MyBioSource, MBS2556608), RSNB1L (MyBioSource, MBS1429671), ECE1 (MyBioSource, MBS2009889), CBX1 (MyBioSource, MBS957039), PSIP1 (MyBioSource, MBS9420574), PRR27 (MyBioSource, MBS1417189), FAM84A (MyBioSource, MBS204842), SPN (MyBioSource, MBS205051), P3H4 (MyBioSource, MBS1577638), UBL7 (MyBioSource, MBS1392728), PAG1 (MyBioSource, MBS7101460), Heparin (heparin coated plates, VWR, 20140005-3), FGFR3 (MyBioSource, MBS952862), Plexin D1 (R&D Systems, 4160-PD-050), were coated on Maxisorb microtiter 96-well plates (Thermo Fisher Scientific, 439454) in a 2-fold dilution (starting concentration 10 µg/mL) ON at 4 °C. Plates were washed three times using PBS-T (PBS, 0.05% Tween 20), and the buffer was left in the wells for 1 h before applying the samples Purified human IgG at a concentration of 25 µg/mL and 10 µg/mL in PBS-T with 2% fetal calf serum was added to the plates and incubated for 2 h at RT shaking at 80 rpm. Plates were washed three times using PBS-T and incubated with polyclonal rabbit anti-human IgG/HRP (1:2,000; Agilent P021402-2) for 1 h at RT, shaking at 80 rpm. Plates were washed five times using PBS-T and incubated in the dark with 100 µL/well of 1-Step Ultra TMB (Thermo Fisher Scientific, 34029) for 10 min. The reaction was stopped with 50 µL of 0.5 M H_2_SO_4_. The plates were read at 450 nm with 630 nm subtraction on a Cytation 5 reader (BioTek).

#### ANA IgG ELISA

Antinuclear IgG antibodies (ANA) were measured from serum using a commercial ANA screening ELISA according to the manufacturer’s instructions (ORGENTEC Diagnostika GmbH, Germany). Absorbance was measured at 450 nm using a microplate reader (Cytation 5 reader; BioTek). Results were expressed as an index value relative to the kit calibrator, and samples were classified as negative or positive based on the assay-specific cutoff criteria.

#### Antigen-specific antibody isotype/subclass analysis

Plasma samples were analyzed using the antibody isotyping/subclassing multiplex assay (SeromYx Systems, Woburn, Massachusetts), which uses fluorescently coated microspheres to profile the isotype/subclass distribution of serum samples in an antigen-specific manner. Briefly, MED20 recombinant protein (MyBioSource; MBS5308819) and a negative control antigen (Feline Immunodeficiency Virus core protein) were covalently coupled to Magplex Luminex beads via a two-step carbodiimide reaction. The beads were activated for 30 min at RT using 100 mM monobasic sodium phosphate, pH 6.2 with 5 mg/mL Sulfo-NHS (N-hydroxysulfosuccinimide, Pierce, A39269) and 5 mg/mL ethyl dimethylaminopropyl carbodiimide hydrochloride (EDC), then washed with 50 mM MES, pH 5.0, and incubated with 25 μg of antigen in 50 mM MES, pH 5.0 for 2 h on a rotator. The coupled beads were blocked in 5% BSA-PBS. After blocking, coupled beads were washed in PBS-Tween, resuspended in PBS, and stored at 4°C. Coupled beads were diluted to a concentration of 100 microspheres/μL and incubated with serum diluted in PBS at RT for 2 h, shaking at 800 rpm. For total IgG, IgG1, IgG2, IgG3, and IgG4 measurements, a 1:300 dilution was used. Two dilutions, 1:100 and 1:1000, were used in IgM measurements. Each sample was run in duplicate. The bound antigen-specific antibodies were subsequently probed with phycoerythrin (PE)-labeled antibodies detecting human IgG, IgG1, IgG2, IgG3, IgG4, or IgM (Southern Biotech). Fluorescence was analyzed using a Stratedigm S1000EON. The data are reported as the median fluorescence intensity of PE for a specific bead channel. Mean values of the duplicates are reported.

#### Antigen-specific Fc receptor binding

Plasma samples were analyzed using the antigen-specific Fc receptor binding multiplex assay (SeromYx Systems, Woburn, Massachusetts). Briefly, the Fc receptor Luminex array uses fluorescently coded microspheres to determine the ability of antigen-specific antibodies to interact with Fc receptors. MED20 recombinant protein (MyBioSource; MBS5308819) and a negative control antigen (Feline Immunodeficiency Virus core protein) were covalently coupled to Magplex Luminex beads via a two-step carbodiimide reaction and blocked in BSA-PBS, as described for antigen-specific antibody isotyping/subclassing. Coupled beads were diluted to a concentration of 100 microspheres/μL and incubated with serum diluted to 1:300 with PBS at RT for 2 h, shaking at 800 rpm. Each sample was run in duplicate. Bound antibodies were subsequently stained with PE-labeled tetramerized recombinant Fc receptors (FCGR2A 131H/R, FCGR2B, FCGR3A 158V/F, FCGR3B, FCAR) and C1q, for 1 h at RT, shaking at 800 rpm. Fluorescence was analyzed using a Stratedigm S1000EON. The data are reported as the median fluorescence intensity of PE for a specific bead channel. Mean values of the duplicates are reported.

#### Antibody-dependent cellular phagocytosis

Plasma samples were analyzed using the multiplex antibody-dependent cellular phagocytosis (ADCD) assay (SeromYx Systems, Woburn, Massachusetts). Briefly, antibody-dependent cellular phagocytosis assesses the ability of antibodies to induce phagocytosis of antigen-functionalized fluorescent beads by monocytes via Fc receptors. MED20 recombinant protein (MyBioSource; MBS5308819) was covalently linked to yellow-green fluorescent, carboxylate-modified polystyrene beads (Invitrogen, F8823) using the carbodiimide reagent EDC and amine-reactive Sulfo-NHS Ester. Coupled beads (10 µL/well) were added to 96-well plates with 10 µL/well of serum diluted to 1:80 with PBS and incubated for 4 h at 37 °C to form immune complexes. Each sample was run in duplicate. Following incubation, the immune complexes were washed with PBS, and THP-1 cells (1.5×10^4^ cells/well) were added and incubated for 16 h at 37 °C. After incubation, supernatant was removed, and cells were fixed with 4% PFA for 15 min. Fluorescence was acquired with a Stratedigm S1000EON. The data are reported as the average phagocytic score, which was calculated using the formula below:

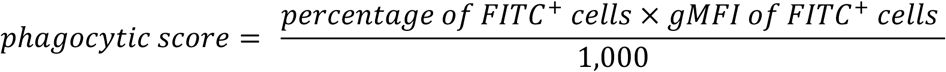

#### Antibody-dependent complement deposition

Plasma samples were analyzed using the multiplex antibody-dependent complement deposition (ADCD) assay (SeromYx Systems, Woburn, Massachusetts). Briefly, the multiplex antibody-dependent complement deposition assay assesses the recruitment of complement component C3b on the surface of antigen-coupled beads. MED20 recombinant protein (MyBioSource, MBS5308819) and a negative control antigen (Feline Immunodeficiency Virus core protein) were covalently coupled to Magplex Luminex beads via a two-step carbodiimide reaction and blocked in BSA-PBS, as described for antigen-specific antibody isotyping/subclassing. Coupled beads were diluted to a concentration of 100 microspheres/μL and incubated with serum diluted to 1:100 with PBS at RT for 2 h, shaking at 800 rpm. Each sample was run in duplicate. Commercially available guinea pig complement was then added as a source of complement (Sigma, G9774) and diluted 1:60 in Gelatin Veronal Buffered Saline (Boston BioProducts, IBB-300X). Following a 50-minute incubation at 37°C, the complement was washed away with 15mM EDTA in PBS, and a FITC-conjugated mAb specific for guinea pig C3b (MP Biomedicals, 0855385) was added for 20 min. Complement deposition was measured using a Stratedigm S1000EON and is reported as the geometric mean fluorescent intensity of FITC for a specific bead channel. Mean values of the duplicates are reported.

#### Anti-MED20 IgG Effector Function Comparison Across Groups

Fc effector function profiling data were analyzed in R (tidyverse, janitor, rstatix, ggpubr, patchwork). We focused on measurements of antibodies targeting the MED20 antigen and their ability to interact with Fc receptors. For each sample, we analyzed total IgG MED20 antibodies, the relative amounts of the four main IgG subclasses (IgG1–IgG4), binding to Fc gamma receptors (FcγRIIA-H, FcγRIIA-R, FcγRIIB, FcγRIIIA-V, FcγRIIIA-F, and FcγRIIIB), and functional readouts such as complement activation (ADCD), cellular phagocytosis (ADCP), and IgM levels measured. For each sample, summary metrics were calculated, including the proportion of each IgG subclass relative to total IgG, the IgM-to-IgG ratio, and an activating-to-inhibitory (A:I) index defined as the sum of activating FcγR binding signals (FcγRIIA-H + FcγRIIA-R + FcγRIIIA-V + FcγRIIIA-F + FcγRIIIB when present) divided by inhibitory FcγRIIB, which reflects the balance between immune-stimulating and immune-suppressing antibody signals based on Fc receptor binding. Selected features were log10-transformed prior to analysis. Group comparisons were performed for each feature using Kruskal–Wallis tests, with Benjamini–Hochberg (BH) correction applied across features to control the false discovery rate. Where significant, post hoc pairwise group comparisons were conducted using Dunn’s test with BH-adjusted q-values.

#### Anti-MED20 IgG effector function symptom analysis

Fc effector function profiling data were analyzed in R to identify associations between MED20-specific antibody features and clinical symptoms among LC participants. The same antibody features described above were used as continuous variables, and symptom variables were encoded as binary indicators. For each antibody–symptom pair, a point-biserial correlation was computed using Pearson correlation between the antibody signal (MFI) and the binary symptom variable, requiring a minimum of five complete sample pairs and variability in symptom status. Two-sided p-values were calculated for all tests and adjusted for multiple comparisons using the Benjamini–Hochberg procedure. Results were visualized using a dot plot in which dot color represents the point-biserial correlation coefficient (r), with blue indicating negative correlations and red indicating positive correlations; color intensity increases with absolute effect size (weak: |r| < 0.30; moderate: 0.30–0.59; strong: ≥ 0.60). Associations not reaching nominal significance (p ≥ 0.05) are shown in gray. Dot size reflects nominal p-value thresholds.

#### UTSW Autoantigen Protein Microarray

Autoantibody reactivities of IgM and IgG were examined at the Microarray Core facility at the University of Texas Southwestern Medical Center in Dallas, TX. Plasma samples were shipped on dry ice to the facility. A total of 120 human antigens, linked to various immune-related diseases or allergic conditions, were included. These encompassed ACE2, Aggrecan, Albumin, α Fodrin, Amyloid β(1-40), Amyloid β(1-42), AQP4, BAFF, BCOADC-E2, BPI, Calprotectin/S100, CD4, CD40, CENP-A, CENP-B, collagen I, II, III, IV, V, Complement C1q, C3, C4, C5, C6, C7, C8, C9, CRP, Cytochrome C, DFS70, dsDNA, EJ, Factors B, H, I, P, fibrinogen Type I-S, fibronectin, GAD65, GBM, genomic DNA, Gliadin, gp210, GP2, H/K-ATPase, histones H1, H2A, H2B, H3, HSPG, IA-2, IF, IFNγ, IL-6, IL-12/NKSF, IL-17A, Jo-1, KS, KU (P70/P80), La/SS-B, Laminin, LC1, LKM 1, LPS, lysozyme, M2, MBP, MDA5, Mi-2, mitochondrion, MPO, myosin, Nrp1, nucleolin, nucleosome, Nup62, NXP2, OGDC-E2, P0, P1, P2, PCNA, PDC-E2, PL-7, PL-12, PM/Scl-75, PM/Scl 100, PR3, proteoglycan, prothrombin, Ro/SS-A(52 kDa), Ro/SS-A(60 Kda), SAE1/SAE2, Scl-70, SLA/LP, Sm, Sm/RNP, SmD, SmD1, SmD2, SmD3, SP100, SRP54, ssDNA, Tau, thyroglobulin, TIF1γ, TLR4, TNFα, TPO, tTG, U1-snRNP 68/70kDa, U1-snRNP A, U1-snRNP C, U-snRNP B/B’, Vimentin, Vitronectin, and ß2-Glycoprotein 1), were printed on 16-pad nitrocellulose FAST slides. Eight positive control proteins (human IgM and IgG, anti-human IgM and IgG) were also included as controls. Briefly, plasma samples treated with DNAse I (Thermo Fisher, #AM2222) were applied to the arrays at a 1:50 dilution. Binding was detected with cy3-labeled anti-human IgG antibodies (Jackson ImmunoResearch Laboratories, #IgG-109-166-098). The slides were scanned at 532 nm and 635 nm wavelengths using a Genepix 4400A scanner. Image analysis was performed with Genepix Pro v7.0 software (Molecular Devices) to generate report files (GPR). The net signal intensity (NSI) for each antigen was obtained by subtracting negative control values. Normalized NSI values, using the positive controls, were used for case-control comparisons. Signal-to-noise ratios (SNR) were calculated with the formula: (foreground median – background median)/standard deviation. Antigens with SNR<3 in over 10% of samples were excluded. The mean plus 2 Standard Deviations (SD) from the Healthy Control group was used as the threshold for positivity.

#### GPCR and Ionotropic Receptor Autoantibody Profiling by ELISA

IgG AAB against Angiotensin II Type 1 Receptor, Angiotensin II Type 2 Receptor, Endothelin A Receptor, Endothelin B Receptor, Alpha-1 Adrenegic Receptor, Alpha-2A Adrenergic Receptor, Alpha 2B Adrenergic Receptor, Alpha-2C Adrenergic Receptor, Beta-1 Adrenegic Receptor, Beta-2 Adrenegic Receptor, Muscarinic Acetylcholine Receptor M3, Muscarinic Acetylcholine Receptor M4, Nicotinic Acetylcholine Receptor Alpha 1, Nicotinic Acetylcholine Receptor Alpha 2, Nicotinic Acetylcholine Receptor Alpha 7, Protease Activated Receptor 1, Stabilin-1 Scavenger Receptor, Serotonin 5-HT2A Receptor, GLP1, GLP2, NMDAR1, NMDAR2b, NMDAR2c were measured using respective sandwich ELISA kits by CellTrend GmbH (Luckenwalde, Germany) in an EN ISO250 certified laboratory (CellTrend). In brief, serum samples were diluted at a 1:100 ratio and AAB levels were calculated as units per mL by extrapolating from the standard curve. The ELISA kits were validated in accordance with the Food and Drug Administration’s Guidance for Industry: Bioanalytical Method Validation and intraassay coefficient of variation ranged from 3.9% to 15.2% depending on the respective assay. Long COVID AAB levels were compared to internal reference based on normal values of healthy individuals.

#### Passive transfer and behavior analysis

Mice were injected intraperitoneally with 38.4 mg/kg of total IgG from participants (Healthy Controls, Convalescent Controls, and LC) or with PBS. One individual donor IgG per mouse was used. Mice were co-housed, with one mouse per cage for each experimental condition. All the experiments were performed between 1 and 6 PM, and mice were allowed to acclimate in the room for at least 1 h.

#### Open Field Test

The Open Field test was performed to characterize locomotor activity and anxiety-like behaviors. The apparatus consisted of a 50 cm × 50 cm arena surrounded by 45 cm opaque plexiglass walls. For testing, the mice were placed in the center of the arena and left to explore for 10 min, after which they were returned to their home cage. EthoVision XT 15 software (Noldus, Wageningen, the Netherlands) was used to track distance traveled in central and peripheral zones, the ratio of time spent in central and peripheral zones, movement, distance traveled, activity, and velocity.

#### Elevated Zero Maze Test

Elevated Zero Maze was administered to measure anxiety-like behaviors and behavioral disinhibition. The apparatus consisted of an O-ring arena elevated 60 cm above the ground, with two opposing arms enclosed in plexiglass walls (50 cm × 10 cm × 40 cm) and two opposing open arms (50 cm × 10 cm). Mice were placed in the open arm and left to explore the arena for 5 min, after which they were returned to their home cage. EthoVision XT 15 software was used to track movement, velocity, and time spent in the closed and open arms.

#### Rotarod

To assess the motor coordination of the mice, an accelerated rotarod test was performed. On test day, mice were placed on the resting rotarod apparatus (Ugo Basile, Gemonio, Italy) for at least 1 min. The speed of the rotarod was accelerated from 4 to 80 rpm over a 300 s period. The mice were subjected to four trials with at least 15 min intervals between trials. The retention time of the rod in each trial was recorded, and the average of all the trials was plotted.

#### Grip Strength Test

Limb strength (fore and hind limb) was assessed using a commercially available grip strength meter (Ugo Basile), incorporating a stainless-steel orientable grid and force transducer. The mice were held gently by the base of the tail and pulled across the horizontal orientable grid so that they were able to grip the bar with their four paws. A digital readout of the maximum force applied is given once the grip is released. The test was performed at least three times on each mouse to measure force values.

#### Hot Plate test

To evaluate thermal sensitivity, mice were placed on a hot plate (Ugo Basile) with the plate set at 55 °C. The time to the first sign of nociception (i.e., paw licking, flinching, or jump response) was measured, and the animal was immediately removed from the hot plate. A cut-off period of 20 s was maintained to avoid damage to the paws.

#### Blood Pressure and Heart Rate

Measurement of mouse blood pressure was carried out using the CODA mouse tail-cuff blood pressure system (Kent Scientific Corp., Torrington, CT, US). Measurements were made during the afternoon (1–6 pm) with the mice awake, restrained on the CODA apparatus, and lying down on a warm plate, covered with a dark blanket. Each blood pressure measurement was carried out for 20 cycles for acclimatization and 20 cycles with recording, with a deflation time of 20 s. The mean blood pressure, systolic blood pressure, diastolic blood pressure, and heart rate were determined by the average of all 20 measurements during the recording cycles.

#### Treadmill

For the treadmill analysis, the electricity intensity of the shock grid was set to 0.3 mA. If a mouse remained in the fatigue zone (i.e., on the shock grid) for five continuous seconds, the electric stimulation was automatically stopped, and the mouse is promptly removed from the treadmill. The total running duration and distance were recorded. On the training day, mice were first allowed to explore freely for 3 min at a speed of 0.1 cm/s, followed by 7 min at 10 cm/s, for a total training time of 10 min. Mice were then returned to their home cage. On the second day, the mice were placed on the treadmill for a ramped speed test: 10 cm/s for 10 min for warm up and followed by a sequential increases to 15 cm/sc (5 min), 20 cm/s (3 min), 25 cm/s (3 min), 30 cm/s (3 min), 35 cm/s (3 min), 40 cm/s (3 min), 50 cm/s (3 min), 60 cm/s (3 min), and finally 70 cm/s maintained until the mice reach fatigue. Fatigue-like behavior was defined as remaining in the fatigue zone (shock grid) for 5 consecutive seconds. After the ramp test, mice were returned to their home cages and allowed to rest for one whole day, during which hIgG injection was performed. On the following days, mice were subjected to the same ramp-speed treadmill protocol to assess fatigue-like behavior.

#### iDISCO+ and ClearMap analysis in the brain samples

Whole-brain staining was performed following the iDISCO+ protocol previously described^21^. Briefly, perfused brains were dehydrated in methanol 20%, 40%, 60%, 80% and 100% (Sigma-Aldrich) and incubated ON in a 66% dichloromethane solution (Sigma-Aldrich) in methanol. Samples were bleached ON at 4 °C in methanol containing 5% hydrogen peroxide (Sigma-Aldrich) and then rehydrated after incubation in methanol at 60%, 40%, and 20% concentrations. After permeabilization, samples were incubated with rabbit c-FOS antibody, 1:4000 (Synaptic Systems 226 008) at 37 °C for 10 days. Secondary antibodies conjugated to Alexa 546 were used (Life Technologies). After a final incubation in methanol for dehydration, brain samples were washed in dichloromethane 100% for clearing and stored in dibenzyl ether (both from Sigma-Aldrich). The imaging acquisition was performed at the Yale West Campus Imaging Core in a light sheet microscope (LaVision BLAZE) equipped with a sCMOS camera and an LVMI-Fluor x4 objective lens with a 6 mm working distance dipping cap. Images were taken every 6 μm (z steps), and the numerical aperture was set to 0.03NA. Reconstructed images were further processed with ClearMap2 software (https://github.com/ChristophKirst/).

#### Modified iDISCO+ and IMARIS analysis in the paw samples

After perfusion, hindpaws were dissected out, and post-fixed in PBS/0.5% PFA/10% sucrose at 4 °C ON and then washed with PBS at RT for 1 h three times. The tissues were then cleared following the iDISCO+ protocol with the following modifications: after dehydration, bleaching, and rehydration, the tissues were decalcified in 350 mM EDTA-Na (pH 6.5) at 37 °C for 72 h, with the fresh buffer changed every 24 h. The tissues were immunolabeled with the rabbit anti-PGP9.5 antibody (Proteintech Group, Cat#14730-1-AP, RRID: AB_2210497) diluted (1:250) in PBS/0.2% Tween-20/10 μg/mL heparin/5% normal donkey serum/25 mM EDTA (pH 6.5) at 37 °C for 96 h. Secondary antibody conjugated to Alexa 647 was used (Life Technologies). Imaging acquisition was performed on the light sheet microscope with the same settings for brain samples. Images were converted (tif to .ims) and imported to IMARIS 10.0.1 (Oxford Instruments) for 3D reconstruction. PGP9.5 isosurface, as well as the autofluorescence isosurface (which represents the entire hindpaw structure), were thresholded and constructed, and the total volume of these isosurfaces was calculated with IMARIS built-in functions. Axon volume is quantified as the total PGP9.5 isosurface volume normalized by autofluorescence isosurface volume. The statistics were performed with the Kruskal-Wallis test followed by Dunn’s multiple comparison^82^ test, which is built into GraphPad Prism 9.3.1.

#### Mouse necropsy and histopathology analysis

All necropsies, histopathological analyses, and complete blood counts were performed by Yale Comparative Pathology Research Core by a certified pathologist in a blinded manner. Five days post-injection of hIgG, 7-week-old male and female mice were transcardiacally perfused with PBS containing heparin (20 IU/mL), followed by 10% neutral buffered formalin. Whole-body necropsies were performed immediately following euthanasia. Major organs were carefully dissected and examined macroscopically for signs of gross pathology. Tissues were postfixed ON in 10% neutral buffered formalin, then embedded in paraffin, and sectioned at 5 μm thickness. Tissue sections were stained with hematoxylin and eosin (H&E) for general histopathological assessment. Slides were evaluated by a blinded pathologist (C.J.B.) using light microscopy. Histopathological features were scored semi-quantitatively based on pre-defined criteria. All histological images were captured by K.S.G.S using BioTek Cytation 5 imaging reader (Agilent).

#### IEFN Skin biopsy analysis

Mice were transcardiacally perfused with PBS containing heparin (20 IU/mL), followed by 4% paraformaldehyde in PBS. Paws were then postfixed ON in paraformaldehyde at 4 °C, followed by successive ON incubations in PBS containing 15% sucrose and then 30% sucrose. All specimens were further processed by personnel blind to the treatment groups for all subsequent steps. Glabrous skin from the hind paw was cut from the phalanges, embedded in OCT (Sakura Finetek, 4583), and cut into 50 µm thick sections on a cryostat along a plane perpendicular to the long axis of the digit. Sections were mounted on SuperFrost™ PLUS Adhesion Slide (Electron Microscopy Sciences, 71869-11); each slide contained specimens from multiple treatment groups. Slides were baked 30 min at 60 °C, then blocked at RT for 1 h in PBS with 1% BSA, 10% goat serum, and 0.3% Triton X-100. Specimens were stained ON at 4 °C in PBS containing 1% BSA, 0.3% Triton X-100, a 1:250 dilution of rabbit anti-PGP9.5 (Abcam, ab108986), and a 1:500 dilution of Alexa 488-conjugated donkey anti-human IgG (Jackson ImmunoResearch, 709-545-149). Slides were washed in PBS with 0.1% Tween-20 and incubated 1h at RT in a 1:500 dilution of Alexa 594-conjugated goat anti-rabbit IgG (Jackson ImmunoResearch, 111-585-144). After final washing in PBS/Tween-20, nuclei were stained with DAPI, and coverslips were mounted on slides with Prolong Gold (Cell Signaling Technology, 9071S). Images were collected through a 40×/1.3NA oil-immersion objective on a Stellaris confocal microscope using identical power and gain settings across treatment groups within a given experiment. PGP9.5-positive intraepidermal nerve fibers (IENF) crossing the basement membrane dividing the dermal layer from the stratum spinosum of the epidermis were counted on each section and normalized to the length of the basement membrane across the section, in accord with diagnostic criteria for small fiber neuropathy diagnosis by skin biopsy^83^.

#### Longitudinal IENF intravital imaging

IENF were visualized with heterozygous transgenic mice offspring of the Cre driver line B6.129(Cg)-*Scn10atm2(cre)Jwo*/TjpJ^84^ (a gift from John Wood, University College London) and the Cre-dependent tdTomato reporter line B6.Cg-*Gt(ROSA)26Sortm14(CAG-tdTomato)Hze*/J (Jackson Labs, Bar Harbor, 007914). These animals exhibit tdTomato fluorescence in Scn9a-expressing cells, which in the epidermis consist only of the delicate nerve fibers of nociceptive sensory neurons^84^. For longitudinal imaging, mice were anesthetized with an intraperitoneal injection of 117 mg/kg ketamine and 13 mg/kg xylazine, and the distal joint of the hindpaw third digit was attached to a coverslip mounted into a specimen holder (Narishige, CP-1). Intravital imaging was conducted on a two-photon microscope (Miltenyi Biotec, Cologne, Germany) equipped with a Ti:Sapphire pulsed laser (Coherent, Saxonburg, PA) tuned to 920 nm and supplying irradiance of ∼30mW/mm^2^ post-objective. Fluorescence of tdTomato was collected through a 615±10nm filter, and the fluorescence of endogenous collagen in the dermis produced by second harmonic generation^85^ was collected with a 435±90 filter—z-stacks collected through a 25×/1.1NA water-immersion objective (Nikon). Animals were examined 24h before treatment with purified IgG samples (38.4 mg/kg, i.p.) and then re-examined at 1, 2, 4, 8, and 15 days post administration.

Images were preprocessed to remove any dead pixels with the Remove Outliers function within ImageJ. For a given animal, images across sessions were then registered to a common coordinate space using a Python implementation of the simple Insight Tool kit (simpleITK). A standard volume present across all samples for an animal was then analyzed for IENF by applying the Triangle threshold algorithm to both fluorescence channels. Voxels positive for tdTomato fluorescence and negative for collagen signal served as seed points for the label spreading algorithm in the scikit-learn Python library. This approach results in specific segmentation of nociceptor fibers within the epidermis, which then allows for determining the number of fibers as well as their total volume. The dermal:epidermal junction was extracted from volumetric data by first applying a 50-pixel Gaussian filter on the thresholded collagen image, then defining the voxels closest to the epidermis that still retained collagen signal. This yielded an irregular surface representing the basement membrane; the thresholded nociceptor fiber signals 10 microns above each point on this surface were then analyzed by the scikit-image Python library to identify connected components. Counting these objects produced the total number of nociceptor fibers that had crossed the threshold defined as the shallowest extent of collagen signal in the image volume.

#### Cytokines and neurotransmitters measurement

Three days post-injection hIgG, 6–8 weeks old female mice underwent transcardially perfusion with PBS containing heparin (20 IU/mL). Whole brains were carefully dissected, weighed, and homogenized using lysing matrix D tubes (MP Biomedicals, 116913500) and FastPrep-24 homogenizer (MP Biomedical). Five days post-injection of IgG, 6-8 weeks old female mice were anesthetized with 20% (w/v) isoflurane in propylene glycol, administered in a certified ducted hood. The absence of a response to toe pinch confirmed adequate anesthesia. Retro-orbital blood was then collected using Microvette lithium heparin tubes (Microvette, 20.1282.100) and kept on ice until processed. Samples were centrifuged at 5,000 g for 10 min at 4 °C, plasma was aliquoted and stored at –80 °C for downstream analysis.

Neurofilament-L was measured in plasma using Pathscan® Total Neurofilament-L Sandwich ELISA Kit (Cell Signaling Technology, 99175C), following the manufacturer’s instructions.

Cytokines, including IFN, IL-12, MCP-1, TNF, IL-6, and IL-10, were measured on brain homogenate and plasma using BD CBA Mouse Inflammation Kit (BD, 552364), following the manufacturer’s instructions. Neurotransmitters were quantified in brain homogenate using ELISA kits from ImmuSmol, according to the manufacturer’s protocol: Glutamate (Glutamate ELISA kit Any sample, BA-E-2400), Gaba (GABA ELISA kit Any sample, BA-E-2500), Serotonin (Serotonin ELISA kit Ultra-Sensitive – Any sample, BA-E-5900R), Dopamine (Dopamine ELISA kit – Ultra-Sensitive Any sample, BA-E-5300R).

## QUANTIFICATION AND STATISTICAL ANALYSIS

### Linear Regression Analysis of Autoantibody Tissue Reactivity for Immunofluorescence Adjusted for Age and Sex

To evaluate differences in autoantibody intensity across clinical groups, we performed linear regression modeling on immunofluorescence (IF) data obtained from tissue-based autoantibody staining. Autoantibody mean of fluorescence intensity was modeled using linear regression for each tissue separately, with group status HC, CVC, or LC as the primary predictor and age and sex included as covariates. Two sets of models were fit using either HC or CVC as the reference group. For each tissue, we extracted the estimated group effect, 95% confidence intervals, and associated p-values. P-values were adjusted for multiple testing using the Benjamini–Hochberg False Discovery Rate (FDR) method, and associations with adjusted p-value < 0.1 were considered significant. Forest plots were generated to visualize effect sizes and confidence intervals for each group comparison (LC vs. HC, CVC vs. HC, LC vs. CVC), highlighting statistically significant differences.

### Association of Tissue-Specific IF Positivity with Clinical Symptoms in Long COVID

To assess the relationship between tissue-specific autoantibody immunofluorescence (IF) reactivity and symptom profiles in LC, we analyzed the IF mean of fluorescence intensities, symptom annotations, and demographic metadata. For each tissue and binary symptom pair, we computed a positivity threshold based on healthy controls (HC), defined as the tissue-specific mean + 1 SD of IF signal across HC samples. Samples exceeding this threshold were labeled as IF-positive. We then stratified LC samples by their symptom status (positive vs. negative) and evaluated the co-occurrence of IF and symptom positivity.

For each tissue–symptom pair, we generated a 2×2 contingency table comparing IF and symptom status and conducted a Fisher’s exact test to evaluate enrichment (implemented with fisher.test() in R). Only comparisons with ≥10 LC samples and at least one observation in each category were included. Results were visualized as grouped bar plots showing the proportion of symptom-positive and -negative individuals within IF-positive groups.

Additionally, for each tissue–symptom combination with sufficient sample size (n ≥ 6 per group), we performed a two-sided Wilcoxon rank-sum test comparing the mean IF signal between symptom-positive and symptom-negative patients. P-values were adjusted for multiple comparisons using the Benjamini–Hochberg false discovery rate (FDR) method. Results were visualized via dot plots for each tissue, showing the effect size (difference in mean IF signal between groups) and significance level. Summary plots were generated to highlight symptom-tissue associations with FDR-corrected p-values, using color and size scales corresponding to −log10(adjusted p-value).

All analyses were performed in RStudio using the tidyverse, janitor, scales, and ggplot2 packages.

### Immunofluorescence ROC analysis

Using the mean MFI from HC + standard deviation (SD) of HC as a threshold, we classified LC and non-LC samples (HC + CVC combined) as predicted positive or negative within each tissue, and derived true positives (TP), false negatives (FN), true negatives (TN), and false positives (FP). Sensitivity and specificity were then calculated and 95% confidence intervals for these proportions were obtained using Wilson’s binomial method (DescTools::BinomCI).

To evaluate the overall discriminatory performance of IF intensity independent of a single cutoff, we performed receiver operating characteristic (ROC) analysis separately for each tissue–marker combination using the pROC package in R. For each subset, LC status (LC vs non-LC) was used as the binary outcome and raw IF intensity as the predictor. ROC curves were fit with LC as the positive class; if the initial area under the curve (AUC) was <0.5, the predictor was inverted to ensure that higher values corresponded to higher LC probability. AUC values were computed along with 95% confidence intervals using DeLong’s method, from which standard errors were derived. Z-statistics were used to test whether AUC values differed significantly from 0.5 (random classification), and corresponding p-values were reported.

For visualization, ROC curves were plotted for each tissue–marker pair with specificity on the x-axis and sensitivity on the y-axis, including a diagonal reference line representing random performance. The summary panel for each ROC plot was annotated with the AUC, its standard error, 95% confidence interval, and p-value. The performance of the pre-specified HC mean + SD threshold was overlaid as a single point on the ROC curve to facilitate comparison between this biologically motivated cutoff and the full threshold-independent ROC performance.

### Image Processing For Colocalization Quantification

Immunofluorescence images of meninges were acquired as .tif hyperstacks and processed using Fiji/ImageJ (NIH). Prior to colocalization analysis, channels of interest were duplicated and converted to 16-bit format to standardize intensity scaling. Background subtraction was performed using a rolling-ball algorithm (radius = 50 pixels), followed by Gaussian smoothing (σ = 2) to reduce high-frequency noise and improve signal-to-noise ratio. These preprocessing steps were applied uniformly across all samples to ensure comparability. Regions of interest (ROIs) were defined to restrict analysis to tissue-containing areas of each image. Colocalization analysis was performed using the Coloc 2 plugin in Fiji/ImageJ. For each image, the hIgG channel was analyzed independently against the NG2 and CD31 channels. Colocalization thresholds were determined using Costes’ automatic threshold regression method. Manders’ overlap coefficient tM2 was used as the primary metric for downstream analysis. Specifically, tM2 represents the fraction of thresholded signal intensity in the hIgG channel that overlaps with signal in the comparison channel (NG2 or CD31). To facilitate interpretation, tM2 values were stratified into predefined overlap categories: minimal overlap (0–0.3), moderate overlap (0.3–0.5), high overlap (0.5–0.7), and very high overlap (0.7–1.0). These categories were used for visualization and comparison across samples.

### GPCR and Ionotropic Receptor Autoantibody Group Comparison

GPCR and Ionotropic Receptor autoantibody data were analyzed in R (tidyverse, janitor, rstatix, ggpubr, patchwork). Positivity thresholds were defined for each antibody as the healthy control mean plus two standard deviations (mean + 2×SD), and samples exceeding this threshold were classified as positive. Group differences in antibody levels were tested using Kruskal–Wallis tests followed by false discovery rate (FDR) correction using the Benjamini–Hochberg method. For antibodies with significant overall group effects, post hoc pairwise comparisons between groups were performed using Dunn’s test with FDR-adjusted p-values. For Long COVID participants, the number of positive GPCR antibodies per individual was calculated to quantify overall antibody burden and visualized using a bar plot.

### ROC analysis of GPCR and Ionotropic Receptor autoantibodies

We analyzed a prespecified panel of GPCR and Ionotropic Receptor related autoantibodies measured by ELISA. Using the mean from HC + standard deviation (SD) of HC as a threshold, samples were classified as predict positive or predict negative. True positives (TP), false negatives (FN), true negatives (TN), and false positives (FP) were then computed using LC status (LC = positive; HC + CVC = negative). Sensitivity and specificity were then calculated and 95% confidence intervals for these proportions were obtained using Wilson’s binomial method (DescTools::BinomCI).

To evaluate discrimination independent of a fixed cutoff, we performed receiver operating characteristic (ROC) analysis for each antibody using the proc package in R. LC status (LC vs. non-LC [HC + CVC]) was used as the binary outcome, and the antibody MFI values were used as predictors. ROC curves were generated with LC as the positive class. When the initial AUC was <0.5, predictor directionality was inverted to ensure that higher antibody values reflected stronger association with LC. The Area Under The Roc Curve (AUC) and its 95% CI were calculated using DeLong’s method. Standard errors were estimated from CI width, and z-statistics were used to test whether each AUC differed significantly from 0.5 (random classification). P-values were computed using both normal approximation (Prism-style) and DeLong’s one-sample test.

For visualization, ROC curves were plotted for each tissue–marker pair with specificity on the x-axis and sensitivity on the y-axis, including a diagonal reference line representing random performance. The summary panel for each ROC plot was annotated with the AUC, its standard error, 95% confidence interval, and p-value. The performance of the pre-specified HC mean + SD threshold was overlaid as a single point on the ROC curve to facilitate comparison between this biologically motivated cutoff and the full threshold-independent ROC performance.

### GPCR t-SNE analysis

GPCR autoantibody reactivity data were analyzed in R using the tidyverse, janitor, stringr, Rtsne, cluster, factoextra, and vegan packages. To normalize feature values across the cohort, HC-referenced z scores were computed independently for each GPCR feature. A threshold of z > 2 was used to define elevated GPCR reactivity. For unsupervised analysis, only LC samples were retained. Two-dimensional embeddings were generated using t-distributed stochastic neighbor embedding (t-SNE; Rtsne package). Clustering was performed on the t-SNE embedding using k-means (k = 2, seed = 123). Cluster centroids were defined as the mean coordinates of samples within each cluster. For each feature, the mean z score within each cluster was calculated, and the top 10 features per cluster (ranked by mean z score) were defined as cluster-driving GPCR features. Cluster structure was evaluated using multiple complementary approaches. Silhouette analysis was performed using Euclidean distances computed from the t-SNE coordinates, and the mean silhouette width was used to quantify cluster separation. In addition, a permutation-based multivariate analysis of variance (adonis2; 999 permutations, Euclidean distance) was used to test whether cluster assignment explained variation in the embedding. For visualization, LC samples were plotted in t-SNE space and colored by cluster assignment. Cluster labels were positioned at centroid coordinates.

### Heatmap visualization of cluster-driving GPCR features

To visualize GPCR features associated with cluster identity, heatmaps were generated using HC-referenced z-score data from Long COVID (LC) samples. Analyses were performed in R using the tidyverse, pheatmap, and RColorBrewer packages. Cluster-driving GPCR features were defined based on their mean z score within each cluster, as previously described. For heatmap visualization, the top features per cluster (based on mean z score) were selected proportionally to ensure equal representation across clusters, with the total number of features capped (typically 20). Alternatively, features were ranked globally based on their maximum mean z score across clusters, and the top features were selected irrespective of cluster origin. Heatmaps were generated using the pheatmap package with row-wise scaling (z-score normalization per feature) to emphasize relative differences in GPCR reactivity across samples. Hierarchical clustering was applied to both rows and columns using default distance metrics and linkage methods.

### Cell-type enrichment analysis of cluster-associated GPCR targets

To investigate the cellular context of cluster-associated GPCR autoantibody targets, enrichment analyses were performed using single-cell RNA expression data from the Human Protein Atlas (HPA). All analyses were conducted in R using the tidyverse, janitor, stringr, and forcats packages. HPA single-cell expression data were filtered to include only genes with detectable expression (nCPM ≥ 1) within each cell type. A gene was considered associated with a given cell type if it was expressed above this threshold in that cell type. Cluster-specific gene sets were defined by aggregating unique gene symbols corresponding to cluster-driving GPCR features for each cluster. Analyses were restricted to comparisons between two clusters. For each cell type, enrichment was evaluated by constructing a contingency table comparing the presence or absence of cluster-associated genes within that cell type across the two clusters. Specifically, counts were defined as: (a) genes from cluster 1 present in the cell type, (b) genes from cluster 1 absent from the cell type, (c) genes from cluster 2 present in the cell type, and (d) genes from cluster 2 absent from the cell type. Odds ratios were calculated using a continuity-corrected estimator to account for zero counts, and log2-transformed odds ratios were used to quantify the direction and magnitude of enrichment. Positive log2 odds ratios indicated enrichment of cluster 1-associated genes in a given cell type, whereas negative values indicated enrichment of cluster 2-associated genes. Cell types were ranked based on log2 odds ratios, and the top enriched cell types for each cluster were identified by selecting the highest and lowest values. Results were visualized using scatter plots of log2 odds ratios, with cell types displayed along the y-axis and enrichment magnitude along the x-axis. Points were colored according to the cluster in which enrichment was observed.

### HuProt Data Preprocessing and Batch Correction

For the preprocessing of HuProt data, the signal intensities were log_2_-transformed (log_2_MFI). The dataset included samples from three batches with healthy controls (HC), convalescent controls (CVC), and long COVID (LC) groups. To preserve biological variation across groups while correcting for batch effects, we used the empirical Bayes ComBat method implemented in the sva R package ^86^. Covariates included age, sex (encoded as binary), and clinical status (HC or CVC and LC). A model matrix was created as ∼ age + sex + status. The batch with more samples (batch_2), containing bridge samples, was selected as the reference batch.

To evaluate batch-related variance before and after ComBat correction, we performed principal component analysis (PCA) using the FactoMineR^87^ package and visualized the results with factoextra. To evaluate reproducibility of duplicate bridge samples across batches, we performed Bland–Altman analysis, calculating mean bias, standard deviation of the differences, and 95% limits of agreement. The results showed low mean bias (ranging from ∼0.01 to 0.96) with differences distributed within the calculated limits of agreement, supporting high reproducibility across runs. Samples were colored by batch, and confidence ellipses were drawn to assess batch clustering. We further quantified residual batch effects using PERMANOVA (adonis2 from the vegan package), with Euclidean distances computed on the corrected expression matrix and batch label used as the independent variable. After correction, PERMANOVA test results were: R^2^ = 0.0102 with F = 0.8235 and p-value = 0.624.

For downstream threshold analysis, Z-scores were computed per sample using the healthy controls as the reference distribution. Z-scores were calculated for Long COVID, convalescent controls, and healthy controls. To estimate autoantibody burden, we applied Z-score thresholds of 3 (per manufacture’s suggestion – CDI labs) and counted the number of proteins exceeding each threshold per sample. These counts were summarized across samples by group, and group-level means and standard errors were visualized.

For each protein, the mean and SD across HC samples were computed, and Z-scores were calculated per sample. Proteins with Z-scores > 3 were considered positive hits. Private hits were defined as proteins that were positive in one or more LC samples but not in any CVC or HC samples.

All analyses were performed in RStudio version 2023.06.0+421.

### Pathway Enrichment Analysis of Private and Positive HuProt Hits for Long COVID

To investigate the biological pathways associated with positive and private autoantibody targets in LC samples, we performed gene set enrichment analyses using unique gene symbols from HuProt hits and mapped to Entrez Gene IDs using the *clusterProfiler* R package and the *org. Hs.eg.db* annotation database. Successfully mapped genes were utilized for overrepresentation analysis across Gene Ontology Biological Process (GO-BP). GO-BP enrichment was performed using the enrichGO() function with the whole human Entrez gene universe, applying Benjamini– Hochberg false discovery rate (FDR) threshold of 0.1. Results were visualized using custom dot plots generated from the top 15 significantly enriched terms. Neurological-related pathways were further filtered based on keyword pattern matching (e.g., “neuro”, “synap”, “brain”), and enriched terms were visualized separately. Neurological gene members were extracted from the GO-BP results and exported for downstream analyses.

### Autoantibody Target Tissue and Cell Annotation in Long COVID for HuProt data

To investigate the tissue and cell type specificity of positive and private autoantibody responses in LC, gene symbols corresponding to each protein were mapped using a reference annotation file, and private hits were cross-referenced with the Human Protein Atlas (HPA) metadata, which includes tissue and single-cell RNA expression specificity (TPM) annotations.

For each gene, tissue, and cell type, specificity was parsed from HPA metadata (values > 0 TPM only), and counts were generated to identify the most frequently targeted tissues and cell types across the cohort. Bar plots and per-tissue/per-cell type plots were generated to visualize the top targeted tissues and cells. Gene-level positive hit frequencies were also computed across LC patients and annotated with tissue and cell-type information to assess common targets. The number of autoantibodies that target neurological tissues (defined as those with expression in brain, retina, choroid plexus, or pituitary gland) was quantified per patient, and both private and positive hits were analyzed by group. Pairwise comparisons of tissue-level reactivity between groups were conducted using Wilcoxon rank-sum tests, followed by Benjamini-Hochberg correction for multiple testing. Dot plots were generated to visualize group-wise differences per tissue, with circle size representing the statistical significance (−log₁₀ adjusted *p*-value) and color indicating the direction and magnitude of difference in mean tissue reactivity. All analyses were performed in RStudio version 2023.06.0+421 with the tidyverse, rstatix, viridis, and ggpubr packages.

### Autoantibody Heatmap Generation and Neurological Proteins Annotation Analysis

To visualize the top autoantibodies that target neurological tissues across participant groups, we identified proteins associated with neurological processes by retrieving Gene Ontology (GO) terms related to neurological function (e.g., neurogenesis, synaptic signaling) and extracted associated gene symbols using the org.Hs.eg.db and GO.db packages. For each protein associated with a neurological process, we calculated the mean z-score per sample. Proteins were then ranked by the total summed z-score across LC samples, and the top 50 most reactive protein targets were selected for visualization. The final heatmap was generated using the pheatmap package in R, displaying standardized autoantibody reactivity. Samples were grouped by clinical status.

### t-SNE and Cluster Analysis of Neurological Autoantibody Signatures

To investigate groups of neurological autoantibody profiles among Long COVID (LC) patients, we performed t-distributed stochastic neighbor embedding (t-SNE) using z-score normalized IgG reactivity values derived from HuProt protein array data. LC samples were embedded in two-dimensional space using the Rtsne package (perplexity selected based on sample count), applied to protein-level and neuro-tissue-level expression matrices of the proteins. K-means clustering was performed on the resulting t-SNE coordinates to identify distinct patient subgroups. Features enriched in each cluster were determined by comparing average z-scores per feature within each cluster, and the top 20 cluster-driving features were visualized using barplots. Additional plots were generated by overlaying average expression across top features onto the t-SNE map, with cluster centroids annotated. Analyses were performed using custom R scripts incorporating tidyverse, AnnotationDbi, and ggplot2.

### Heatmap Visualization of Cluster-Defining Features

To visualize the expression patterns of key features driving patient clustering, a heatmap was generated using the top 40 cluster-defining proteins. Sample clustering assignments were previously defined from t-SNE analysis. The final heatmap was constructed using the pheatmap package in R. Expression values were normalized by Z-score using the HC group as a reference, and both samples and proteins were hierarchically clustered.

### Correlation of Positive and Private Long COVID Autoantibodies with Symptom and Pain Phenotypes

To investigate the clinical relevance of positive and private LC autoantibody targets, we analyzed associations between HuProt and mass spec data hits and patient-reported symptoms and neuropathic pain phenotypes. Only LC samples were retained for symptom analysis. Symptoms were filtered to include only those with at least 3 positive and 3 negative cases to ensure statistical robustness. Point-biserial correlations were computed between positive and private LC protein intensities (continuous) and binary symptom variables (0/1) using Pearson’s correlation test. Resulting p-values were adjusted for multiple comparisons using the Benjamini-Hochberg FDR method. Significant associations (FDR < 0.1) were visualized in a dot plot, with effect sizes represented as –log₁₀(p-value) and color-coded by significance. In parallel, we assessed whether private LC proteins were associated with neuropathic pain subtypes defined by the SLANSS (Self-Report Leeds Assessment of Neuropathic Symptoms and Signs) classification. LC samples with available SLANSS pain category were grouped, and protein expression values were compared across SLANSS groups using the Kruskal-Wallis test. For each protein, group-level means were computed, and test statistics were used to identify significant differential expression. Results were visualized via a dot plot stratified by SLANSS category, with point size and color encoding statistical significance and p-value strength.

### HuProt autoantibody burden ROC analyses

To evaluate whether the overall autoantibody burden or LC-private antibodies could discriminate LC from control samples, we performed receiver operating characteristic (ROC) analysis using the PROC package in R, restricted to LC cases and controls (HC + CVC). Neuro-associated proteins were defined based on Gene Ontology (GO) terms related to nervous system biology, including *nervous system development* (GO:0007399), *central nervous system development* (GO:0007417), and *synaptic transmission* (GO:0007268). GO term–to–Entrez ID mappings and Entrez-to-gene symbol conversions were performed using ORG.HS.EG.DB, and HuProt probes whose mapped gene symbols overlapped this list were annotated as “neuro” proteins.

For each sample, several burden metrics were derived: (i) the total number of positive proteins across the entire array (n_pos_all) and (ii) the number of positive proteins restricted to GO-defined neuro-associated proteins (n_pos_neuro). LC-private proteins were defined as proteins that were positive in at least one LC sample and negative in all controls (HC + CVC). From this set, we calculated (iii) the total number of private LC hits per sample (n_priv_all) and (iv) the number of private LC hits among neuro-associated proteins (n_priv_neuro).

ROC curves were generated for each burden metric using LC status as the binary outcome. The area under the ROC curve (AUC) was computed with 95% confidence intervals using DeLong’s method. Standard errors were estimated from the 95% CI width, and z-statistics were used to test whether AUC values differed significantly from 0.5 (random classification). For visualization, ROC curves were plotted with specificity on the x-axis and sensitivity on the y-axis, including a diagonal reference line and annotations summarizing AUC, standard error, 95% CI, and p-value.

### Antibody Pull-Down Followed by Mass Spectrometry Batch Effect Assessment and Correction

To assess and correct for batch effects in the proteomics dataset, we analyzed log₂-transformed protein expression data; missing values imputed as 0. Principal Component Analysis (PCA) was performed using the FactoMineR package to visualize variance across samples.

PERMANOVA test was conducted using the adonis2 function from the vegan package to quantify the proportion of variance in the expression matrix explained by batch, age, and sex. PCA plots colored by batch were generated with factoextra. To preserve biological variation across groups while correcting for batch effects, we used the empirical Bayes ComBat method implemented in the sva R package ^86^. Covariates included age, sex (encoded as binary), and clinical status (HC, CVC, or LC). We further quantified residual batch effects using PERMANOVA (adonis2 from the vegan package), with Euclidean distances computed on the corrected expression matrix and batch label used as the independent variable. After correction, PERMANOVA test results were: R^2^ = 0.034 with F = 0.83 and p-value = 0.516.

To quantify protein auto-reactivity across clinical groups, we calculated the mean and standard deviation (SD) for each protein among HC samples. We used these values to compute a Z-score for each observation. Proteins were considered positive in a sample if their Z-score exceeded a defined threshold (Z > 5). We then computed the number of positive proteins per sample. Group differences in the number of positive proteins were assessed using the Kruskal-Wallis test followed by Dunn’s post hoc test with Benjamini-Hochberg FDR correction. Significant pairwise comparisons (FDR < 0.05) were annotated on plots.

### Antibody Pull-Down Followed by Mass Spectrometry Pathway Enrichment Analysis of Private Hits for Long COVID

To identify biological pathways enriched among proteins uniquely detected in LC samples, we performed Gene Ontology (GO) Biological Process (BP) enrichment analysis. Gene symbols corresponding to LC-specific proteins were mapped to Entrez Gene IDs using the bitr() function from the clusterProfiler R package and the org.. Hs.eg.db annotation database (v3.18.0). GO BP enrichment analysis was then conducted using enrichGO() with the following parameters: ontology = “BP”, multiple testing correction via the Benjamini-Hochberg method, and a q-value significance cutoff of 0.05. The enrichment was performed using all annotated human genes as the background universe. GO terms associated with neurological processes (e.g., involving “neuro”, “synap”, “brain”, etc.) were identified by keyword filtering of term descriptions. To visualize the results, the top 15 significantly enriched GO BP terms were plotted using a custom ggplot2-based dot plot. In this plot, the x-axis represents the number of LC proteins annotated to each enriched GO term (“Protein Count”), point color encodes the adjusted p-value (p.adjust), and point size reflects the number of associated proteins.

### Mass spectrometry autoantibody burden ROC analysis

To evaluate whether the overall autoantibody burden or LC-private antibodies could discriminate LC from control samples, we performed receiver operating characteristic (ROC) analysis using the PROC package in R, restricted to LC cases and controls (HC + CVC). For classification analyses, we restricted the dataset to LC cases and controls, where controls were defined as HC + CVC, and created a binary outcome variable (LC = 1, control = 0). We then derived two per-sample burden metrics: (i) the total number of positive proteins across all measured proteins, and (ii) the number of LC-private proteins positive per sample. LC-private proteins were defined at the protein level as those that were positive in at least one LC sample and never positive in any control sample in the chosen comparison.

For each predictor, ROC curves were generated with LC status as the response and the burden metric as the predictor. The area under the ROC curve (AUC) was calculated, and if an initial AUC was <0.5, the predictor was inverted so that higher values corresponded to higher LC probability. Ninety-five percent confidence intervals for the AUC were obtained using DeLong’s method, and standard errors were estimated from the CI width. Z-statistics were then used to test whether observed AUC values differed significantly from 0.5 (random classification), and corresponding p-values were reported. For visualization, ROC curves were plotted with specificity on the x-axis and sensitivity on the y-axis, including a diagonal reference line and in-plot annotations summarizing AUC, standard error, 95% confidence interval, and p-value.

### Cross-platform overlap analysis of high-burden autoantibodies patients

To compare high-burden Long COVID patients across platforms, we integrated four assay-level summaries: GPCR autoantibody panel, HuProt autoantibody array, tissue immunofluorescence, and mass spectrometry–based autoantibody profiling. High-burden status was defined using fixed, assay-specific thresholds chosen *a priori*: For GPCR and Ionotropic Receptor panel ≥2 autoantibody hits; For HuProt array ≥100 positive HuProt proteins; For Immunofluorescence ≥2 IF-positive tissues; For Mass spectrometry: ≥20 positive protein hits. For each dataset, patients were classified as high burden if the number of hits met or exceeded the corresponding threshold, and as tested if they had a non-missing value. We then performed a coverage-aware overlap analysis. For each pair of datasets, we restricted to patients tested in both assays and computed: (i) the number of patients high-burden in both datasets, (ii) the number high in each assay separately, (iii) an agreement rate (defined as the proportion of co-tested patients classified as high burden in both assays, normalized by the total number of co-tested patients),(iii) and (iv) a Jaccard index for high burden (defined as the number of co-tested patients classified as high burden in both assays divided by the number of co-tested patients classified as high burden in at least one of the two assays). To summarize cross-platform burden at the individual level, we constructed a wide matrix with one row per patient and one column per dataset, encoded as TRUE (tested and high-burden), FALSE (tested and not high-burden), or NA (not tested). For each patient, we calculated the number of datasets in which they were high burden and enumerated patients high in ≥2 assays, ≥3 assays, and all 4 assays. These results were exported as coverage-aware matrices and patient lists. For visualization, we generated a bar plots summarizing the number of patients high in ≥2, ≥3, and 4 assays, as well as bar charts of pairwise agreement rate and Jaccard index between datasets.

### HuProt–GPCR and Ionotropic Receptor cross-platform correlation analysis

To evaluate concordance between HuProt autoantibody reactivity and GPCR and Ionotropic Receptor-targeted autoantibodies, HuProt features were mapped to gene symbols and matched to corresponding antibody measurements in the GPCR and Ionotropic Receptor ELISA dataset. When multiple HuProt probes corresponded to the same gene, signal intensities were averaged to generate a single gene-level log₂ mean fluorescence intensity (MFI) per sample. Spearman rank correlation analyses were performed separately within each clinical group (HC, CVC, LC) to assess the association between HuProt gene-level reactivity and GPCR antibody levels. Correlations (ρ) were calculated only for comparisons with at least three samples per group and non-constant variables. For each antibody–gene pair, the number of samples (n), Spearman correlation coefficient (ρ), and two-sided p-value were obtained using the cor.test function. For visualization, scatter plots were generated for each antibody–gene pair within each group, displaying HuProt gene-level log₂ MFI on the x-axis and GPCR and Ionotropic Receptor antibody signal on the y-axis. A fitted linear regression line with a 95% confidence interval was overlaid, and each plot was annotated with the corresponding Spearman ρ and p-value.

### Cross-Platform Comparison of Private LC Autoantibody Targets in HuProt and Proteomics Datasets

To assess concordance between LC autoantibody targets identified by HuProt and mass spectrometry, we performed an integrated comparative analysis. Private LC hits were defined separately in each platform as described before, and a curated list of positive and private hits was used for analysis. Frequency of autoantibodies per positive and private hits was visualized using bar plots to compare the number of LC patients with positive hits per protein per platform. A Venn diagram was generated to compare the total number of private hits identified uniquely by each platform or shared between them.

### Statistical analysis in vivo experiments

To classify the mice that were positive or negative for a specific behavior, the threshold was defined as the mean of the healthy control group + 1 SD and is represented as a dashed line.

Multiple group comparisons were performed with one-way or two-way analysis of variance (ANOVA) followed by Tukey’s or Śídák post-hoc multiple comparisons test. The differences in the values obtained for two different groups were evaluated using Kruskal–Wallis test followed by Dunn’s post-hoc multiple comparisons test. The differences in the frequency of participants or mice that were positive for a specific symptom were evaluated by the Chi-square test. Analyses were performed using the GraphPad PRISM (Version 10.2.3; GraphPad, San Diego, CA, US).

---

**Supplementary Figure 1.**
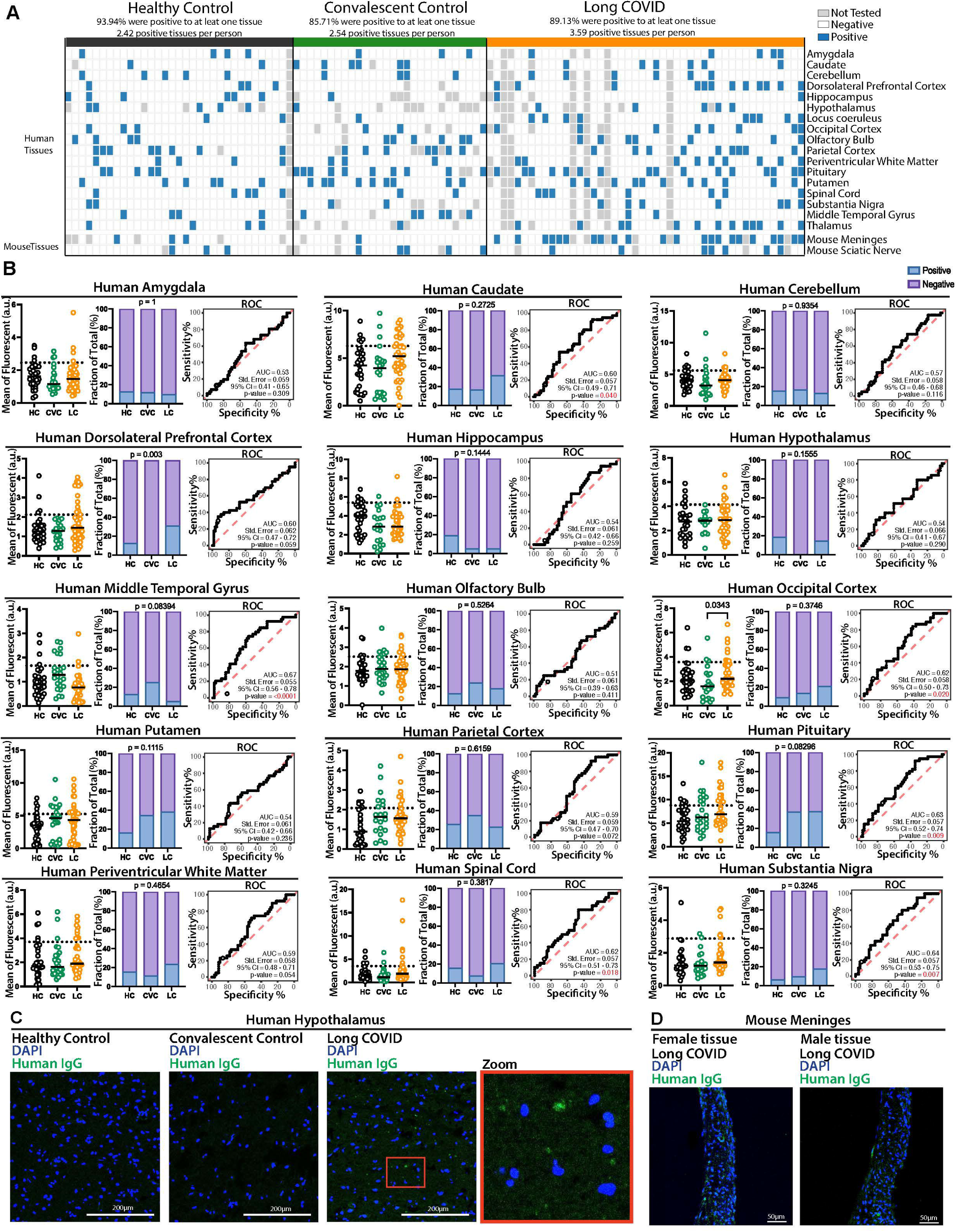
AAB profile for neurological tissues is diverse and increased in Long COVID participants. Confocal microscopy showing human and mouse tissues immunostained with human total IgG (green) purified from Long COVID, healthy or convalescent controls, as indicated, and nuclear DNA stain (DAPI, blue). **A.** Heatmap showing positive across different tissues between participants, each row is a different tissue, and each column is a different individual. **B.** Mean of fluorescence, percent of positivity, and Receiver operating characteristic (ROC) analysis for human amygdala, caudate, cerebellum, dorsolateral prefrontal cortex, hippocampus, hypothalamus, middle temporal gyrus, olfactory bulb, occipital cortex, putamen, parietal cortex, pituitary, periventricular white matter, spinal cord, and substantia nigra. **C.** Representative images of human hypothalamus immunostaining. Each dot in the figure represents the value obtained from an individual participant. Data are presented as the mean. **D.** Staining of meninges from a male and a female mouse using a sample from one patient previously considered as positive. Significant p-values are described in the image, as determined by Kruskal-Wallis test followed by Dunn’s post-hoc multiple comparisons test.

**Supplementary Figure 2.**
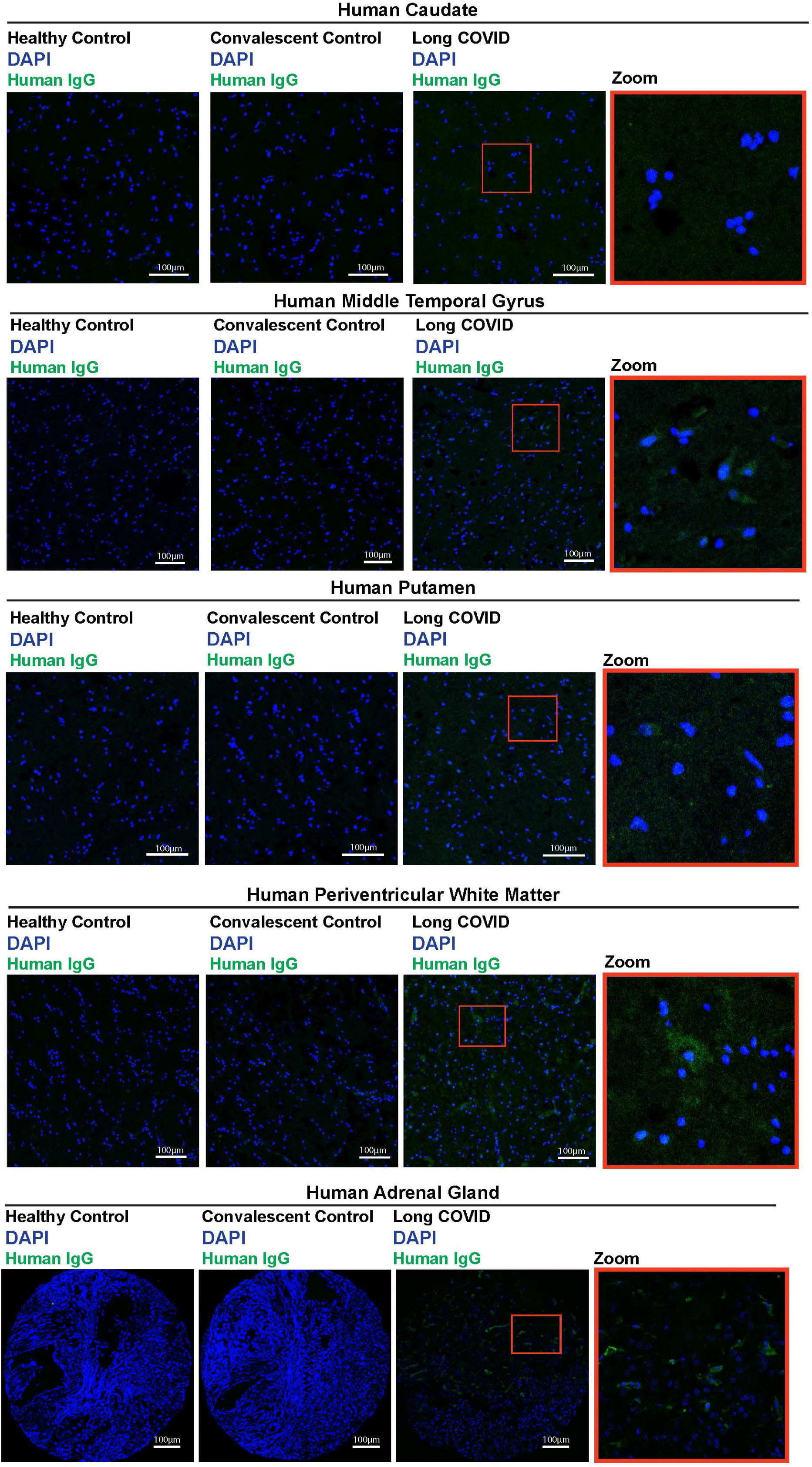
Representative images of immunofluorescence for neurological tissues and peripheral tissues. Confocal microscopy showing human caudate, middle temporal gyrus, putamen, white matter, and adrenal gland tissues immunostained with human total IgG (green) purified from Long COVID, healthy or convalescent controls, as indicated, and nuclear DNA stain (DAPI, blue).

**Supplementary Figure 3.**
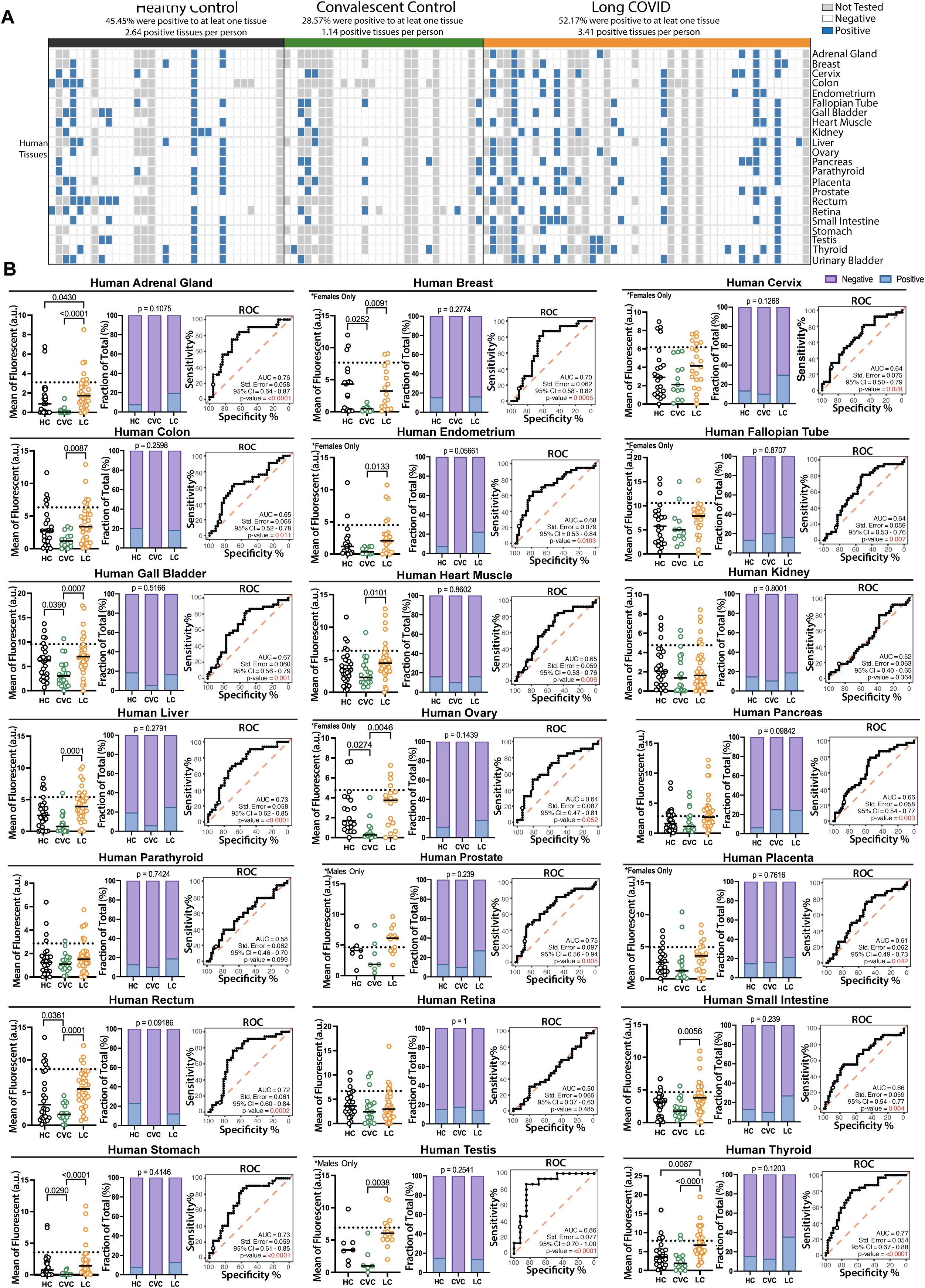
AAB profile for peripheral tissues is diverse and increased in Long COVID participants. Confocal microscopy showing human and mouse tissues immunostained with human total IgG (green) purified from Long COVID, healthy or convalescent controls, as indicated, and nuclear DNA stain (DAPI, blue). **A.** Heatmap showing positive across different tissues between participants, each row is a different tissue, and each column is a different individual. **B.** Mean of fluorescence, percent of positivity, and Receiver operating characteristic (ROC) analysis for human adrenal gland, breast, cervix, colon, endometrium, fallopian tube, gall bladder, heart muscle, kidney, liver, ovary, pancreas, parathyroid, prostate, placenta, rectum, retina, small intestine, stomach, testis, thyroid, urinary bladder. Each dot in the figure represents the value obtained from an individual participant. Data are presented as the mean. Significant p-values are described in the image, as determined by Kruskal-Wallis test followed by Dunn’s post-hoc multiple comparisons test.

**Supplementary Figure 4.**
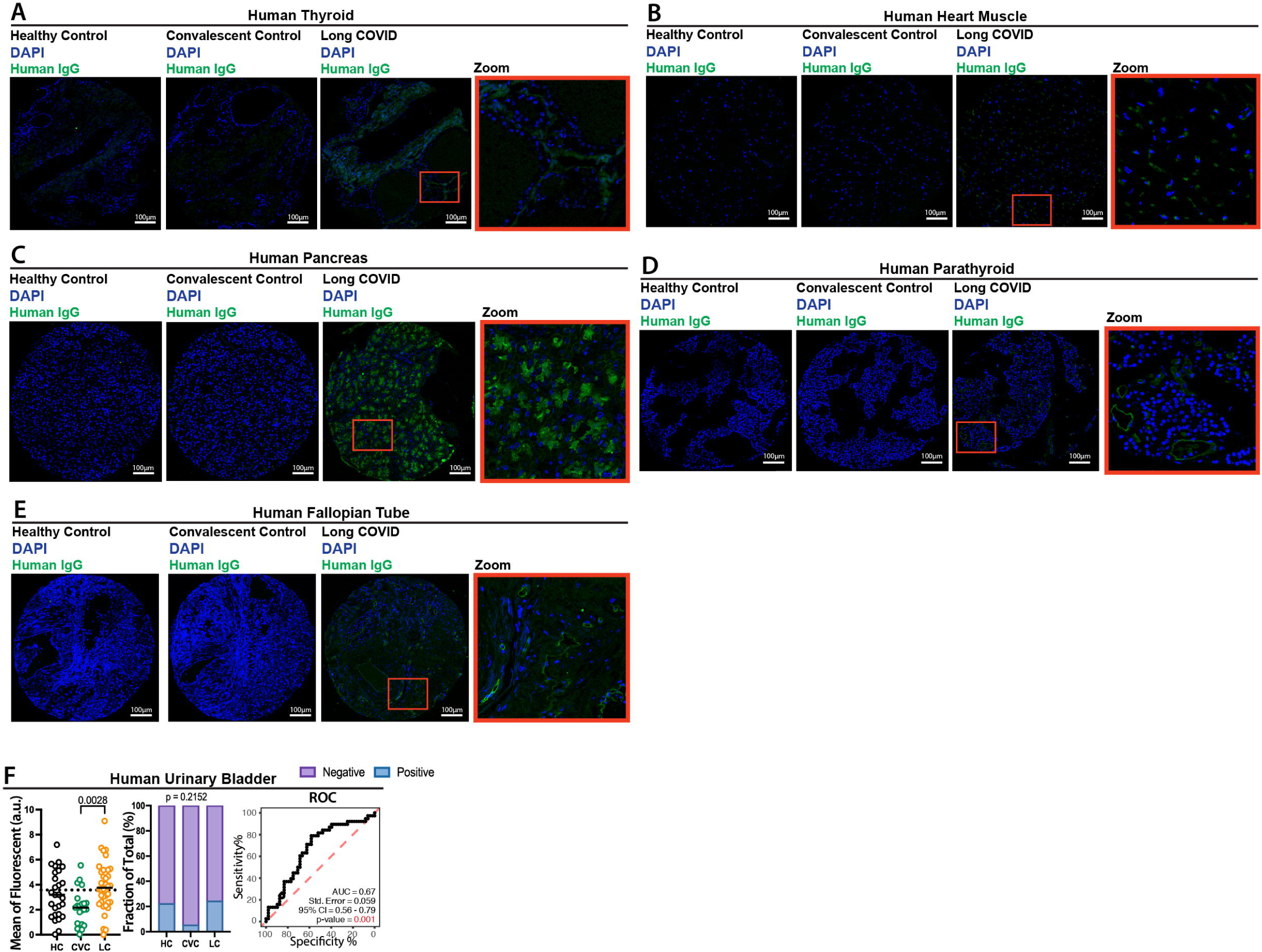
Representative images of immunofluorescence for peripheral tissues. A-E. Confocal microscopy showing human thyroid, heart muscle, pancreas, parathyroid, fallopian tube tissues immunostained with human total IgG (green) purified from Long COVID, healthy or convalescent controls, as indicated, and nuclear DNA stain (DAPI, blue). **F.** Mean of fluorescence, percent of positivity, and Receiver operating characteristic (ROC) analysis for human urinary bladder.

**Supplementary Figure 5.**
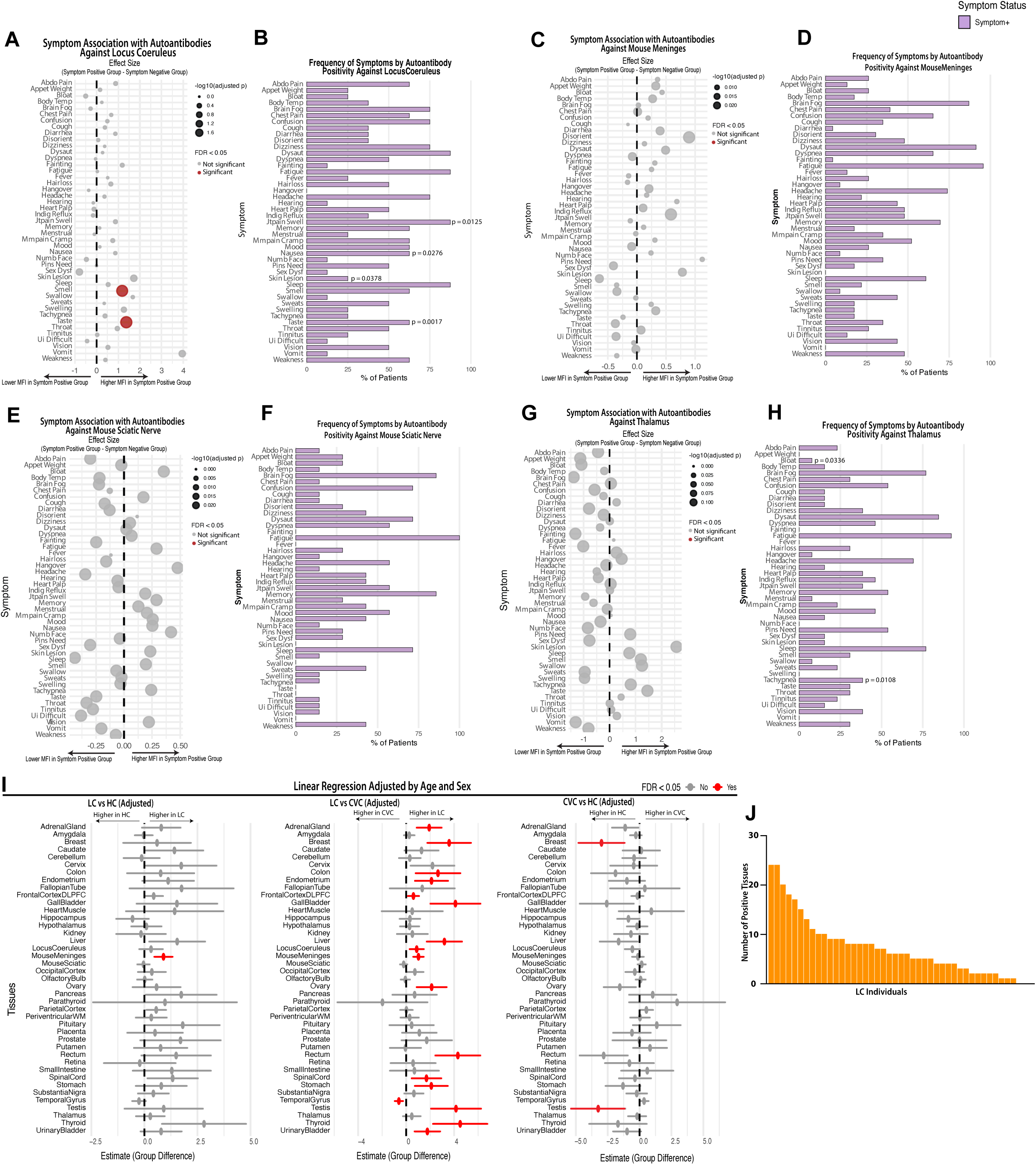
IgG from participants with long COVID that reacts to locus coeruleus is associated with symptoms. A, C, E, **G.** Dot plot graph showing the effect size of human locus coeruleus, mouse meninges, mouse sciatic nerve and human thalamus mean of fluorescence divided by symptoms displayed by the participants. Significant p-values are described in the image, determined by a two-sided Wilcoxon rank-sum test adjusted for multiple comparisons using the Benjamini–Hochberg false discovery rate (FDR). **B, D, F, G**. Frequency of symptoms displayed by the participants who had autoantibody reactivity against human locus, mouse meninges, mouse sciatic nerve and human thalamus coeruleus. Significant p-values are described in the image, determined by Fisher’s exact test. **I.** Linear regression modeling on immunofluorescence (IF) data obtained from tissue-based autoantibody staining. Graph showing estimated group effect, 95% confidence intervals, and associated significant p-values. P-values were adjusted for multiple testing using the Benjamini–Hochberg False Discovery Rate (FDR) method. **J**. Bar plot showing the number of tissues with positive autoantibody binding by immunofluorescence for each LC individual.

**Supplementary Figure 6.**
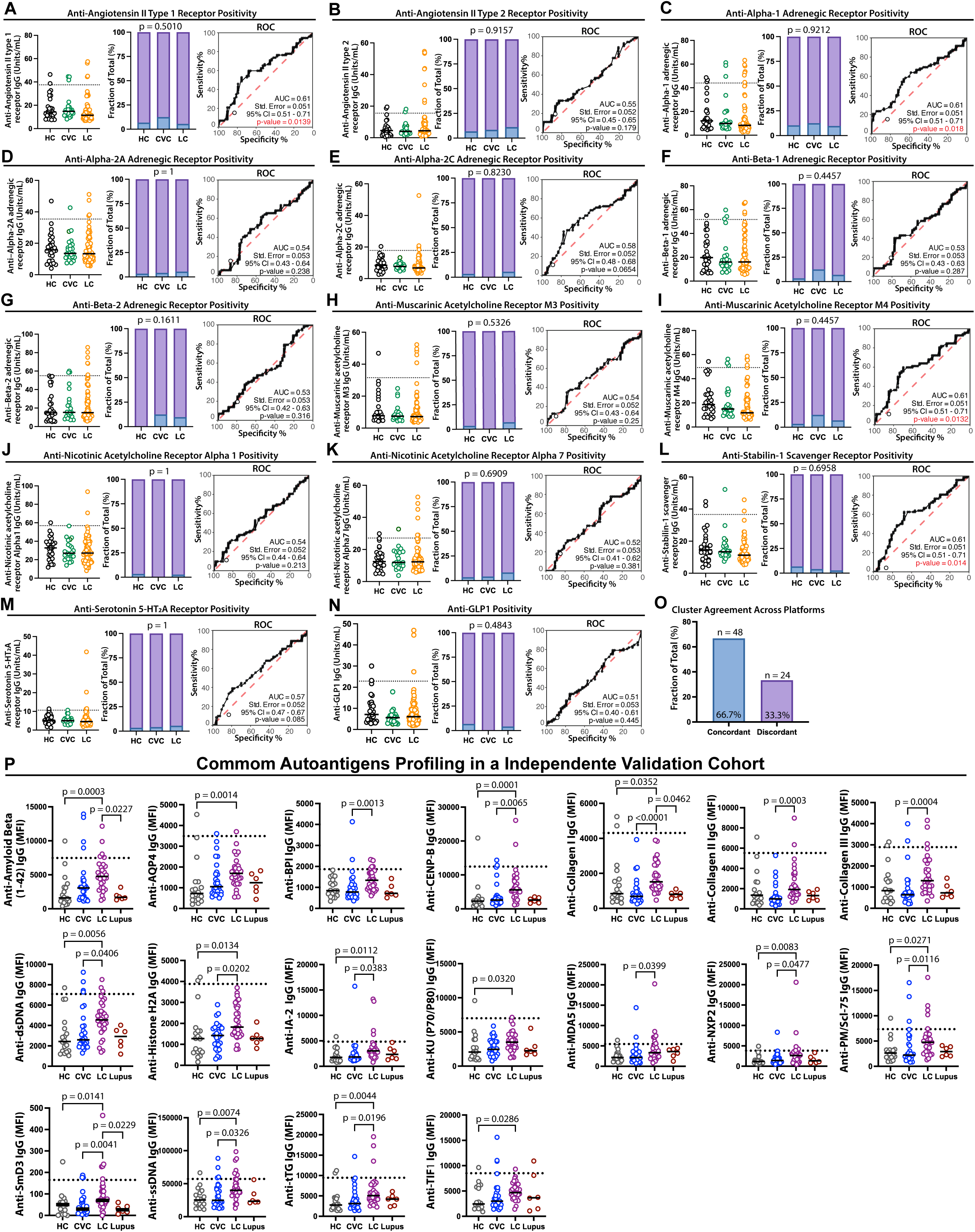
Autoantibody screening in LC individuals. A-N. ELISA for anti-GPCRs. Each dot represents one individual. P-values are described in the figure and were determined by the Kruskal-Wallis test followed by post hoc Dunn’s multiple comparisons test. **O.** Cluster assignment agreement between Huprot and GPCR ELISA. **P.** Protein microarray analysis performed in an independent validation cohort. Each dot represents one individual. Dashed lines indicate positivity thresholds defined as the mean of HC ± SD. P-values are shown in the figure and were determined using the Kruskal–Wallis test followed by Dunn’s post hoc multiple comparisons test.

**Supplementary Figure 7.**
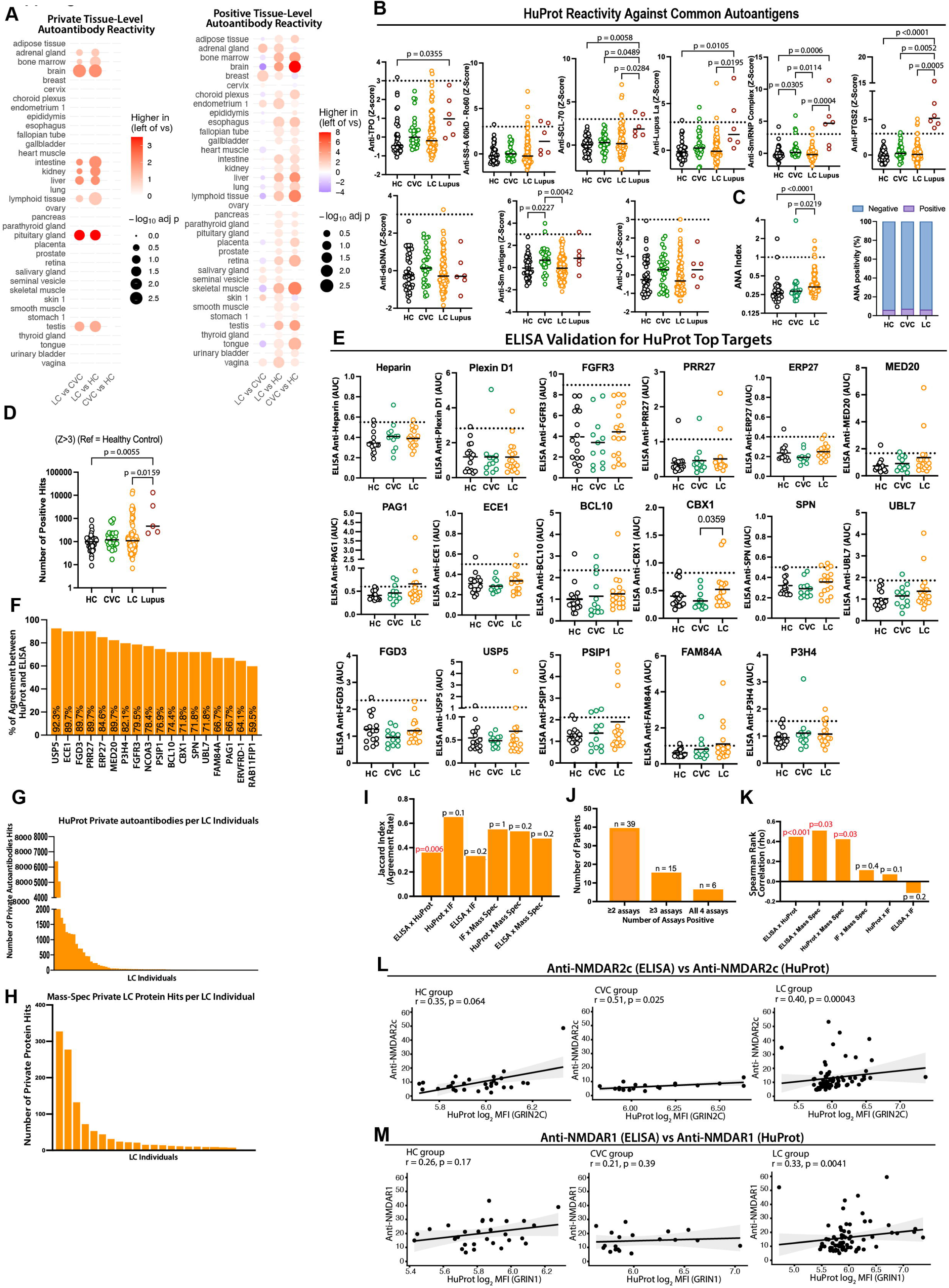
Autoantigen validation and common reactivity. **A**. Dot plot showing pairwise comparisons of tissue-level reactivity between groups; circle size represents the statistical significance (−log₁₀ adjusted *p*-value) and color indicates the direction and magnitude of difference in mean tissue reactivity determined by Wilcoxon rank-sum tests, followed by Benjamini-Hochberg correction for multiple testing (FDR). **B.** Number of HuProt auto-reactivities for each person against common autoantigens, p-values are described in the figure and were determined by the Kruskal-Wallis test followed by post hoc Dunn’s test. **C.** ANA index and ANA positivity. Each dot represents one individual. Dashed lines indicate positivity thresholds defined as the mean of HC ± 2 SD. P-values are described in the figure and were determined by the Kruskal-Wallis test followed by post hoc Dunn’s multiple comparisons test. **D.** Number of HuProt auto-reactivities for each person, compared with lupus patients, p-values are described in the figure and were determined by the Kruskal-Wallis test followed by post hoc Dunn’s test. **E.** ELISA and Area Under Curve (AUC) analysis for top hit targets identified by Huprot and mass spec. Each dot in the figure represents the value obtained from an individual person. **F**. Jaccard similarity analysis comparing HuProt and ELISA assays. **G**. Bar plot showing the number of private autoreactivities per LC individuals for HuProt. **H.** Bar plot showing the number of private autoreactivities per individual for mass-spec analysis. **I.** Jaccard index showing overlap of positive samples across platforms. **J.** Number of LC patients identified as positive in each assay. **K**. Threshould-free Spearman Rank-Based Correlation. **L-M.** Spearman correlation analyses between ELISA and discovery datasets for NMDAR reactivities: **L.** anti-NMDAR2C (ELISA) vs. anti-NMDAR2C (HuProt), **M** anti-NMDAR1 (ELISA) vs. anti-NMDAR1 (HuProt). All p-values were adjusted for multiple comparisons using the Benjamini-Hochberg method, FDR method.

**Supplementary Figure 8.**
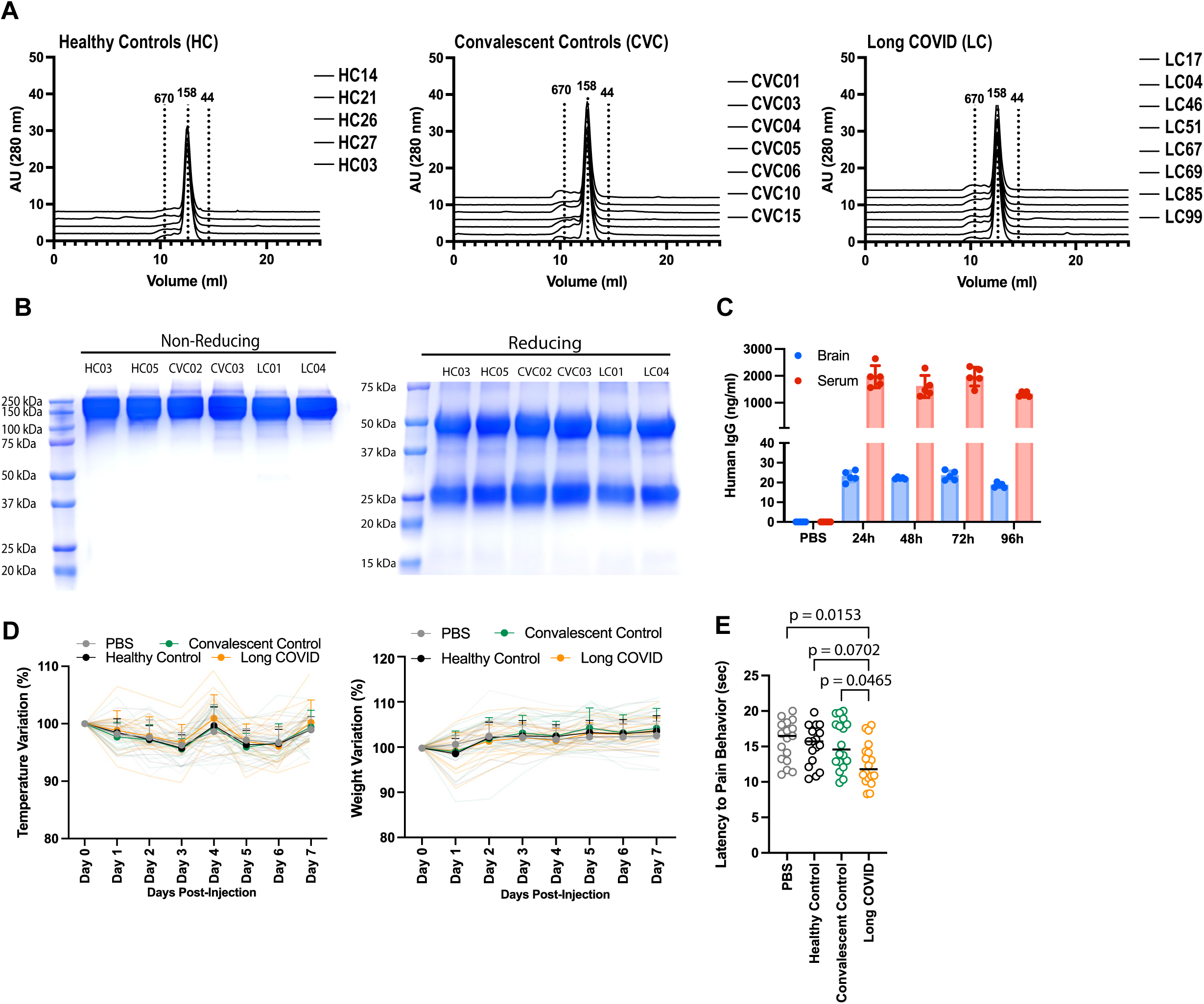
Quality analysis of purified hIgG. **A**. Size-exclusion chromatography (SEC) profiles of the purified proteins run in a Fast Protein Liquid Chromatography (FPLC). Purity was determined by peak integration with the ChromLab Software. **B**. Non-reducing and reducing SDS-PAGE gel stained with Coomassie Blue showing hIgG purified from participants. **C**. Time course analysis of human IgG concentration by ELISA in brain homogenate and serum for four days post-injection. **D**. Temperature and weight variation longitudinal analysis after passive transfer of hIgG in mice. **E.** Hot plate analysis performed after passive transfer by a blinded independent observer. Each dot in the figure represents the value obtained from an individual mouse. Data are presented as the mean ±SD. Significant p values are described in the image, as determined by a T-test for two-group comparisons or one-way ANOVA followed by Tukey’s post-hoc multiple comparisons test.

**Supplementary Figure 9.**
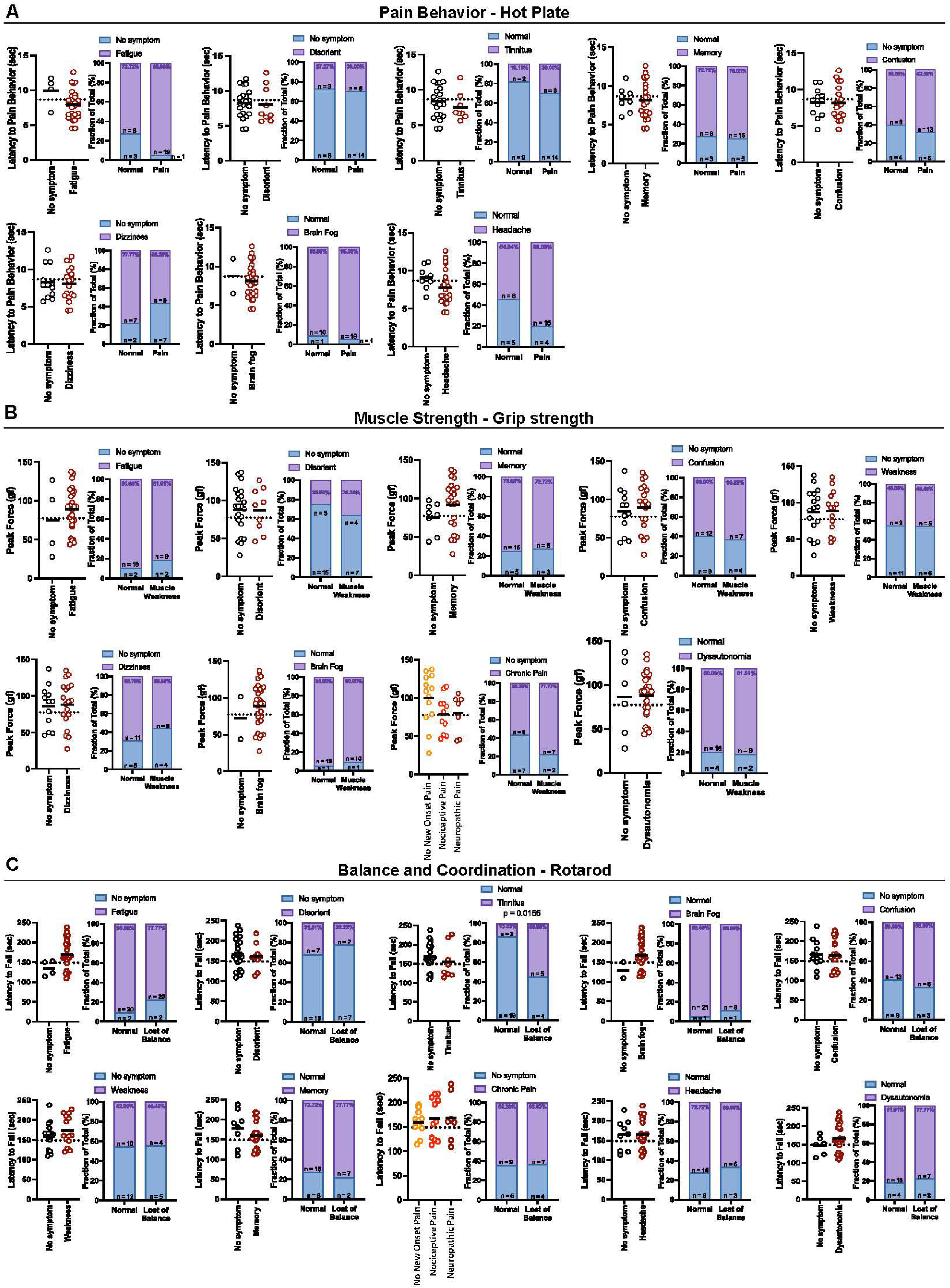
Symptoms evaluated in the passive transfer that were not associated with the hot plate test and grip strength. Doses of 38.4 mg/kg of total IgG purified from healthy, convalescent controls and Long COVID participants were administered to 6-8 weeks-old C57BLl/6 female mice by intraperitoneal (IP) injection, which were followed up for 5 days. **A.** Hot plate test divided by symptoms displayed by the participants, such as fatigue, disorientation, tinnitus, memory, confusion, dizziness, brain fog, and headache. **B.** Grip strength test was divided by participants who displayed fatigue, disorientation, memory, confusion, weakness, dizziness, brain fog, chronic pain, and dysautonomia. **C**. Rotarod test divided by symptoms displayed by the participants, such as fatigue, disorientation, tinnitus, brain fog, confusion, weakness, memory, chronic pain, headache, and dysautonomia. Each dot in the figure represents the value obtained from an individual mouse. Each mouse received antibodies isolated from a single human participant. Data are presented as the mean. Significant p values are described in the image, as determined by one-way ANOVA followed by Tukey’s post-hoc multiple comparisons test.

**Supplementary Figure 10.**
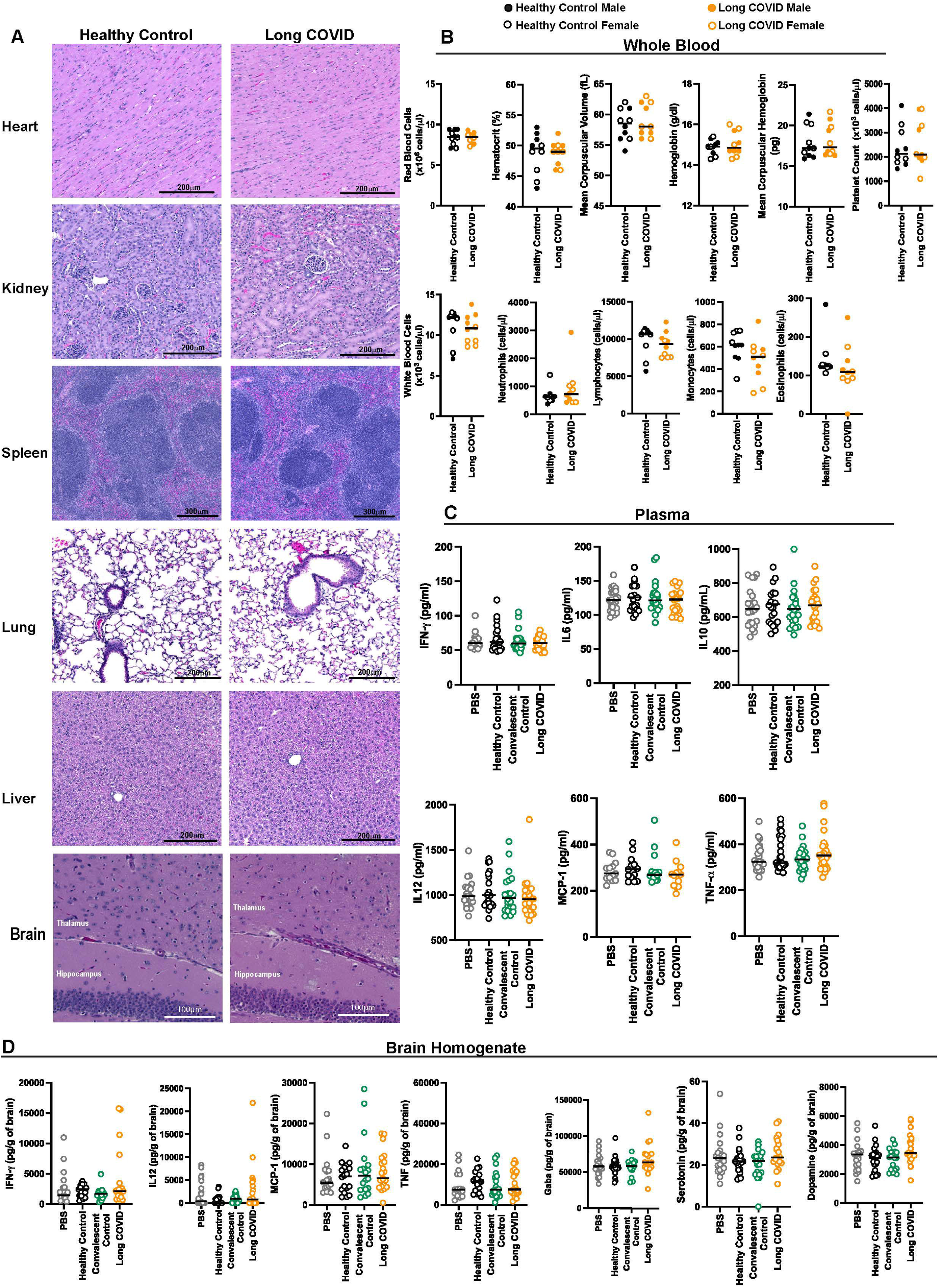
Mice necropsy, whole blood cells, neurotransmitters, and cytokine analysis. A dose of 38.4 mg/kg of total IgG purified from healthy and Long COVID participants was administered to 7-week-old C57BL/6 female and male mice by intraperitoneal (IP) injection. The mice were euthanized after five days for necropsy, whole blood cells, and plasma cytokine analysis. Mice were euthanized 3 days post-injection for brain homogenate cytokines and neurotransmitters analysis. **A.** Representative images of hematoxylin and eosin heart, kidney, spleen, lung, and liver of mice at day 5 post-injection. All histological images were captured by K.S.G.S using BioTek Cytation 5 imaging reader (Agilent). **B.** Whole blood cells count 5 days post-injection. **C.** Cytokine profile in plasma 5 days post-injection. **D.** Neurotransmitters and cytokine profile in brain homogenate 3 days post-injection. Each dot in the figure represents the value obtained from an individual mouse. Each mouse received antibodies isolated from a single human participant. Data are presented as the mean. Significant p values are described in the image, as determined by a T-test for two-group comparisons or one-way ANOVA followed by Tukey’s post-hoc multiple comparisons test.

**Supplementary Figure 11.**
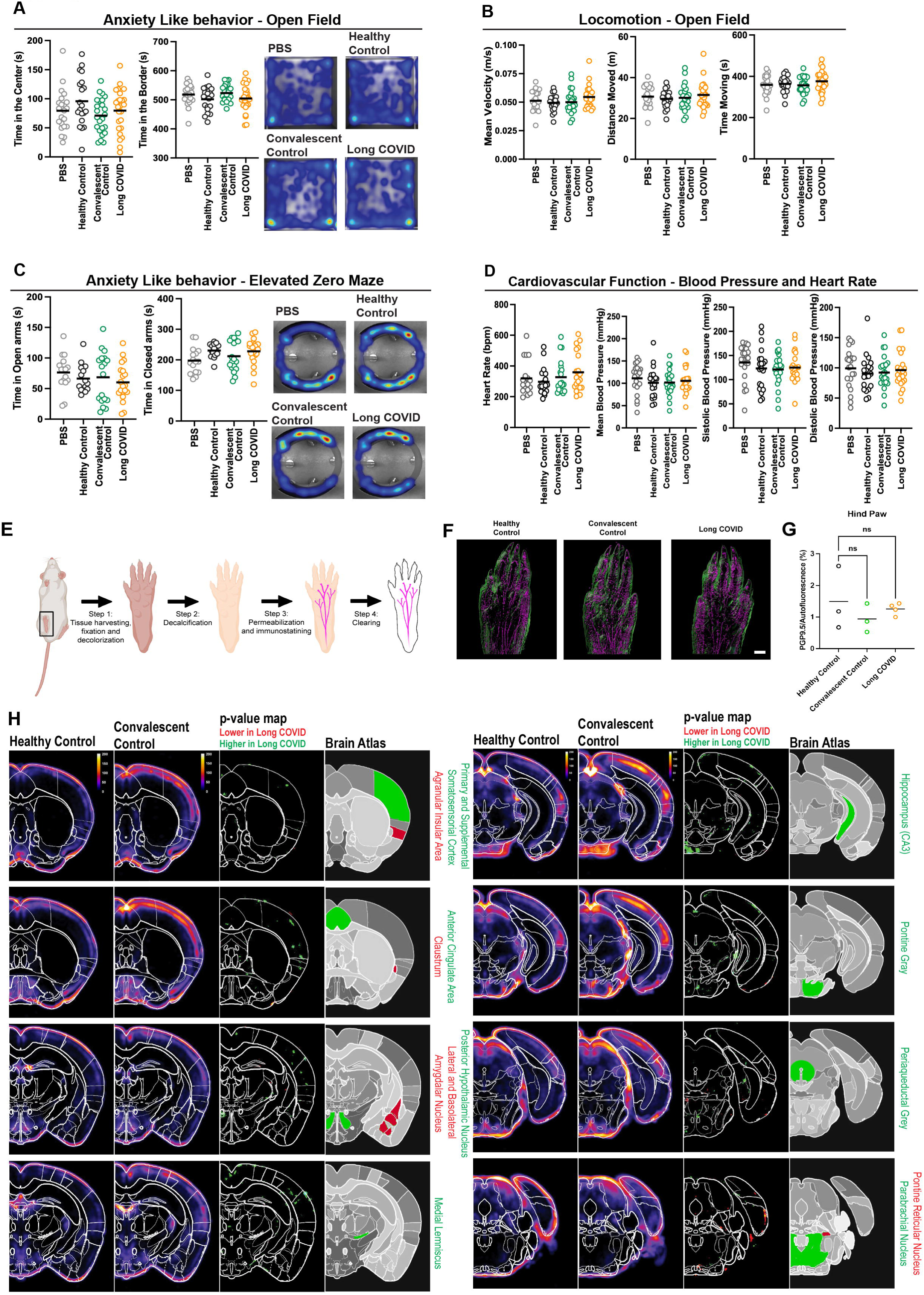
Passive transfer does not induce anxiety-like behavior, locomotion, and cardiovascular function symptoms in mice. A dose of 38.4 mg/kg of total IgG purified from healthy, convalescent controls and Long COVID participants was administered to 6-8 weeks-old C57BL/6 female mice by intraperitoneal (IP) injection, which were followed up for 5 days. **B**. Cumulative time spent in the center, cumulative time spent in the border, and representative heat map showing mice movement in the open field test. **C**. Mean velocity, distance moved, and time moving during the open field test. **D**. Cumulative time in open arms, cumulative time in closed arms, and representative heatmap showing mice movement in the elevated zero maze test. **E**. Heart rate, mean blood pressure, systolic blood pressure, and diastolic blood pressure measurements. Each dot in the figure represents the value obtained from an individual mouse. Each mouse received antibodies isolated from a single human participant. Data are presented as means. Significant p values are described in the image, as determined by one-way ANOVA followed by Tukey’s post-hoc multiple comparisons test. **E.** Schematic for whole hind paw PGP9.5 stain for mice after passive transfer of hIgG. **F.** PGP9.5 mapping results for the hind paw. **G.** Percent mean of fluorescence. **H.** Fos mapping results for brain areas comparing healthy controls and convalescent controls, first two columns showing heatmap with Fos intensity, third column represents p-value maps by Kruskal-Wallis test, followed by Dunn’s multiple comparison test, last column represents Allen Brain annotation. The heatmaps are the average of all mice in each group (n = 6 per group). Each dot in the figure represents the value obtained from an individual mouse. Each mouse received antibodies isolated from a single human participant. Data are presented as means. Significant p values are described in the image, as determined by one-way ANOVA followed by Tukey’s post-hoc multiple comparisons test.

