## Supplementary material for "A causal link between autoantibodies and neurological symptoms in long COVID": Supp Table 2

**Supplementary Table 2: Clinical Demographics of My-LC-II Cohort**

| Demographics | Healthy Control Mean (±SD) or N(%) | Convalescent Control Mean (±SD) or N(%) | Long COVID Mean (±SD) or N(%) | p-value* |
|---|---|---|---|---|
| Cohort Size | 18 | 29 | 32 | |
| Age | 43.41 (±14.37) | 37.07 (±9.49) | 38.84 (±8.36) | 0.16** |
| Sex | | | | |
| Female | 8 (47.1%) | 19 (65.5%) | 20 (62.5%) | 0.44 |
| Male | 9 (52.9%) | 10 (34.5%) | 12 (37.5%) | 0.44 |
| BMI | 26.38 (5.15) | 26.79 (±5.03) | 24.54 (±4.91) | 0.051** |
| Race | | | | |
| White | 16 (94.1%) | 25 (86.2%) | 24 (75.0%) | 0.51* |
| Asian | 1 (5.9%) | 3 (10.3%) | 4 (12.5%) | 0.51* |
| Other | 0 | 1 (3.4%) | 4 (12.5%) | 0.51* |
| Ethnicity | | | | |
| Hispanic or Latino | 0 | 2 (6.9%) | 1 (3.1%) | 0.60* |
| COVID-19 Clinical testing | | | | |
| Only PCR positive test | 0 | 12 (52.2%) | 4 (14.8%) | |
| Only Ab Positive Test | 0 | 5 (21.7%) | 8 (29.6%) | <0.0001♦ |
| PCR and Ab Positive Test | 0 | 6 (26.1%) | 15 (55.6%) | |
| Not Tested | 17 | 0 | 0 | |
| Acute COVID-19 Hospitalized | 0 | 0 | 1 (3.1%) | 1 |
| Days from Acute Infection | 0 | 372.03 (±258.06) | 632.59 (±309.69) | 0.0012**♦ |
| Vaccination Status | | | | |
| Total number of doses | 2.82 (±0.39) | 3.65 (±0.76) | 3.25 (±1.58) | 0.81**● |
| Past Medical History | | | | |
| Hypertension | 3 (17.6%) | 2 (6.9%) | 0 | 0.035 |
| Diabetes | 0 | 1 (3.4%) | 0 | 0.58 |
| Chronic Kidney Disease | 0 | 0 | 1 (3.1%) | 1 |
| Asthma | 3 (17.6%) | 4 (13.8%) | 2 (6.2%) | 0.42 |
| Chronic Lung Disease | 1 (5.9%) | 0 | 0 | 0.21 |
| Obesity | 0 | 3 (10.3%) | 1 (3.1%) | 0.34 |
| Immunodeficiency | 0 | 1 (3.4%) | 0 | 0.58 |
| Cancer | 2 (11.8%) | 1 (3.4%) | 1 | |
| Anxiety | 5 (29.4%) | 10 (34.5%) | 6 (18.8%) | 0.40 |
| Depression | 6 (35.3%) | 6 (20.7%) | 5 (15.6%) | 0.29 |
| Psychiatric Disease | 0 | 0 | 3 (3.9%) | 0.23 |
| Inflammatory Bowel Disease | 0 | 3 (10.3%) | 7 (21.9%) | 0.08 |
| EQ-5D Visual Analog Scale | | | | |
| Mobility problems | 0 | 1 (3.4%) | 22 (68.8%) | <0.001● |
| Self-care problems | 2 (11.8%) | 2 (6.9%) | 27 (84.4%) | <0.001● |
| Usual activities difficulty | 8 (47.1%) | 13 (44.8%) | 21 (65.6%) | 0.081● |
| Pain/Discomfort | 1 (5.9%) | 0 | 14 (43.8%) | <0.001● |
| Anxiety/Depression | 5 (29.4%) | 7 (24.1%) | 29 (90.6%) | <0.001● |
| Overall Health Good | 17 (100%) | 25 (86.2%) | 5 (15.6%) | <0.001● |
| Overall Health Fair | 0 | 4 (13.8%) | 11 (34.4%) | <0.001● |
| Overall Health Poor | 0 | 0 | 16 (50.0%) | <0.001● |
| Modified Medical Research Council (mMRC) Dyspnea Scale | | | | |

| | | | | |
|---|---|---|---|---|
| Mild | 15 (88.2%) | 26 (89.7%) | 8 (25.0%) | <0.001• |
| Moderate | 2 (11.8%) | 3 (10.3%) | 11 (34.4%) | <0.001• |
| Severe | 0 | 0 | 13 (40.6%) | <0.001• |
| Fatigue Severity Scale (FSS) | 2.12 (±0.66) | 2.12 (±0.85) | 7.4 (±9.92) | <0.001**• |
| Depression screening PHQ-2 | | | | |
| Anhedonia | 4 (23.5%) | 6 (20.7%) | 16 (50.5%) | 0.03 |
| Depressed Mood | 7 (41.2%) | 9 (31.0%) | 18 (56.2%) | 0.13 |
| Depression Positive Screen | 0 | 1 (3.4%) | 10 (31.2%) | 0.0011 |
| Anxiety screening (GAD-7) | | | | |
| Mild | 4 (23.5%) | 4 (13.8%) | 7 (21.9%) | 0.09 |
| Moderate | 0 | 1 (3.4%) | 4 (12.5%) | 0.09 |
| Severe | 0 | 0 | 4 (12.5%) | 0.09 |
| Pain intensity (visual analog scale) | | | | |
| Normal | 17 (100%) | 29 (100%) | 5 (22.7%) | <0.001 |
| Mild | 0 | 0 | 6 (27.3%) | |
| Severe | 0 | 0 | 11 (50.0%) | |
| PROMIS Sleep disturbance score | | | | |
| Normal | 3 (17.6%) | 2 (6.9%) | 13 (40.6%) | <0.001 |
| Moderate | 5 (29.4%) | 5 (17.2%) | 8 (25.0%) | |
| Severe | 9 (52.9%) | 22 (75.9%) | 11 (34.4%) | |
| Post-Exertional Malaise (PEM) | 0 | 2 (6.9%) | 27 (84.4%) | <0.001 |

* Chi-square or Fisher test

** Kruskal-Wallis test

♦ Comparison performed only between CVC and LC groups

• Comparison performed between Controls (HC+CVC) and LC groups
